# Safety and Immunogenicity of An Egg-Based Inactivated Newcastle Disease Virus Vaccine Expressing SARS-CoV-2 Spike: Interim Results of a Randomized, Placebo-Controlled, Phase 1/2 Trial in Vietnam

**DOI:** 10.1101/2022.02.01.22270253

**Authors:** Anh Duc Dang, Thiem Dinh Vu, Ha Hai Vu, Van Thanh Ta, Anh Thi Van Pham, Mai Thi Ngoc Dang, Be Van Le, Thai Huu Duong, Duoc Van Nguyen, Saranath Lawpoolsri, Pailinrut Chinwangso, Jason S. McLellan, Ching-Lin Hsieh, Adolfo Garcia-Sastre, Peter Palese, Weina Sun, Jose L. Martinez, Irene Gonzalez-Dominguez, Stefan Slamanig, Juan Manuel Carreño, Johnstone Tcheou, Florian Krammer, Ariel Raskin, Huong Minh Vu, Thang Cong Tran, Huong Mai Nguyen, Laina D. Mercer, Rama Raghunandan, Manjari Lal, Jessica A. White, Richard Hjorth, Bruce L. Innis, Rami Scharf

## Abstract

Production of affordable coronavirus disease 2019 (COVID-19) vaccines in low- and middle-income countries is needed. NDV-HXP-S is an inactivated egg-based Newcastle disease virus (NDV) vaccine expressing the spike protein of severe acute respiratory syndrome coronavirus 2 (SARS-CoV-2) Wuhan-Hu-1. The spike protein was stabilized and incorporated into NDV virions by removing the polybasic furin cleavage site, introducing the transmembrane domain and cytoplasmic tail of the fusion protein of NDV, and introducing six prolines for stabilization in the prefusion state. Vaccine production and clinical development was initiated in Vietnam, Thailand, and Brazil. Here the interim results from the first stage of the randomized, dose-escalation, observer-blind, placebo-controlled, phase 1/2 trial conducted at the Hanoi Medical University (Vietnam) are presented. Healthy adults aged 18-59 years, non-pregnant, and with self-reported negative history for SARS-CoV-2 infection were eligible. Participants were randomized to receive one of five treatments by intramuscular injection twice, 28 days apart: 1 μg +/-CpG1018 (a toll-like receptor 9 agonist), 3 μg alone, 10 μg alone, or placebo. Participants and personnel assessing outcomes were masked to treatment. The primary outcomes were solicited adverse events (AEs) during 7 days and subject-reported AEs during 28 days after each vaccination. Investigators further reviewed subject-reported AEs. Secondary outcomes were immunogenicity measures (anti-spike immunoglobulin G [IgG] and pseudotyped virus neutralization). This interim analysis assessed safety 56 days after first vaccination (day 57) in treatment-exposed individuals and immunogenicity through 14 days after second vaccination (day 43) per protocol. Between March 15 and April 23, 2021, 224 individuals were screened and 120 were enrolled (25 per group for active vaccination and 20 for placebo). All subjects received two doses. The most common solicited AEs among those receiving active vaccine or placebo were all predominantly mild and included injection site pain or tenderness (<58%), fatigue or malaise (<22%), headache (<21%), and myalgia (<14%). No higher proportion of the solicited AEs were observed for any group of active vaccine. The proportion reporting vaccine-related AEs during the 28 days after either vaccination ranged from 4% to 8% among vaccine groups and was 5% in controls. No vaccine-related serious adverse event occurred. The immune response in the 10 μg formulation group was highest, followed by 1 μg +CpG1018, 3 μg, and 1 μg formulations. Fourteen days after the second vaccination, the geometric mean concentrations (GMC) of 50% neutralizing antibody against the homologous Wuhan-Hu-1 pseudovirus ranged from 56.07 IU/mL (1 μg, 95% CI 37.01, 84.94) to 246.19 IU/mL (10 μg, 95% CI 151.97, 398.82), with 84% to 96% of vaccine groups attaining a ≥ 4-fold increase over baseline. This was compared to a panel of human convalescent sera (N=29, 72.93 95% CI 33.00-161.14). Live virus neutralization to the B.1.617.2 (Delta) variant of concern was reduced but in line with observations for vaccines currently in use. Since the adjuvant has shown modest benefit, GMC ratio of 2.56 (95% CI, 1.4 - 4.6) for 1 μg +/-CpG1018, a decision was made not to continue studying it with this vaccine. NDV-HXP-S had an acceptable safety profile and potent immunogenicity. The 3 μg dose was advanced to phase 2 along with a 6 μg dose. The 10 μg dose was not selected for evaluation in phase 2 due to potential impact on manufacturing capacity. ClinicalTrials.gov NCT04830800.

## 1. Introduction

A considerable imbalance remains in the global distribution of coronavirus disease 2019 (COVID-19) vaccines, with access in low- and middle-income countries (LMIC) considerably lagging behind (1). Control of the COVID-19 pandemic in LMICs, where 75% of the global population resides, will be achieved only when a sustainable supply of affordable vaccines can be secured. The manufacturing capacity for egg-based inactivated influenza vaccines (IIV) is constitutes some of the largest vaccine production capacity in the world. These facilities, some in middle-income countries and operating for less than six months per year, use locally produced embryonated eggs to make more than a billion doses annually of affordable human vaccines (2). To enable these manufacturers to respond to the COVID-19 pandemic by harnessing their experience with IIV and utilizing existing infrastructure, we developed a COVID-19 vaccine for production in eggs, based on a Newcastle disease virus (NDV) expressing the ectodomain of a novel membrane-anchored, prefusion-stabilized severe acute respiratory syndrome coronavirus 2 (SARS-CoV-2) Wuhan-Hu-1 spike protein construct. This virus (NDV-HXP-S) is purified from allantoic fluid, inactivated by betapropriolactone (BPL), and then formulated (3-5).

From September to November 2020, manufacturers in Vietnam, Thailand, and Brazil modified their IIV manufacturing process to optimize production of BPL-inactivated NDV-HXP-S, achieving high yields at pilot scale; the result was three similar processes. Preclinical evaluation of their vaccine candidates, formulated with and without CpG1018, a toll-like receptor 9 (TLR-9) agonist adjuvant (Dynavax Technologies) (6) confirmed that they were highly immunogenic and protective in hamsters (3,5) with no sign of toxicity in rats at the maximum human doses planned for evaluation (3 μg spike protein+1·5 mg CpG1018; 10 μg spike protein) (manuscript in preparation). All three manufacturers initiated clinical development of their vaccine candidates and the interim analysis from Thailand is available (7). Herein, we report interim safety and immunogenicity data generated in the phase 1 portion of a phase 1/2 clinical trial evaluating the NDV-HXP-S vaccine candidate (COVIVAC) developed by the Vietnam Institute of Vaccines and Medical Biologicals (IVAC). The clinical development program for the NDV-HXP-S vaccine candidate in Vietnam began in March 2021, The Vietnamese government received its first AstraZeneca COVID-19 vaccines in February 2021 and then the BBIBP-CorV (Vero Cells) vaccine from Sinopharm in June 2021. These products were authorized for emergency use by the Vietnamese Ministry of Health and administered to health care personnel, older adults, and other high-risk groups. Our aim is to attain authorisation for the NDV-HXP-S vaccine candidate as soon as possible to supply a domestically produced, affordable vaccine for COVID-19 prevention and control.

These results provide additional evidence in humans that the recombinant NDV technology expressing a six-proline prefusion-stabilized spike protein offers a unique platform for affordable manufacturing of a well-tolerated and highly immunogenic COVID-19 vaccine.

## 2. Materials and methods

### Study design and participants

The phase 1 portion of a phase 1/2 randomized, observer-blind, placebo-controlled trial was conducted at Hanoi Medical University (Hanoi, Vietnam). Participants were recruited from individuals known to the university and through advertisements. Healthy adults 18–59 years of age with body mass index 17 to 40 kg/m^2^, negative for hepatitis B surface antigen, without known history of SARS-CoV-2 infection, HIV, and hepatitis C, were eligible to participate. A negative urinary pregnancy test was required of women having reproductive capacity prior to administration of each study vaccine dose. Complete eligibility criteria are described in the trial protocol provided in Appendix A. Written informed consent was obtained from all participants. The trial complied with the Declaration of Helsinki and Good Clinical Practice. This study was jointly approved by the Institutional Review Board of the Vietnam National Institute of Hygiene and Epidemiology as well as the Independent Ethics Committee of the Vietnam Ministry of Health (Approval Ref: 24/CN-HDDD dated 23 February, 2021) and authorized by the Vietnam Ministry of Health (Authorization reference: 1407/QD-BYT dated 26 February, 2021).

### Randomization and masking

Enrolled subjects were stratified by age (18–39 years or 40–59 years) and gender and randomly assigned in sequence to one of 5 groups (vaccine containing 1 μg SARS-CoV-2 spike (S) with or without 1.5 mg CpG1018 adjuvant, 3 μg S, 10 μg S, or saline placebo). Subjects were enrolled in 5 cohorts, each including active treatment and placebo groups, using a computer-generated randomization sequence prepared by an unblinded statistician; an unblinded pharmacist team dispensed each treatment according to the randomization sequence. The dose escalation steps are outlined in Figure 1. Briefly, for each formulation, a sentinel group was given the vaccine or placebo, followed by an eight-day monitoring period for reactogenicity and safety. Safety data from each sentinel group were reviewed by the Protocol Safety Review Team (PSRT) who cleared the administration of the vaccine to the remainder of the dose/formulation group (i.e., antigen/+/-adjuvant) as well as the administration of the next dose/formulation to the next sentinel group.

**Figure 1.**
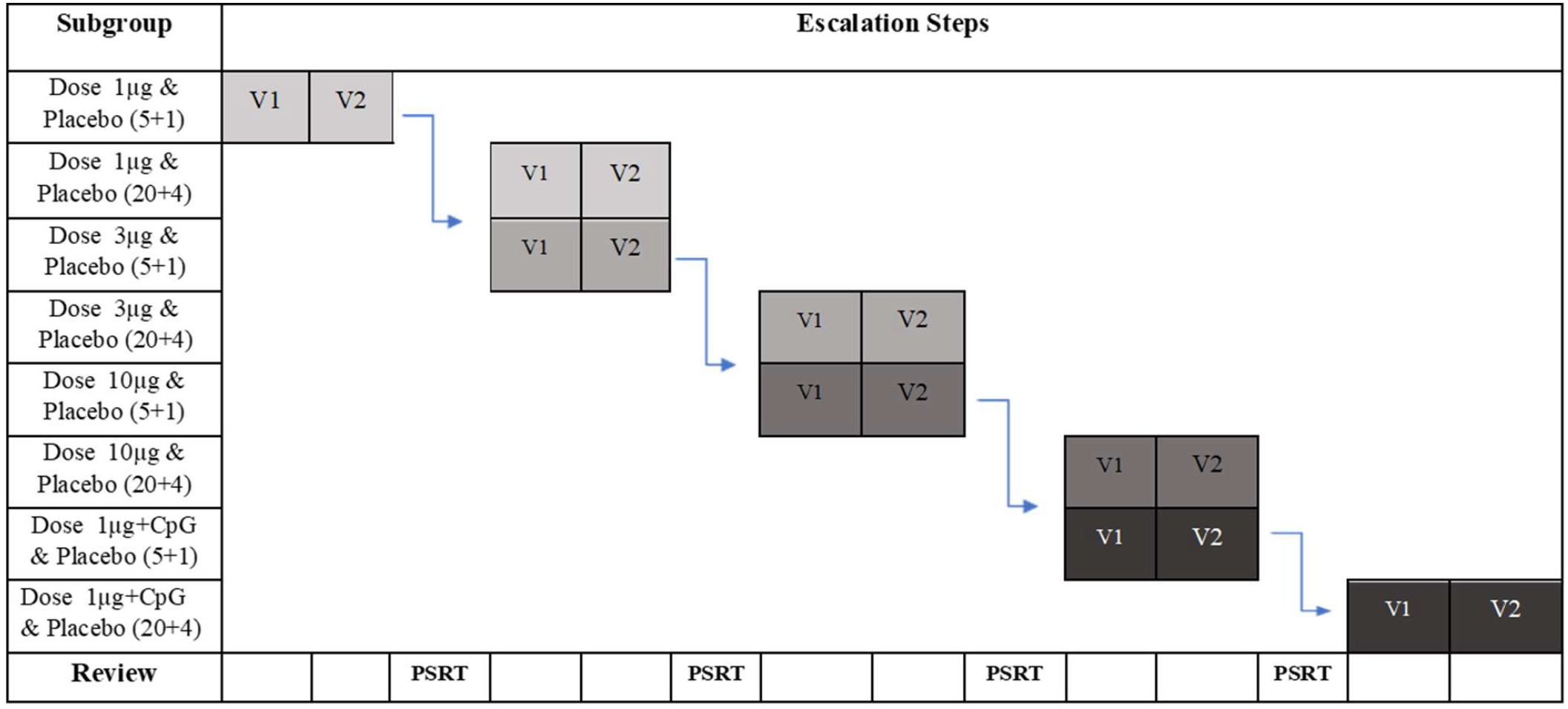
Randomization and dose escalation schedule of the phase 1 stage of the phase 1/2 randomized, placebo-controlled, observer-blind trial to assess the safety and immunogenicity of COVIVAC vaccine produced by IVAC in adults aged 18-75 years in vietnam. Subjects were randomized to one of five groups and were enrolled in five cohorts. Each cohort included active and placebo groups. Next cohort vaccination proceeded after a protocol safety review team PSRT safety review.

All participants and personnel other than the unmasked pharmacy team and vaccinators were masked to treatment.

### Investigational product

The recombinant NDV-HXP-S vaccine (COVIVAC) was manufactured according to current Good Manufacturing Practice by IVAC in their Influenza Vaccine Plant (Nha Trang, Vietnam) using locally procured embryonated eggs inoculated with a master virus seed made and extensively tested for adventitious agents by the Icahn School of Medicine at Mount Sinai (New York, United States). After incubation for 72 hours at 37°C, eggs were chilled overnight at 4°C, then the allantoic fluids were harvested, clarified, and concentrated. Recombinant virus particles were purified from the concentrated harvest by continuous-flow sucrose-gradient centrifugation, diafiltered against phosphate-buffered saline (PBS), inactivated by treatment with 1:4000 BPL for 24 hours at 4°C, and 0.2-micron filter-sterilized. Vaccine potency (i.e., amount of HXP-S antigen per dose) was measured by direct enzyme-linked immunosorbent assay (ELISA) using a human monoclonal antibody (CR3022) (8). which binds to the receptor binding domain on the SARS-CoV-2 spike glycoprotein S1 (LakePharma, Inc.) and an NDV-HXP-S standard that had been calibrated to a purified HXP-S reference (4) by sodium dodecyl sulphate polyacrylamide gel electrophoresis (SDS-PAGE) densitometry.

### Clinical procedures

Blinded staff administered study treatments by intramuscular injection of 0·5 mL on study days 1 and 29. Blood samples were drawn and clinical assessments were done for safety and immunogenicity endpoints before vaccination on days 1 (first dose), 8, 29 (second dose), 36, and 43; a clinical assessment for safety only on day 57 was the last time point considered for this interim analysis of the phase 1 cohort, although there will be additional immunogenicity and safety assessments on study day 197. Subjects were observed in the clinic for 30 minutes after each vaccination and were asked to record any adverse events (AEs) using paper diary cards during the 7 days after each vaccination.

Solicited injection site reactions (pain/tenderness, swelling/induration, erythema) and systemic symptoms (headache, fatigue, malaise, myalgia, arthralgia, nausea, vomiting, and fever defined as oral temperature ≥38°C) were recorded by subjects in a diary card for 7 days post vaccination that included intensity, which were then reported by the investigators. These events were not assessed for causality. Subjects also recorded and reported AEs for 28 days; the investigator included these in the study database after interviewing the subjects, grading them for intensity and categorizing them as serious or not. The investigators also identified the following AEs of special interest: potential immune-mediated medical conditions and AEs of special interest associated with COVID-19. Intensity of AEs was graded 1–4 as follows: 1 or mild (minimal interference with daily activities), 2 or moderate (interferes with, but does not prevent, daily activities), 3 or severe (prevents daily activities, intervention required), and 4 or potentially life-threatening (medical intervention required to prevent disability or death). Investigators assessed unsolicited AEs for causality (related to vaccination or not). AEs were graded according to US Department of Health and Human Services severity grading tables (Food and Drug Administration, Center for Biologics Evaluation and Research [September 2007] and National Institutes of Health, Division of AIDS [version 2.1, July 2017]). A PSRT regularly reviewed blinded safety data. A Data Safety Monitoring Board (DSMB) monitored unblinded safety data.

### Assessment of anti-S IgG binding and neutralization of SARS-CoV-2

Total anti-SARS-CoV-2 spike (S) IgG was measured using a validated indirect ELISA at Nexelis (Laval, Canada). Purified recombinant SARS-CoV-2 pre-fusion spike (Nexelis) at 1μg/ml in phosphate buffered saline (PBS, Wisent Bioproducts) was adsorbed to 96 well Nunc Maxisorb microplates (Thermo Fischer Scientific) and blocked with 5% skim milk in PBS, containing 0.05% Tween 20. Serial dilutions of test samples and the assay standard plus controls were added in the plates and incubated for 60 minutes at room temperature (15–30°C). After washing, horseradish peroxidase (HRP) enzyme-conjugated goat anti-human IgG-Fc (Jackson ImmunoResearch Laboratories) was added for 60 minutes at room temperature (15–30°C), then washed. Bound secondary antibody was reacted with 3,3′,5,5′-tetramethylbenzidine ELISA peroxidase substrate (Bio-Rad Laboratories) and incubated for 30 minutes at room temperature (15–30°C) before the reaction was stopped with 2N H_2_SO_4_. Plates were read at 450 nm with a correction at 620 nm to assess the level of anti-S IgG bound to the microtiter plate. A reference standard on each plate determined the quantity of anti-S IgG in arbitrary units (AU/mL). Concentrations were transformed to binding antibody units per mL (BAU/mL), based on the World Health Organization (WHO) International Standard for anti-SARS-CoV-2 immunoglobulin (9), using a conversion factor determined during assay validation (1/7.9815). The assay ‘s cut-off and lower limit of quantitation (LLOQ) was 6.3 BAU/mL.

Serum neutralizing activity against the Wuhan-Hu-1 strain of SARS-CoV-2 was measured in a validated pseudotyped virus neutralization assay (PNA) that assessed particle entry-inhibition (10). Briefly, pseudotyped virus particles containing a luciferase reporter for detection were made from a modified vesicular stomatitis virus (VSVΔG) backbone expressing the full-length spike glycoprotein of SARS-CoV-2 (MN908947, Wuhan-Hu-1) from which the last 19 amino acids of the cytoplasmic tail were removed (11). Seven two-fold serial dilutions of heat-inactivated serum samples were prepared in 96-well round-bottom transfer plates (Corning). Pseudotyped virus was added to the serum dilutions at a target working dilution (100,000 RLU/well) and incubated at 37°C with 5% CO_2_ for 60 ± 5 minutes. Serum-virus complexes were then transferred onto 96-well white flat-bottom plates (Corning), previously seeded overnight with Vero E6 cells (Nexelis) and incubated at 37°C and 5% CO_2_ for 20 ± 2 hours. Following this incubation, luciferase substrate from ONE GloTM Ex luciferase assay system (Promega) was added to the cells. Plates were then read on a SpectraMax^®^ i3x plate reader (Molecular Devices) to quantify relative luminescence units (RLU), inversely proportional to the level of neutralizing antibodies present in the serum. The neutralizing titre of a serum sample was calculated as the reciprocal serum dilution corresponding to the 50% neutralization antibody titre (NT50) for that sample; the NT50 titres were transformed to international units per mL (IU/mL), based on the WHO international standard for anti-SARS-CoV-2 immunoglobulin, using a conversion factor determined during assay validation (1/1.872). The assay ‘s cut-off and LLOQ were 5.3 IU/mL (10 as NT50) and 5.9 IU/mL, respectively.

To benchmark vaccine immunogenicity assessed in BAU/mL and IU/mL, group-level results were compared to a panel of human convalescent serum samples (HCS) collected 14 days after symptom onset from cases, consecutively collected, of mild to moderate COVID-19 illness among health care personnel seen as outpatients in Quebec, Canada during mid-2020.

Live virus neutralization by sera from vaccinees was also assessed as previously described (12) (13) (14). Vero E6 cells were seeded onto 96-well cell culture plates (20,000 cells/well) one day prior to the assay. Serum samples were heat-inactivated at 56 °C for 1 hour. Serial dilutions of sera were prepared in 1X minimal essential medium (MEM; Life Technologies) at a starting dilution of 1:10. Work with wild type (WT) SARS-CoV-2 (isolate USA-WA1/2020) and Delta variant (B.1.617.2) viruses was performed in a biosafety level 3 (BSL3) facility. For this, 1000 50% tissue culture infectious doses (TCID50s) /ml of virus were incubated with serially diluted sera for 1 h at room temperature. Media was removed from cell monolayers and 120 μl of virus-serum mix were added to the cells for 1 hour at 37°C. The virus-sera mix was removed and 100 μl of each corresponding serum dilution was added to every well. In addition, 100 μl of 1X MEM supplemented with 2% fetal bovine serum (FBS, Thermofisher) were added to every well. Plates were incubated for 48 hours at 37°C, then media was removed and cells were fixed at 4°C overnight with 150 μl of a 10% formaldehyde (Polysciences) solution. Cells were permeabilized and stained using the anti-nucleoprotein antibody 1C7C7 as previously described in detail (12) (13, 14). The 10 convalescent serum samples used in the live virus neutralization study were collected from participants in the longitudinal observational PARIS (Protection Associated with Rapid Immunity to SARS-CoV-2) study (13,15) This cohort follows health care workers longitudinally since April 2020. The study was reviewed and approved by the Mount Sinai Hospital Institutional Review Board (IRB-20-03374). All participants signed written consent forms prior to sample and data collection. All participants provided permission for sample banking and sharing.

### Outcome

The primary outcomes were frequency and intensity of solicited injection site and systemic AEs during the seven days after vaccination; frequency, intensity, and relatedness of clinically significant haematological and biochemical measurements at seven days after each vaccination; frequency, intensity, and relatedness of unsolicited AEs during 28 days after each vaccination; and occurrence of medically-attended AEs, serious AEs, and AEs of special interest during the interim analysis period of 57 days after-first vaccination. The secondary immunogenicity outcomes were anti-S IgG and NT50 against Wuhan-Hu-1 strain SARS-CoV-2 pseudotyped virus assessed on days 29 and 43 and expressed as geometric mean titer (GMT) or concentration (GMCs, BAU/mL for ELISA, or IU/mL for PNA), geometric mean fold rise (GMFR) from baseline, and percentage of subjects with ≥ 4-fold increase and ≥ 10-fold increase from baseline. Live virus neutralization assay (VNA) was also performed on a subset of day 43 samples, expressed as NT50 GMT for the Wuhan (isolate USA-WA1/2020) and B.1.617.2 (Delta) strains of SARS-CoV-2.

### Statistical Analyses

In this phase 1 study (ClinicalTrial.gov NCT04830800) 120 subjects were randomized in 8 groups to allow for the appropriate dose escalation and/or introduction of an adjuvanted formulation (**Figure 1**) resulting in 25 subjects per candidate vaccine formulation and 20 subjects assigned to the placebo. All safety assessments took place in the treatment-exposed population, according to the treatment received. All group-level percentages were supplemented with two-sided 95% confidence intervals (CIs) computed via the Clopper-Pearson method. The analysis of immunogenicity was performed in the per protocol population, which excludes subjects with protocol deviations that would affect the immunogenicity assessment. Immunogenicity data were descriptively analysed. Geometric mean antibody responses were reported by treatment and time point, accompanied by 95% CIs. The analysis of geometric means excluded subjects who were seropositive at baseline (defined by anti-S IgG >LLOQ as measured by ELISA). GMFRs were calculated relative to baseline using the log difference of the paired samples, with corresponding CIs computed via the *t*-distribution, utilizing the antilog transformation to present the ratio. The proportions of subjects with GMFRs of NT_50_ ≥ 4 and ≥ 10 from baseline were summarized with 95% CIs. The analysis of immunogenicity relative to baseline included five subjects who were seropositive at baseline, three of which were in the placebo arm. All statistical analyses were performed by an independent statistician using SAS version 9.4.

## 3. Results

### Trial Attributes

Between March 15 and April 17, 2021, 120 healthy adults were enrolled and assigned to one of five treatment groups as shown in **Figure 2**. All subjects received two doses of vaccine or placebo. The baseline characteristics are shown by treatment group in **Table 1**. The exposed population was 48.3% female, had a median age of 38 years (IQR 23, 43.5), and a median body mass index of 22.44 (IQR 20.97-24.1).

**Figure 2.**
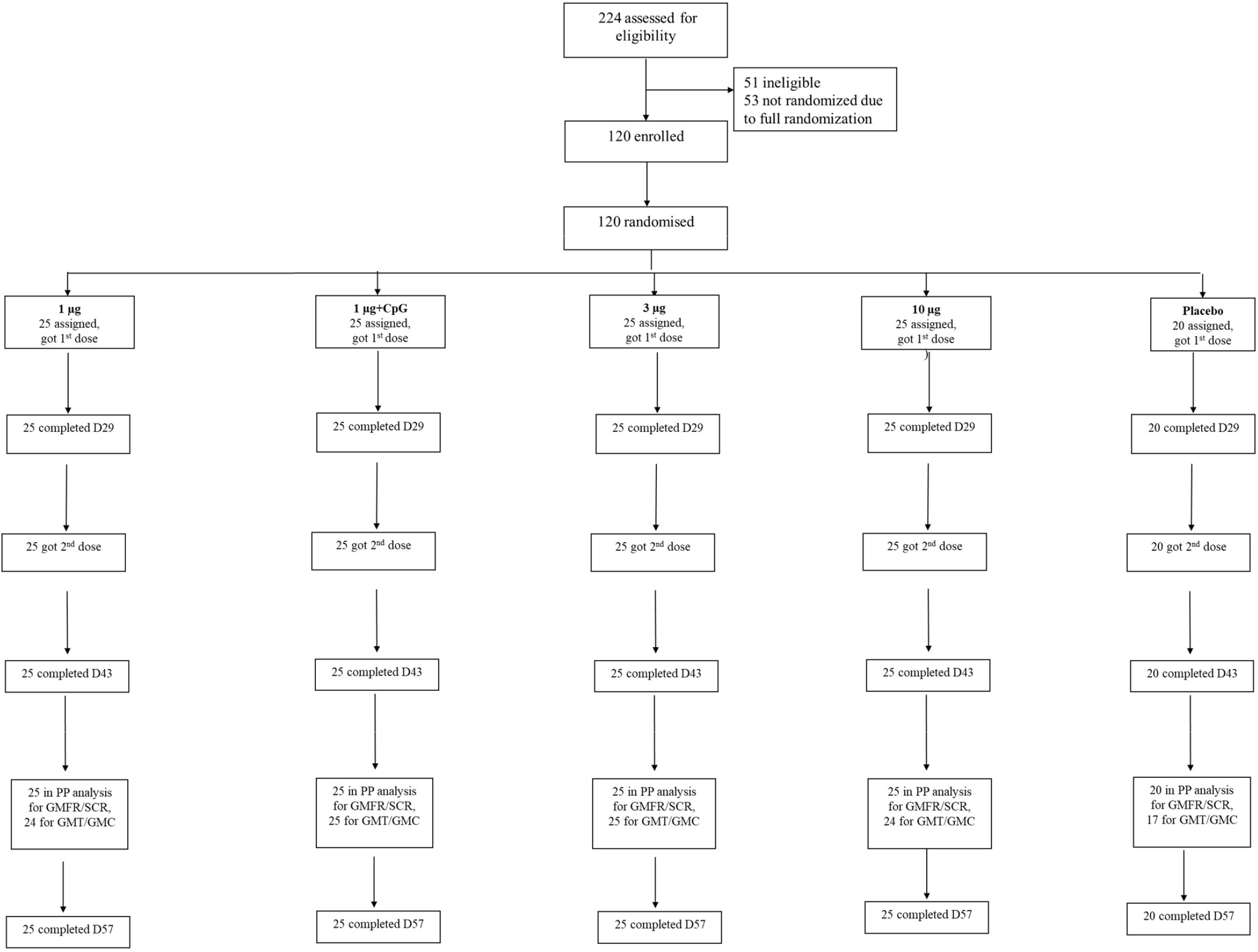
Profile of the phase 1 stage of the phase 1/2 randomized, placebo-controlled, observer-blind trial to assess the safety and immunogenicity of COVIVAC vaccine produced by IVAC in adults aged 18-75 years in vietnam.

**Table 1.** Baseline characteristics of the exposed population of the phase 1 stage of the COVIVAC phase 1/2 study. Data are median (quartile 1-quartile 3) or n (%).

|  | 1 µg<br>(N = 25) | 3 µg<br>(N = 25) | 10 µg<br>(N = 25) | 1 µg<br>+CpG<br>(N = 25) | Placebo<br>(N = 20) |
| --- | --- | --- | --- | --- | --- |
| Age, years | 30.0<br>(24.0-44.0) | 39.0<br>(25.0-45.0) | 40.0<br>(23.0-43.0) | 40.0<br>(22.0-44.0) | 29.0 (23.0-42.5) |
| Sex |  |  |  |  |  |
| Male | 13<br>(52.0%) | 14<br>(56.0%) | 13<br>(52.0%) | 12<br>(48.0%) | 10 (50.0%) |
| Female | 12<br>(48.0%) | 11<br>(44.0%) | 12<br>(48.0%) | 13<br>(52.0%) | 10 (50.0%) |
| Body mass index | 21.66<br>(20.44-23.06) | 22.02<br>(21.43-24.57) | 22.39<br>(21.13-24.71) | 22.53<br>(20.88-23.73) | 22.68 (20.80-23.77) |

### Safety

All four formulations of NDV-HXP-S were well tolerated with no dose-limiting reactogenicity (**Table 2**). Most solicited injection site and systemic reactogenicity during 7 days after each vaccination was mild and transient with no apparent difference between doses 1 and 2. The most common injection site symptoms (**Table 2**) were pain or tenderness; these were most frequent at the highest dose. The most common systemic symptoms (**Table 2**) were fatigue or malaise, headache, and myalgia, all generally in less than one-third of subjects. Fever was uncommon. AEs occurring during 28 days after vaccination (**Table 3**) and judged by the investigator to be treatment-related were infrequent (< 5%). No treatment-related serious adverse event occurred, nor did any AE of special interest reported during the 57-day assessment period. Haematology and serum chemistry laboratory readouts were assessed on day 8 following each vaccination; no clinically notable finding relative to baseline assessment were detected. The independent DSMB expressed no safety concerns with the study proceeding to the phase 2 stage of the study.

**Table 2.**
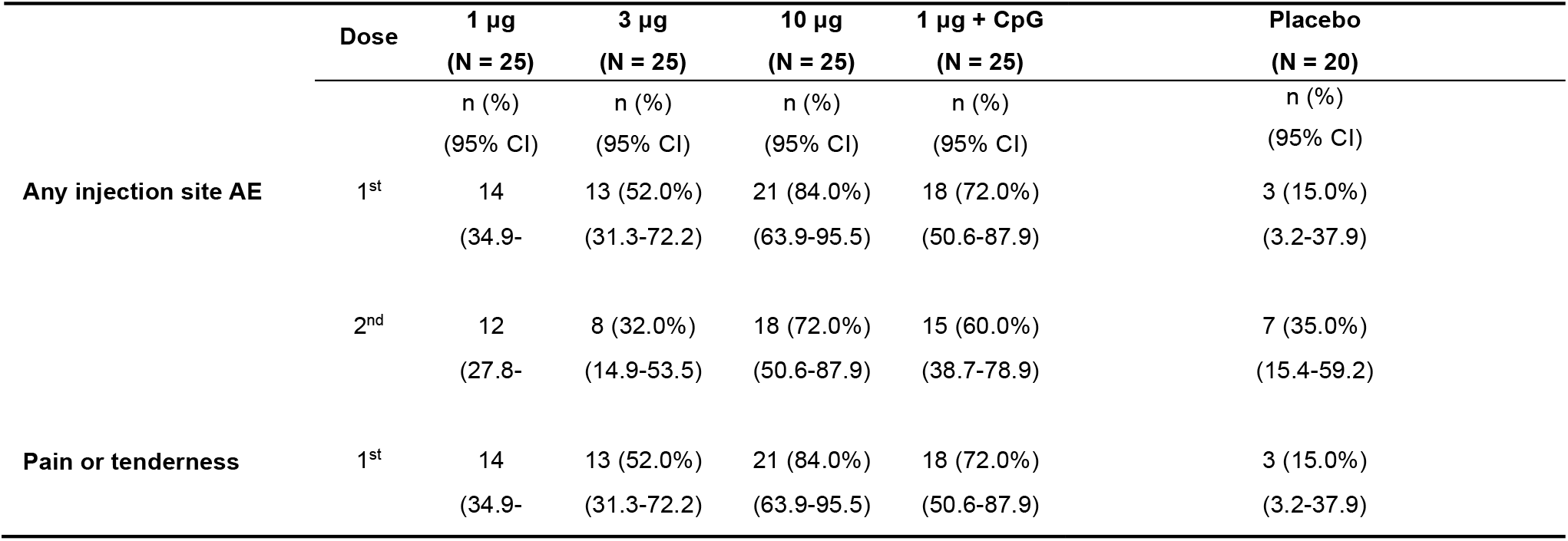
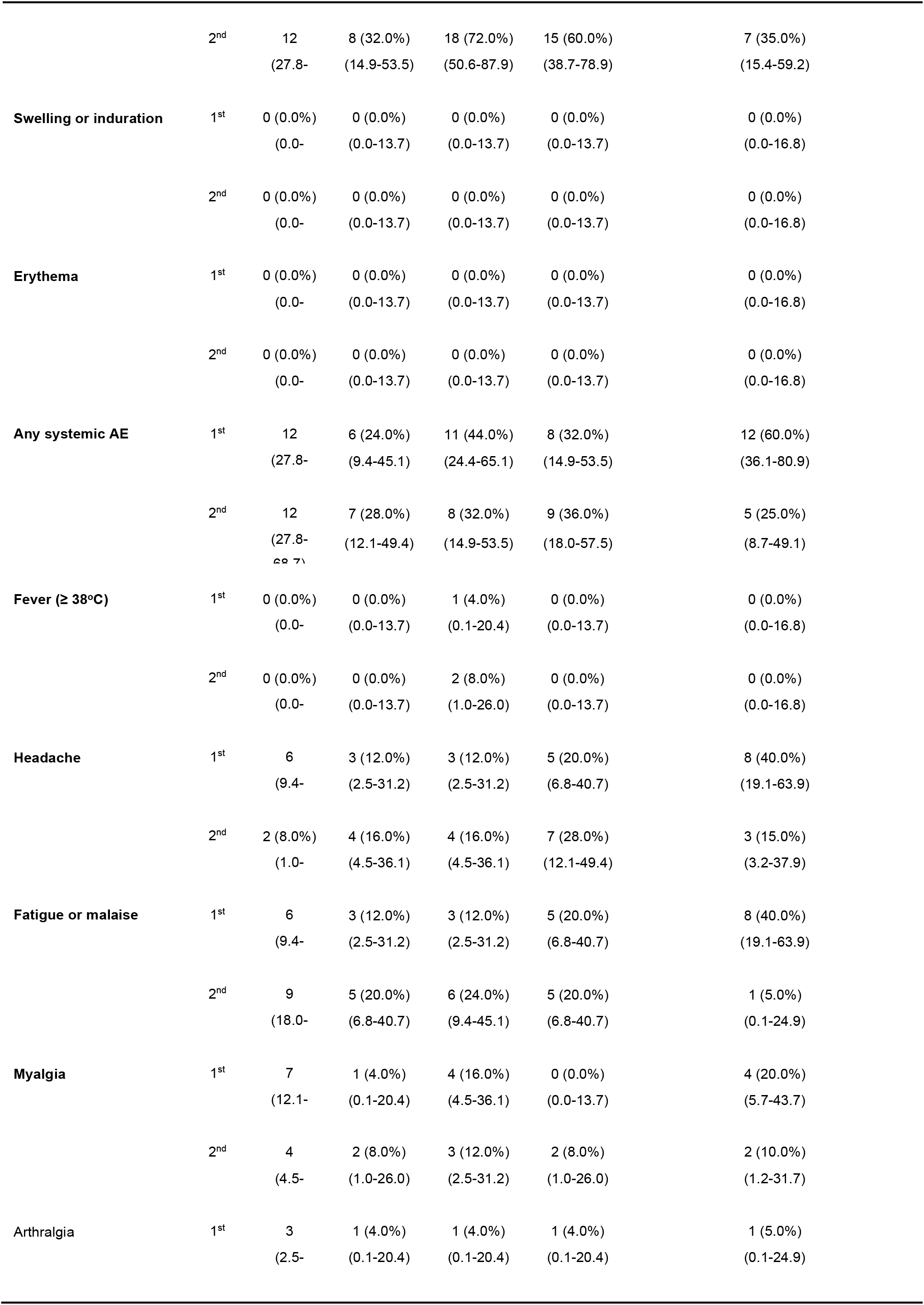
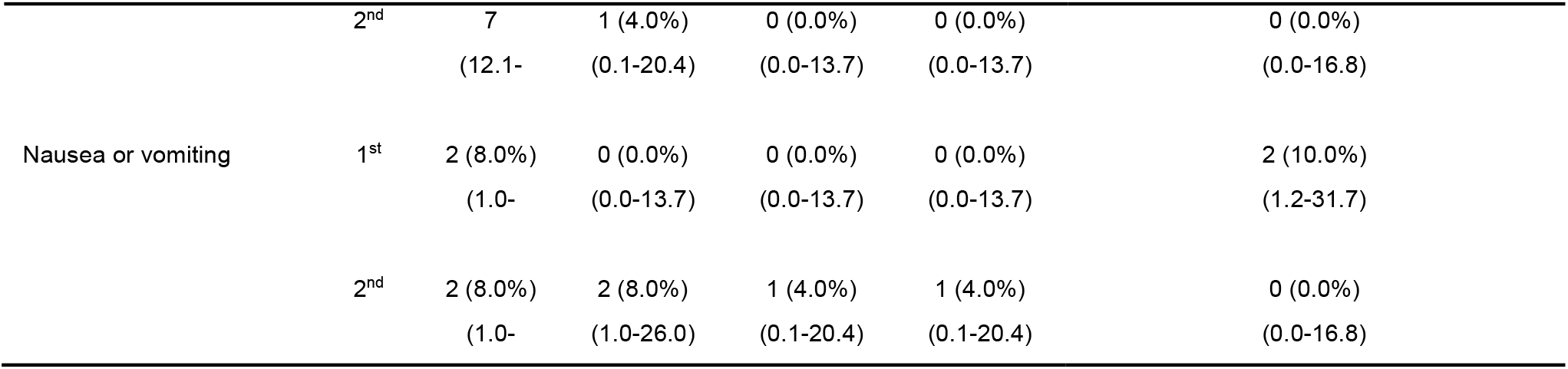
Solicited adverse events (AEs) during 7 days after vaccination with NDV-HXP-S or placebo in the phase 1 stage of the COVIVAC phase 1/2 study. Two-sided 95% confidence intervals (CIs) computed via the Clopper-Pearson method comparing overall dose levels.

**Table 3.** Adverse events (AEs) with onset during 28 days after vaccination with NDV-HXP-S or placebo in the phase 1 stage of the COVIVAC phase 1/2 study, by treatment groups. Two-sided 95% confidence intervals (CIs) computed via the Clopper-Pearson method.

|  | 1 µg<br>(N = 25) | 3 µg<br>(N = 25) | 10 µg<br>(N = 25) | 1 µg +<br>CpG<br>(N = 25) | Placebo<br>(N=20) |
| --- | --- | --- | --- | --- | --- |
|  | n (%)<br>(95% CI) | n (%)<br>(95% CI) | n (%)<br>(95% CI) | n (%)<br>(95% CI) | n (%)<br>(95% CI) |
| **With one or more AE** |  |  |  |  |  |
| Dose 1 | 10<br>(21.1- | 5<br>(6.8- | 8<br>(14.9- | 8<br>(14.9- | 7 (35.0%)<br>(15.4- |
| Dose 2 | 3<br>(2.5- | 6<br>(9.4- | 6<br>(9.4- | 6<br>(9.4- | 5<br>(8.7- |
| **Any vaccine-related** |  |  |  |  |  |
| Dose 1 | 2 (8.0%)<br>(1.0- | 2 (8.0%)<br>(1.0- | 1 (4.0%)<br>(0.1- | 0 (0.0%)<br>(0.0- | 1 (5.0%)<br>(0.1- |
| Dose 2 | 0 (0.0%)<br>(0.0- | 0 (0.0%)<br>(0.0- | 0 (0.0%)<br>(0.0- | 1 (4.0%)<br>(0.1- | 0 (0.0%)<br>(0.0- |
| **Serious** |  |  |  |  |  |
| Dose 1 | 0 (0.0%)<br>(0.0- | 0 (0.0%)<br>(0.0- | 0 (0.0%)<br>(0.0- | 0 (0.0%)<br>(0.0- | 0 (0.0%)<br>(0.0- |
| Dose 2 | 0 (0.0%)<br>(0.0- | 0 (0.0%)<br>(0.0- | 0 (0.0%)<br>(0.0- | 0 (0.0%)<br>(0.0- | 1 (5.0%)<br>(0.1- |
| **Serious vaccine-** |  |  |  |  |  |
| 1<sup>st</sup> Dose | 0 (0.0%)<br>(0.0- | 0 (0.0%)<br>(0.0- | 0 (0.0%)<br>(0.0- | 0 (0.0%)<br>(0.0- | 0 (0.0%)<br>(0.0- |
| 2<sup>nd</sup> Dose | 0 (0.0%)<br>(0.0- | 0 (0.0%)<br>(0.0- | 0 (0.0%)<br>(0.0- | 0 (0.0%)<br>(0.0- | 0 (0.0%)<br>(0.0- |

### Immunogenicity

Two doses of NDV-HXP-S were immunogenic in a formulation and dose dependent manner within the per protocol population. Five subjects, who were seropositive prior to the administration of the first dose (three in the placebo group, one in the 1 μg group and one in the 10 μg group), were excluded from the per protocol analysis. Induction of anti-S IgG was modest following dose one but a marked anamnestic response was observed 14 days after vaccine dose two (**Figure 3A**). Seronegative individuals in the vaccine groups responded 28 days after first vaccination with GMCs of anti-S IgG between 9.89 (1 μg) and 29.33 (10 μg) BAU/mL (**Figure 3 A**), with a ≥ 4-fold increase in 44–84%. The second dose considerably increased anti-S-IgG antibody responses after 14 days to GMCs between 122.54 (1 μg) and 446.5 (10 μg) BAU/mL (**Figure 3A, Table 4**). All individuals in the 3, 10, and 1 μg + CpG vaccine groups had a ≥ 4-fold increase over baseline after the second dose (**Figure 3C, Table 5**). Ninety-six percent (96%) of individuals in the 1 μg vaccine group also had a ≥ 4-fold increase over baseline after the second dose (**Figure 3C, Table 5**). All individuals in the 3 μg group had a ≥ 10-fold increase, as did > 90% of vaccinees in the other three vaccine groups (**Figure 3C**). In this study, the adjuvant effect of CpG 1018 was limited after two vaccine doses (**Table 4**) of the 1 μg dose (the only adjuvanted dose). The non-adjuvanted 1 μg group had a GMC of 122.54 BAU/mL (95% CI 87.06-172.48) while the 1 μg+CpG1018 group had a GMC of 206.51 BAU/mL (95% CI 152.89-278.93), representing a fold increase of 1.69 (95% CI: 1.08-2.62). GMCs of anti-S IgG among the vaccine groups on day 43 exceeded the GMC of the HCS panel (N=29, 72·93 95% CI 33.00-161.14) by 1.7-6.1 (**Table 4**).

**Figure 3.**
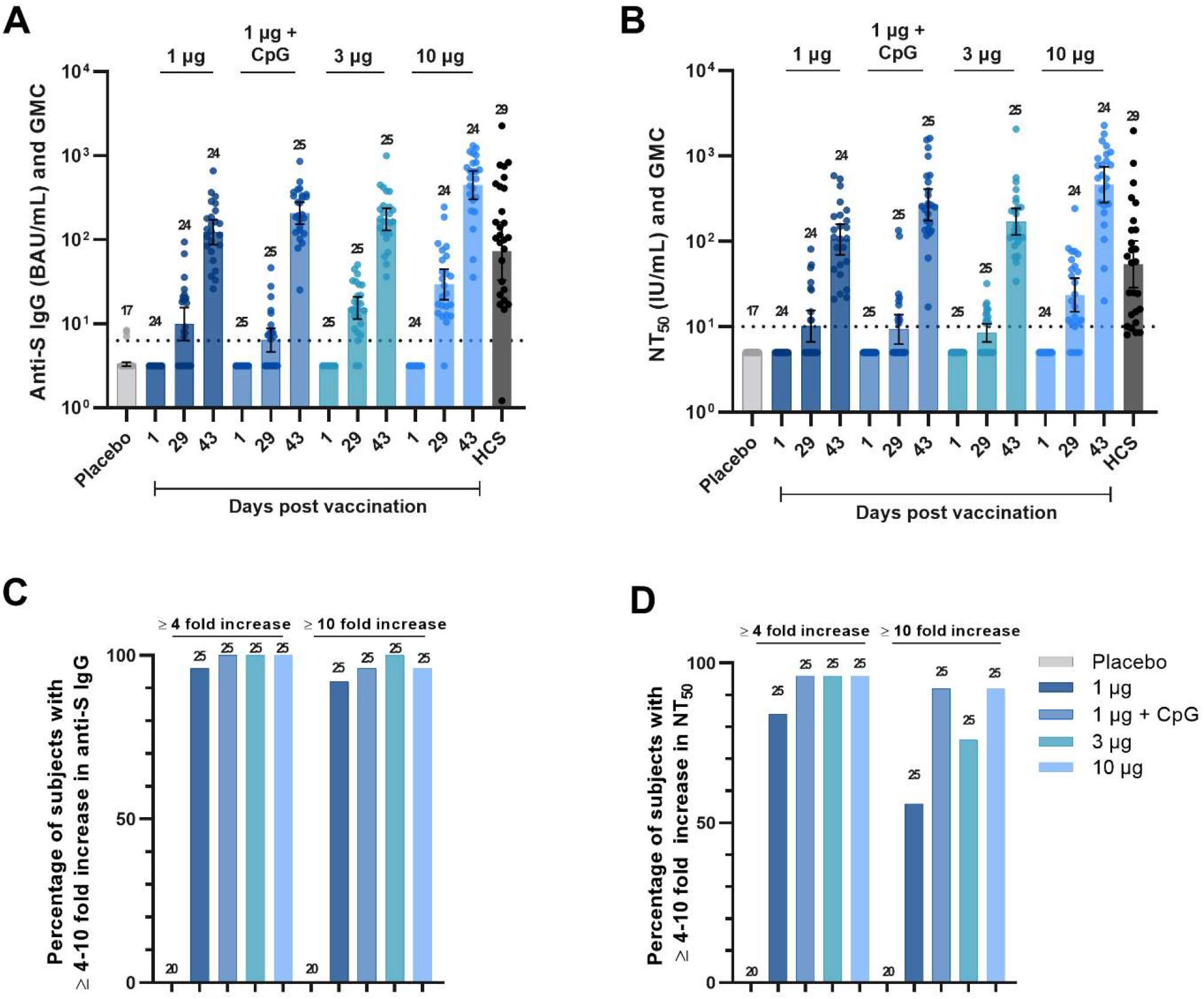
Distribution and geometric mean concentration (GMC) of anti-S IgG (BAU/mL) in placebo, vaccine groups and human convalescent sera (HCS) controls (A), distribution and GMC of NT50 by pseudoneutralization assay (PNA) (IU/mL) in placebo, vaccine groups, and HCS controls (B), percentage of subjects with ≥ 4–10-fold increase in anti-S IgG at day 43 (C), and percentage of subjects with ≥ 4–10-fold increase in NT50 by PNA at day 43 (D). Numbers above columns denote number of per-protocol subjects contributing data.

**Table 4.** Geometric mean concentration (GMC) of anti-S IgG (BAU/mL) and NT50 by pseudoneutralization assay (PNA) (IU/mL) post two doses of COVIVAC (Day 43) and GMC ratios, vaccine to human convalescent sera (HCS) panel. Subjects that were seropositive at baseline were removed from this analysis (one for the 1 μg and 10 μg groups and three for the placebo group).

| | 1 μg<br>(N = 24) | 3 μg<br>(N = 25) | 10 μg<br>(N = 24) | 1 μg + CpG<br>(N = 25) | Placebo<br>(N = 17) |
| --- | --- | --- | --- | --- | --- |
| Anti-S IgG BAU/mL | 122.54 | 173.38 | 446.50 | 206.51 | 3.15 |
| 95% CI | (87.06 , 172.48) | (128.12 , 234.61) | (302.60 , 658.83) | (152.89 , 278.93) | ( - ) |
| GMC ratio, vaccine to HCS panel | 1.68 (0.72, 3.94) | 2.38 (1.03, 5.50) | 6.12 (2.57, 14.61) | 2.83 (1.22, 6.55) |  |

| 95% CI |  |  |  |  |  |
| --- | --- | --- | --- | --- | --- |
| NT50 by PNA | 56.07 | 90.94 | 246.19 | 143.33 | 2.67 |
| 95% CI | (37.01 , 84.94) | (63.63 , 129.98) | (151.97 , 398.82) | (94.01 , 218.52) | ( - ) |
| GMC ratio, vaccine to HCS panel | 1.54 (0.74, 3.22) | 2.51 (1.23, 5.08) | 6.78 (3.13, 14.68) | 3.95 (1.89, 8.27) |  |
| 95% CI |  |  |  |  |  |

Functional antibody responses were assessed by PNA. Low NT50 GMCs were detected in all vaccine groups after the first vaccination (between 5.7 IU/mL and 12.55 IU/mL, **Figure 3B**) with ≥ 4-fold rises in 13% to 40% of the vaccine groups. The second vaccine dose strongly boosted neutralization GMCs (**Figure 3B**) to between 56.07 IU/mL (1 μg, 95% CI 37.01-84.94) and 246.19 IU/mL (10 μg, 95% CI 151.97-398.82), with a ≥ 4-fold increase over baseline in 84% to 96% of vaccine groups (**Table 5**) and a ≥10-fold rise in most individuals (92%) in the 10 μg and 1 μg+CpG1018 groups (**Figure 3D**). A ≥ 10-fold rise was observed in 56% and 76% of individuals in the 1 and 3 μg groups, respectively. Similar to the observation on the effect of adjuvant on levels of binding antibodies, the differences in post-second dose GMCs between the unadjuvanted and adjuvanted 1 μg and 1 μg+CpG1018 groups were limited: 1 μg, 56.07 IU/mL (95% CI 37.01-84.94) versus 1 μg+CpG1018, 143.33 IU/mL (95% CI 94.01 - 218.52), representing a representing a fold increase of 2.56 (95% CI, 1.4 - 4.6).

**Table 5.** Percentage of subjects with a ≥ 4-fold rise from baseline post two doses of COVIVAC (day 43) for anti-S IgG and NT50 by pseudoneutralization assay (PNA). Two-sided 95% CIs computed via the Clopper-Pearson method comparing overall dose levels.

| | 1 μg<br>(N = 25) | 3 μg<br>(N = 25) | 10 μg<br>(N = 25) | 1 μg +<br>CpG | Placebo<br>(N = 20) |
| --- | --- | --- | --- | --- | --- |
| Anti-S IgG n (%) $\geq 4$-fold rise from baseline | 24 (96.0%) | 25 (100%) | 25 (100%) | 25 (100%) | 0 (0.0%) |
| 95% CI | (79.6-99.9) | (86.3-100) | (86.3-100) | (86.3-100) | (0.0-16.8) |
| NT50 by PNA n (%) $\geq 4$-fold rise from baseline | 21 (84.0%) | 24 (96.0%) | 24 (96.0%) | 24 (96.0%) | 0 (0.0%) |
| 95% CI | (63.9-95.5) | (79.6-99.9) | (79.6-99.9) | (79.6-99.9) | (0.0-16.8) |

Based on the vaccine-homologous binding and neutralizing antibody responses, a clear ranking of immunogenicity for the unadjuvanted formulations was apparent, with the 10 μg formulation performing best followed by the 3 μg and 1 μg formulations. The 1 μg+CpG1018 ranked between the 10 and 3 μg. The induction of humoral immunity was strong with post-second dose GMFRs relative to baseline of 34.65-fold (1 μg) to 124.11-fold (10 μg) for anti-S IgG and 20.50-fold (1 μg) to 84.75-fold (10 μg) for NT50 antibodies (**Figure 4**). GMCs of NT50 by PNA among the vaccine groups on day 43 exceeded the GMC of the HCS panel (N=32, 36.30 95% CI 19·43-67.79) by 1.5-6.8-fold depending on the vaccine formulation (**Table 4**). The neutralization titers at day 43 highly correlated with anti-S-protein specific binding IgG (Figure 5, r=0.94).

**Figure 4.**
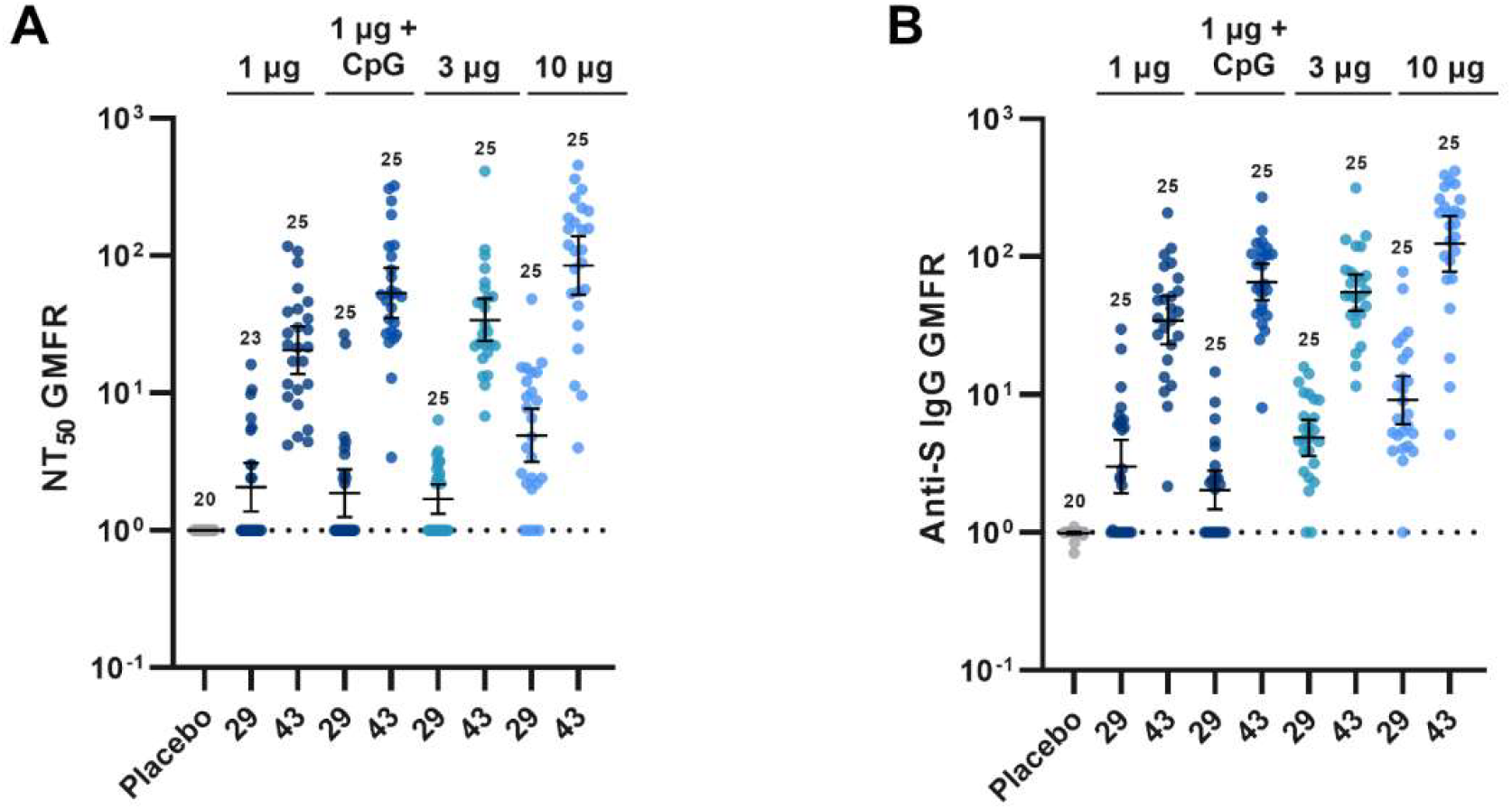
Distribution and geometric mean fold rise (GMFR) in anti-S IgG from baseline (A), distribution and GMFR of fold rise in NT50 by pseudoneutralization assay (PNA) from baseline (B). Numbers above data denote number of per-protocol subjects contributing data.

**Figure 5.**
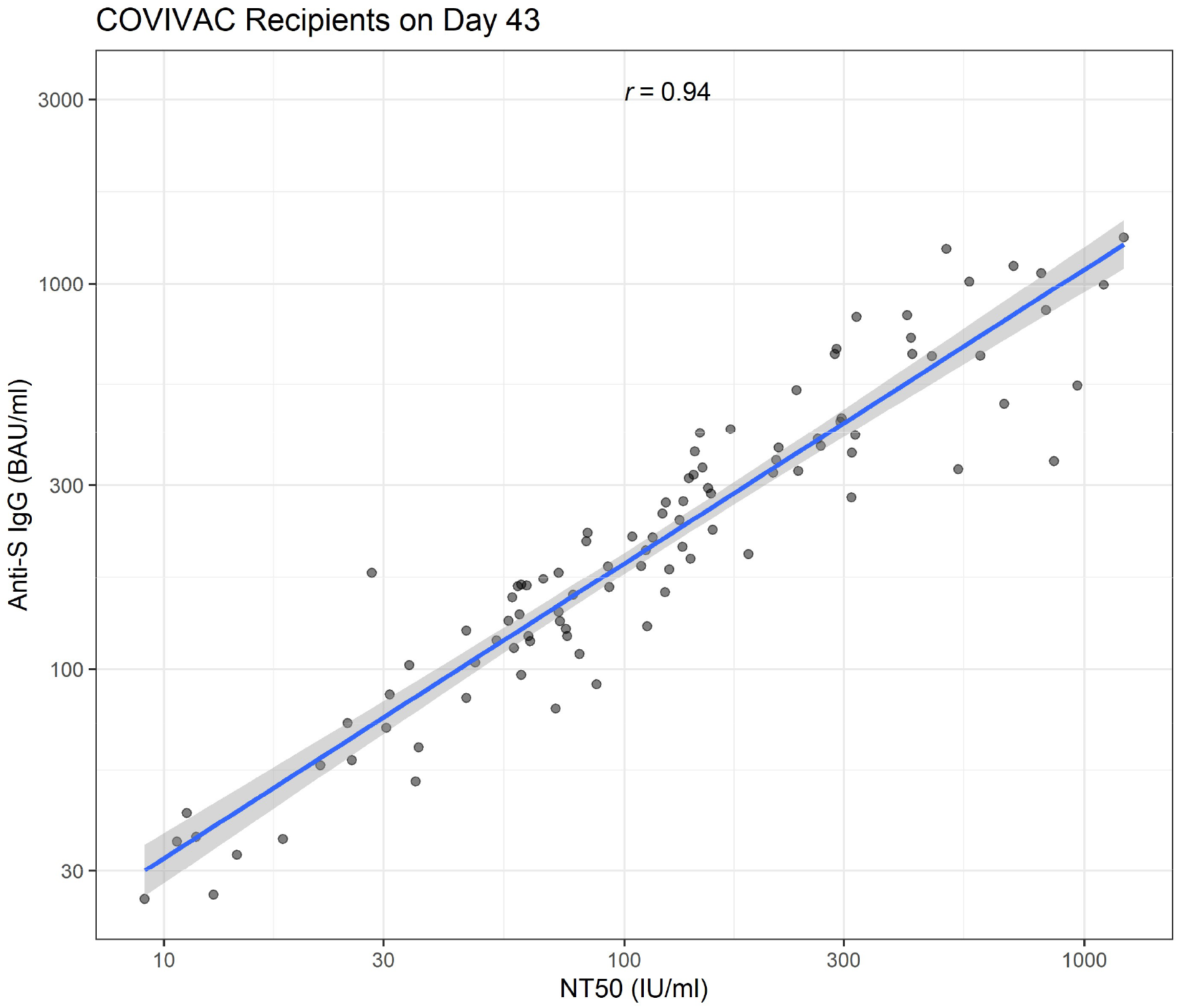
Scatterplot of anti-S IgG (BAU/ml) and NT50 (IU/ml) by pseudoneutralization assay (PNA) (Wuhan-Hu-1 spike) among all COVIVAC recipients on day 43 (14-days post second dose) of phase 1 clinical trial. The blue line provides a fitted linear line on the log scale and the Pearson correlation coefficient estimate is provided.

Additionally, neutralization of the wild type (USA-WA1/2020) and Delta (B.1.617.2) variant viruses was assessed by live virus neutralization of sera from placebo, vaccinees, and HCS controls (**Figure 6**). Reduction in neutralizing potency, relative to anti-wild type neutralising potency, was dose and adjuvant dependent. The 1 μg without or with CpG 1018 group showed a 1.6- and 1.9-fold reduction relative to the Delta variant respectively. The 3 μg group showed a 1.4-fold reduction relative to the Delta variant respectively. The 10 μg group showed a 1.6-fold reduction relative to the Delta variant respectively. No fold reduction for HCS was observed.

**Figure 6.**
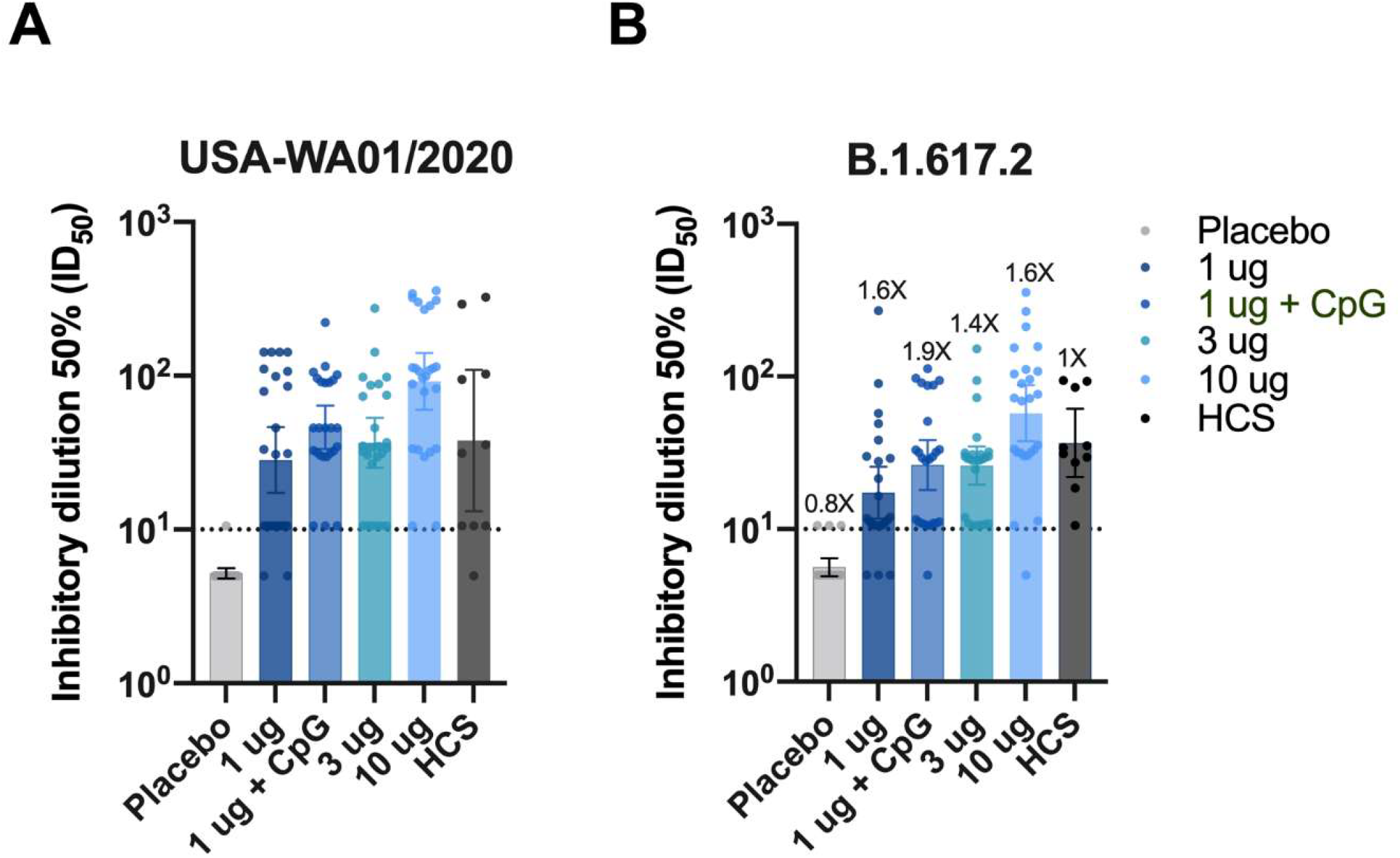
Neutralization of wild type SARS-CoV-2 and B.1.617.2 by vaccinees ‘ sera. Distribution of serum inhibitory dilution 50% (ID50) of sera from placebo, vaccine groups and human convalescent sera (HCS)controls against wild type SARS-CoV-2 USA-WA1/2020 isolate (A) and B.1.617.2 variant (B). Geometric mean titers (GMT) with 95% confidence interval (CI) is shown.

## 4. Discussion

Current production capacity cannot satisfy the global demand for COVID-19 vaccines and vaccine distribution is inequitable with most vaccines acquired and used by high-income countries while LMICs have limited access (1). Furthermore, vaccines requiring specialized cold supply chain and very low temperature storage, such as mRNA COVID-19 vaccines, may not be well suited for use in many LMICs in the near future. Thus, local production of COVID-19 vaccines in LMICs, compatible with prolonged 2–8°C storage in LMICs, preferably using existing manufacturing infrastructure and know-how, would increase global availability and reduce dependence of countries producing these vaccines on international vaccine supply. Here we strengthen the evidence (7) that an engineered inactivated NDV-based vaccine (COVIVAC) expressing a 6-proline stabilized SARS-CoV-2 spike protein (3-5), produced in eggs in an existing influenza virus vaccine production facility at IVAC in Vietnam, shows an acceptable reactogenicity and safety profile in humans and has immunogenicity that suggests its potential clinical benefit. We evaluated a range of vaccine doses (1 μg, 3 μg, 10 μg) having potency quantified as μg of virus envelope-anchored SARS-CoV-2 spike protein. Only the low dose (1 μg) was evaluated in a formulation with and without the TLR-9 agonist CpG1018 as a vaccine adjuvant. The decision to test only the lowest dose with an adjuvant was a result of a mandate by the local health authorities not to exceed 120 subjects in a first-in-human phase 1 clinical study and the need to maintain sufficient group size for the other formulations. Over 28 days after each vaccine dose, all formulations were very well tolerated with little solicited reactogenicity aside from mild injection site pain or tenderness. No clinically important treatment-related AE occurred during the 56 days of observation following first vaccination with any formulations. Moreover, the vaccine was strongly immunogenic in a formulation and dose-dependent manner, inducing levels of vaccine-homologous anti-S IgG and virus-neutralizing antibodies that exceeded by several fold the levels measured in 14-day convalescent sera from cases of health care workers with mild to moderate COVID-19 illness in 2020.

The adjuvant benefit, as measured by enhanced induction of humoral immunity was low, but the sample size was small and the availability of data from only one dose level with adjuvant limited the precision and breadth of the analysis. On the other hand, the vaccine elicited neutralizing antibodies against the Delta variant (B.1.617.2). While neutralizing antibody titres decreased modestly against B.1.617.2, this was expected and in the range observed with sera from recipients of the mRNA vaccines BNT162b2 and mRNA-1273 (16) These data are in line with what was observed in a phase 1/2 study in Thailand with a similar vaccine, manufactured from the same virus seed lot (7). Evidence to the neutralization by NDV-HXP-S of the now prevalent Delta (B.1.617.2) variant may indicate clinical benefit. Finally, we have shown a robust correlation between levels of binding IgG antibodies and neutralizing activity, expressed as NT50. Neutralizing activity has been suggested as a surrogate endpoint for clinical efficacy of COVID-19 vaccines (17-18). Given that the assessment of binding antibodies by ELISA is more reproducible and affordable than neutralizing assays, this correlation may support using ELISA for future COVID-19 vaccine development (e.g., dose selection or lot-to-lot consistency).

The study has several limitations. The sample size per treatment group was small, limiting precision. Also, assessments were restricted to 43 days for immunogenicity and 57 days for reactogenicity and safety, narrowing our perspective to acute outcomes only. These are inherent problems of phase 1 trials and interim analyses in a pandemic response setting. Nevertheless, since clinical trials with similar vaccines are underway in Thailand (NCT04764422) and Brazil (NCT04993209), we determined that publication of early data is a priority, with a follow-on publication of the results of the full study. One additional weakness is the absence of Omicron (B.1.1.529) neutralization data, which was not generated for this initial study.

The study had strengths as well. The vaccine construct is a novel platform expressing a second-generation prefusion-stabilized S protein in a membrane-bound trimeric conformation. We hypothesize that these characteristics contribute to the vaccine ‘s immunogenicity, even without the CpG1018 adjuvant. The anti-S ELISA and PNA used to assess vaccine-homologous NT50 potency were validated and results are expressed in international units (9) for future comparisons. The induction of anti-S binding and neutralizing antibodies was contrasted with mean levels in human convalescent serum and found to be superior, especially in the mid- and high-dose groups. Furthermore, we have shown with a live neutralization assay that the vaccine candidate elicits neutralizing activity against the Delta variant of concern. The neutralizing capacity of NDV-HXP-S vaccine will be further assessed in the phase 2 stage of this study, using the most relevant variants of concern.

Originally, this study was designed as a phase 1/2 study with two-part selection design with elimination of two candidate groups after the first part (i.e., phase 1). The study was designed to have greater than 90% power to identify the candidate with the highest response as measured by the NT_50_ by ranked GMCs, assuming the true GMC is at least 1.5-fold larger than the second highest candidate group and to provide a preliminary safety evaluation of the candidates. After the phase 1 interim analysis, however, proceeding in development with an independent active-controlled phase 2 was deemed more appropriate. Thus, after the interim analysis, one candidate was selected to advance, in addition to a new dose form (6 μg S), as well as an active comparator (AZD1222), at which time 375 additional subjects were randomized 1:1:1 to the two candidate groups and the active control, respectively.

In summary, we show that the inactivated NDV-HXP-S vaccine candidate (COVIVAC) has an acceptable safety profile and is highly immunogenic. The technology for this vaccine can be rapidly transferred to and produced at low cost in any facility designed for production of IIV; such facilities are present in a number of LMICs (2). On the basis of these results and acknowledging the need to balance output of vaccine doses from the manufacturing facility with a robust immune response, the phase 2 stage of the ongoing clinical trial uses the 3 μg unadjuvanted dose together with a 6 μg dose formulation for further assessment in comparison to AZD1222, which has been authorized for use in Vietnam.

## Supporting information

IVAC NDX-HXP-S Phase 1 protocol

## Data Availability

The study protocol is provided in the appendix. Individual participant data will be made available when the trial is complete, upon request directed to Assoc. Prof. Thiem Dinh Vu. After approval of a proposal, data can be shared through a secure online platform.

## Author Contributions

All authors have read and agreed to the published version of the manuscript. Individual author roles are reported using CRediT: Conceptualization, AGS, PP, FK, LDM RS, BLI ; methodology, LDM, RR, ML, JAW, RH ; software, ; validation, SL, PC, JMC, LDM; formal analysis, SL, JMC, JT, LDM ; investigation, ADD, TDV, VTT, ATP, HHV, MTD, WS, JMC, JT, AR, IGD, SS, AR, JAW; resources, ADD, TDV, VTT, HHV, ATP, MTD, BVL, THD, DVN, WS,RR, ML, JAW, BLI, RS ; data curation, SL, PC; writing—original draft preparation, BLI, RS ; writing—review and editing, SL, PC, JSM, CH, AGS, PP, FK, TCT, LDM, RR, JAW, RH, BLI ; visualization, JT, JMC. LDM; supervision, BVL, THD, AGS, PP, FK, HMV, TCT, HMN, ML, JAW ; project administration, ADD, TDV, VTT, ATP, RS ; funding acquisition, THD, BLI.

## Acknowledgments

Activities at PATH and Mount Sinai were supported, in part, by the Bill & Melinda Gates Foundation (INV-021239). PATH was also supported by the Coalition for Epidemic Preparedness (CEPI) and Bayer AG. Under the grant conditions of the Bill & Melinda Gates Foundation, a Creative Commons Attribution 4.0 Generic License has already been assigned to the Author Accepted Manuscript version that might arise from this submission. The findings and conclusions contained within this manuscript are those of the authors and do not necessarily reflect positions or policies of the Bill & Melinda Gates Foundation. The salary of PP was partially funded by NIH (Centers of Excellence for influenza Research and Response, 75N93021C00014), U.S. NIAID grant (P01 AI097092-07), U.S. NIAID grant (R01 AI145870-03), by the NIH Collaborative Influenza Vaccine Innovation Centers contract 75N93019C00051 and a grant from an anonymous philanthropist to Mount Sinai. Design and generation of reagents used in this project in the Kramer laboratory were supported by Centers of Excellence for influenza Research and Response (75N93021C00014) and Collaborative Influenza Vaccine Innovation Centers (75N93019C00051), as was the Garcia-Sastre laboratory. Research and development activities in Vietnam were funded by IVAC, the National Covid-19 Vaccine Fund and the Thien Tam Fund of Vingroup. Testing of clinical trial specimens at Nexelis was supported by CEPI. The authors thank Dr. Randy Albrecht for management of import/export of recombinant Newcastle disease virus at the Icahn School of Medicine at Mount Sinai. We are grateful to Dr. Nina Bardwaj for programmatic and scientific oversight of the Virus and Cell Therapy Laboratory at the Icahn School of Medicine at Mount Sinai, which produced the NDV-HXP-S master virus seed. The authors are grateful to Dr. Viviana Simon for providing the convalescent serum samples used in the live virus neutralization study. The authors thank the members of the DSMB for their work on this study. The authors are grateful to the volunteers who participated in this study.

