## Supplementary material for "Safety and Immunogenicity of An Egg-Based Inactivated Newcastle Disease Virus Vaccine Expressing SARS-CoV-2 Spike: Interim Results of a Randomized, Placebo-Controlled, Phase 1/2 Trial in Vietnam": IVAC NDX-HXP-S Phase 1 protocol

### APPENDIX A – CLINICAL TRIAL PROTOCOL

A Phase 1/2 Randomized, Placebo-controlled, Observer-blind Trial to Assess the Safety and Immunogenicity of COVIVAC Vaccine produced by IVAC in Adults Aged 18-75 Years in Vietnam

### A Phase 1/2 Randomized, Placebo-controlled, Observer-blind Trial to Assess the Safety and Immunogenicity of COVIVAC Vaccine produced by IVAC in Adults Aged 18-75 Years in Vietnam

**Trial Registration:** ClinicalTrials.gov NCT04830800.

**Vietnam Ministry of Health Study Number:** 1407/QD-BYT

**Version 1,0**

**February 05, 2021**

**Sponsored by:**
Institute of Vaccines and Medical Biologicals (IVAC)
Nha Trang, Viet Nam

**Principal Investigator:**
**Prof. Dang Duc Anh and Associate Prof. Vu Dinh Thiem**
Ha Noi, Vietnam

10.5.3. Analysis of Secondary Objectives ... 67
10.5.4. Analysis of Exploratory Objectives ... 67
10.6. Multiplicity ... 68
10.7. Handling of Subject Discontinuations and Missing Data ... 68
11. LABORATORY EVALUATIONS ... 68
11.1. Blood Sample Collection, Distribution, and Storage ... 68
11.2. Safety Clinical Laboratory Assays ... 69
11.3. Immunological Assays (Secondary Objectives) ... 69
11.4. Microbiological Assays ... 69
11.5. Assay Qualification, Standardization, and Validation ... 70
11.6. Biohazard Containment ... 70
12. DATA MANAGEMENT ... 70
12.1. Case Report Form Development and Completion ... 70
12.2. Record Archival ... 71
12.2.1. Archiving Data at Study Site ... 71
12.2.2. Data Storage and Archival ... 71
12.3. Posting of Information on Clinicaltrials.gov ... 72
12.4. Confidentiality ... 72
12.5. Publication ... 72
13. QUALITY ASSURANCE AND QUALITY CONTROL ... 72
13.1. General Considerations ... 72
13.2. External Monitoring ... 72
13.3. Independent Auditing ... 73
13.4. Regulatory Agency Auditing ... 73
14. OBLIGATIONS AND ROLES OF THE SPONSOR, PI, AND STUDY PERSONNEL ... 73
15. ETHICAL CONSIDERATIONS AND INFORMED CONSENT ... 74
15.1. Ethical Review ... 74
15.2. Informed Consent Process ... 75
15.3. Study-Related Injury ... 75
15.4. Risk/Benefit ... 75
15.5. Subject Confidentiality ... 76
15.6. Reimbursement ... 77

15.7. Storage of Specimens ... 77
16. APPENDICES ... 78
16.1. Appendix 1: Solicited Injection Site and Systemic Reactions Toxicity Grading Table ... 78
16.2. Appendix 2: Vital Signs Toxicity Grading Table ... 80
16.3. Appendix 3: Serum and Hematology Toxicity Grading Table ... 81
16.4. Appendix 4: Potential Immune-Mediated Medical Conditions (PIMMC) ... 82
16.5. Appendix 5: Adverse Events Specific to COVID-19 Disease ... 84
16.6. Appendix 6: Template of interim study report for transition to Phase 2 ... 85
16.7. Appendix 7: References ... 86

#### List of tables and figures in the protocol:

- Table 1. Summary of preclinical studies
- Table 2. COVIVAC Treatment Groups (Toxicology Study)
- Table 3. Treatment Groups
- Table 4. Phase 1 Dose Escalation Steps
- Table 5. Schedule of Study Visits and Procedures – Phase 1 and Phase 2
- Table 6. Severity Grading
- Table 7. Probability to observe at least one serious or severe adverse event by sample size and underlying event rate.
- Table 8. Power to select the candidate with the highest true mean to progress to second phase and overall for the Phase 1/2 overall by minimum fold-difference between the highest candidate mean and the other means, assuming log10 variability of 0.5.
- Table 9. Blood Volume Requirements

#### LIST OF ABBREVIATIONS AND ACRONYMS

| ABBREVIATION/ ACRONYM | DEFINITION |
|---|---|
| Ab | antibody |
| ACE2 | angiotensin-converting enzyme 2 |
| ADCC | antibody-dependent cellular cytotoxicity |
| ADE | antibody-dependent enhancement |
| ADR | adverse drug reaction |
| AE | adverse event |
| AESI | adverse event of special interest |
| ALT | alanine transaminase |
| ANCA | anti-neutrophil cytoplasmic antibody |
| ARDS | acute respiratory distress syndrome |
| AST | aspartate transaminase |
| ATC | anatomical therapeutic chemical |
| BMI | Body Mass Index |
| BPL | beta-propiolactone |
| C | Celsius |
| CAPA | corrective action and preventive action |
| CBER | Center for Biologics Evaluation and Research |
| CDC | Centers for Disease Control and Prevention |
| CI | confidence interval |
| Cm | Centimeter |
| CMI | cell-mediated immunity |
| COVID-19 | coronavirus disease 2019 (disease caused by SARS-CoV-2) |
| CREST | calcinosis, Raynaud's phenomenon, esophageal dysmotility, sclerodactyly, and telangiectasia |
| CRF | case report form |
| CRO | contract research organization |
| CSR | clinical study report |
| D | day(s) |
| DSMB | Data and Safety Monitoring Board |
| EC | Ethics Committee |
| EDC | electronic data capture |
| ELISA | enzyme-linked immunosorbent assay |
| ELISpot | enzyme-linked immunospot |
| ELU | ELISA Units |
| EOS | end of study |
| ER | Emergency Room |
| EUA | Emergency Use Authorization |
| F | Fahrenheit |
| FDA | US Food and Drug Administration |
| FIPV | feline infectious peritonitis virus |
| G | Group |
| GCP | good clinical practice |
| GLP | good laboratory practice |
| GMC | geometric mean concentration |
| GMFR | geometric mean fold rise |
| GMP | good manufacturing practice |
| GMT | geometric mean titer |

| | |
|---|---|
| HBsAg | hepatitis B surface antigen |
| hCG | serum human gonadotrophic hormone |
| HCV Ab | hepatitis C virus antibody |
| Hgb | hemoglobin |
| HIV 1/2 Ab | human immunodeficiency virus 1 and 2 antibody |
| HMU | Hanoi Medical University |
| HN | hemagglutinin-neuraminidase |
| HXP | HexaPro |
| IATA | International Air Transport Association |
| IB | Investigator's Brochure |
| ICF | informed consent form |
| ICH | International Conference on Harmonisation |
| ID | identification |
| IIV | inactivated influenza virus vaccine |
| IgG | immunoglobulin G |
| IL | interleukin |
| IM | intramuscular |
| IME | important medical event |
| IN | intranasal |
| INF | interferon |
| ISO | International Organization for Standardization |
| IP | investigational product |
| ISMMS | Icahn School of Medicine at Mount Sinai |
| IV | intravenous |
| IVAC | Institute of Vaccines and Medical Biologicals |
| IWRS | Interactive Web Response System |
| LMIC | low- and middle-income country |
| LSLV | last subject last visit |
| MAAE | medically-attended adverse event |
| MedDRA | Medical Dictionary for Regulatory Activities |
| MERS-CoV | Middle East respiratory syndrome coronavirus |
| MIS-C | multisystem inflammatory syndrome in children |
| mm | millimeter |
| MoH | Ministry of Health |
| N | nucleocapsid |
| NAb | neutralizing antibody |
| NDV | Newcastle disease virus |
| NICVB | National Institute for Control of Vaccines and Biological |
| NIHE | National Institute of Hygiene and Epidemiology |
| $NT_{50}$ | 50% neutralizing antibody titer |
| $NT_{80}$ | 80% neutralizing antibody titer |
| NTF | note to file |
| PBMC | peripheral blood mononuclear cell |
| PBS | phosphate-buffered saline |
| PE | physical examination |
| PI | principal investigator (the term is used throughout to indicate PI or designee) |
| PIMMC | potential immune-mediated medical conditions |
| Plt | platelet |
| PNA | pseudovirus neutralization assay |
| PPE | personal protective equipment |

| | |
|---|---|
| PP IMM | per protocol immunogenicity population |
| PSRT | Protocol Safety Review Team |
| qPCR | quantitative polymerase chain reaction |
| R | recovery |
| $R_0$ | reproduction number |
| RLU | relative luminescence units |
| RNA | ribonucleic acid |
| RSV | respiratory syncytial virus |
| RT-PCR | reverse transcription polymerase chain reaction |
| S | Spike |
| SAE | serious adverse event |
| SAP | statistical analysis plan |
| SARS-CoV | severe acute respiratory syndrome coronavirus |
| SARS-CoV-2 | severe acute respiratory syndrome coronavirus 2 |
| SC | subcutaneous |
| SOC | system organ class |
| SOP | standard operating procedure |
| SSP | study-specific procedure |
| SUSAR | suspected unexpected serious adverse reaction |
| TEAE | treatment-emergent adverse event |
| TEN | toxic epidermal necrolysis |
| Th | T-helper cell |
| TLR-9 | toll-like receptor 9 |
| TMF | trial master file |
| US | United States |
| USP | United States Pharmacopeia |
| V | Visit number (e.g. V1 = first study visit) |
| WBC | white blood cell |
| WHO | World Health Organization |
| WMA | World Medical Association |
| WOCBP | Women of childbearing potential |
| WT | wild-type |

#### STATEMENT OF COMPLIANCE

I am Principal Investigator of the study "A Phase 1/2 Randomized, Placebo-controlled, Observer-blind Trial to Assess the Safety and Immunogenicity of COVIVAC Vaccine produced by IVAC in Adults Aged 18-75 Years in Vietnam"; signing below ensures that the study will be carried out on schedule, according to the content of approved protocol, and in accordance with Good Clinical Practice (GCP) as required by applicable rules of Vietnam: Decision No. 799/2008/QD-BYT dated 07 Mar, 2008, on "Guidance on Good Clinical Practice," Circular No. 29/2018/TT-BYT dated 29 Oct, 2018 on "Regulation on Clinical Trial" and the Circular No. 32/2018/TT/BYT dated 12 Nov, 2018 on "Regulation on Medical Product Registration and Input Materials," Decision 1248/QD BYT dated 11 Jan, 2013, on the National Guidelines on ethical issues in biomedical research; Circular 04/2020/TT-BYT dated 05 Mar, 2020 on the Establishment, Functions and Responsibilities of Biomedical Ethics Committee, and Dispatch No 62/QĐ-K2ĐT dated 02 Jun, 2017, on the guidelines on recording/reporting SAEs in a clinical trial. And the Decision No 3659/QĐ-BYT dated 21 Aug, 2020 on "Guidance on Research, Clinical Trial, Licensing and Use of COVID-19 vaccine".

The study informed consent documents will embody the elements of consent as described in the Declaration of Helsinki.

The study will also be carried out in accordance with Good Clinical Practice (GCP) as required by the following:

- International Conference on Harmonisation (ICH) Guidance for GCP E6(R2)
- World Medical Association (WMA) Declaration of Helsinki – Ethical Principles for Research Involving Human Subjects (Oct 2013 or subsequent amendments)

All key personnel (all individuals responsible for the design and conduct of this study) have completed Human Subjects Protection Training and ICH-GCP training prior to interaction with any participants or to have access to their confidential study data.

**PROTOCOL SIGNATURE PAGE**

The signature below constitutes the approval of this protocol and the attachments, and provides the necessary assurances that this trial will be conducted according to all stipulations of the protocol, including all statements regarding confidentiality, and according to local legal and regulatory requirements and ICH-GCP guidelines as outlined in the 'Statement of Compliance.'

Site Investigator:

Signed: ________________________________________ Date: ____________________

Name: Associate Prof. Vu Dinh Thiem, MD

Title: Director of Clinical Trial Center, NIHE

Signed: ________________________________________ Date: ____________________

Name: Prof. Dang Duc Anh, MD

Title: Director of NIHE

Sponsor's Representative:

Signed: ________________________________________ Date: ____________________

Name:

Title:

#### KEY ROLES AND CONTACT INFORMATION

| | |
|---|---|
| Sponsor | Institute of Vaccines and Medical Biologicals (IVAC) |
| Clinical Trial Center | National Institute of Hygiene and Epidemiology (NIHE) |
| Principal Investigator | Prof. Dang Duc Anh |
| Co- Principal Investigator | Associate Prof. Vu Dinh Thiem, MD |
| Vaccine Manufacturer | IVAC |
| Contract Research Organization for clinical monitoring | Vietstar |
| Contract Research Organization for data management | Local CRO with experience and collaboration with international CRO on data management will be contracted in order to meet the requirements in the ICH E6-R2 and electronic data security. |
| Clinical site | Hanoi Medical University (HMU) for Phase 1.<br>Vu Thu District Hospital, Thai Binh province for Phase 2 |
| Immunology Laboratory | Nexelis<br>525 Cartier West Blvd<br>Laval, Quebec<br>Canada, H7V 3S8 |
| Clinical Laboratory | Clinical lab of HMU's Hospital.<br>No 1, Ton That Tung Street, Hanoi, Vietnam |
| Microbiology Laboratory | Laboratory of virus transmitted from animal to human, Virus Lab Department of NIHE |
| Ethics Committees | NIHE's IRB and Independent Ethics Committee of MoH |
| National Regulatory Agency | Ministry of Health (MoH) in Vietnam |
| Sponsor Medical Monitor: | Associate Prof. Le Van Be |
| Technical support | PATH Vietnam office<br>11th Floor, Hanoi Towers<br>49 Hai Ba Trung<br>Hoan Kiem District<br>Hanoi, Vietnam<br><br>PATH US office<br>2201 Westlake Avenue, Suite 200.<br>Seattle, WA 98121, USA |

### 1. PROTOCOL SUMMARY

| | |
|---|---|
| **TITLE** | **A Phase 1/2 Randomized, Placebo-controlled, Observer-blind Trial to Assess the Safety and Immunogenicity of COVIVAC Vaccine produced by IVAC in Adults Aged 18-75 Years in Vietnam** |
| **STUDY NUMBER** | |
| **PROJECT PHASE** | Phase 1/2 |
| **INVESTIGATIONAL PRODUCT(S)** | **Investigational Vaccine:**<br>Inactivated Newcastle Disease Virus (NDV) chimera expressing a trimeric pre-fusion form of the SARS-CoV-2 Spike (S) protein that contains six proline mutations (HexaPro, HXP); for intramuscular (IM) administration. One formulation of COVIVAC evaluated in this study includes 1.5 mg of the adjuvant CpG 1018 (a 22-mer phosphorothioate-linked oligodeoxynucleotide).<br>**Placebo:**<br>Phosphate-buffered saline (PBS); for IM administration |
| **STUDY HYPOTHESES** | **Safety, Tolerability:**<br>• COVIVAC will have an acceptable safety profile and be well-tolerated at all dose levels when administered as a two-dose regimen either without adjuvant (at 1 µg, 3 µg, and 10 µg of S protein per 0.5 ml dose), or with the adjuvant CpG 1018 (at 1 µg of S protein per 0.5 ml dose).<br>**Immunogenicity:**<br>• COVIVAC will elicit a measurable and dose-dependent neutralizing antibody response. |
| **STUDY OBJECTIVES – PHASE 1** | **Primary Objective:**<br>**Safety, Tolerability:**<br>1. To assess the safety and tolerability of each of four formulations of COVIVAC (at 1, 3 and 10 µg without adjuvant; and at 1 µg with CpG 1018) after the first and second dose of a two-dose regimen administered at a 28-day interval to adults aged 18-59 years<br>**Secondary Objectives:**<br>**Immunogenicity:**<br>1. To assess the functional (neutralizing) humoral immune response elicited by each formulation of NDV-HXP-S, as measured by a SARS CoV-2 pseudovirus-based neutralization assay (PNA), at baseline, 28 days after the first vaccination, and 14 days after the second vaccination (to inform the selection of two formulations for evaluation in Phase 2), and at 6 months and 12 months after the second vaccination, in adults aged 18-59 years<br>2. To assess the Immunoglobulin G (IgG) immune response elicited by each formulation of COVIVAC against the S protein of SARS-CoV-2, as measured by enzyme-linked immunosorbent assay (ELISA), at baseline, 28 days after the first vaccination, and 14 days, 6 months and 12 months after the second vaccination, in adults aged 18-59 years |

<table>
<tr><td>STUDY OBJECTIVES – PHASE 2</td><td>Primary Objective:<br>Safety, Tolerability:<br>1. To assess the safety and tolerability of two selected formulations of COVIVAC (based on Phase 1 safety and immunogenicity) after the first and second dose of a two-dose regimen administered at a 28-day interval to adults aged 18-75 years<br>2. To assess the safety and tolerability of two selected formulations of COVIVAC after the first and second dose of a two-dose regimen administered at a 28-day interval to adults aged 18-59 years and 60-75 years<br>Secondary Objectives:<br>Immunogenicity:<br>1. To assess the functional (neutralizing) humoral immune response elicited by each of two selected COVIVAC formulations, as measured by PNA, at baseline, 28 days after the first vaccination, and 14 days, 6 months and 12 months after the second vaccination, in adults aged 18-75 years<br>2. To assess the functional (neutralizing) humoral immune response elicited by each of two selected COVIVAC formulations, as measured by PNA, at baseline, 28 days after the first vaccination, and 14 days, 6 months and 12 months after the second vaccination, in adults aged 18-59 years and 60-75 years<br>3. To assess the IgG immune response elicited by each of two selected COVIVAC formulations against the S protein of SARS-CoV-2, as measured by ELISA, at baseline, 28 days after the first vaccination, and 14 days, 6 months and 12 months after the second vaccination, in adults aged 18-75 years<br>4. To assess the IgG immune response elicited by each of two selected COVIVAC formulations against the S protein of SARS-CoV-2, as measured by ELISA, at baseline, 28 days after the first vaccination, and 14 days, 6 months and 12 months after the second vaccination, in adults aged 18-59 years and 60-75 years<br>Exploratory Objectives (Immunogenicity):<br>1. To assess the S protein-specific T cell response elicited by each of two selected COVIVAC formulations, as measured by enzyme-linked immunospot assay (ELISpot), at baseline, and 14 days and 6 months after the second vaccination, in adults aged 18-75 (subset of subjects)<br>2. To assess the S protein-specific T cell response elicited by each of two selected COVIVAC formulations, as measured by ELISpot, at baseline, and 14 days and 6 months after the second vaccination, in adults aged 18-59 years and 60-75 years (subsets of subjects)<br>3. To assess the IgG immune response elicited by each of two selected COVIVAC formulations against the hemagglutinin-neuraminidase (HN) protein of NDV, as measured by ELISA, at baseline, 28 days after the first vaccination, and 14 days and 12 months after the second vaccination, in adults aged 18-59 years</td></tr>
<tr><td>STUDY ENDPOINTS</td><td>Primary Endpoints:<br>Safety, Tolerability:<br>• Number and severity of solicited local and systemic adverse events (AEs) during the first 7 days after each vaccination</td></tr>
</table>

| | |
|---|---|
| | • Number, severity, and relatedness of clinically significant hematological and biochemical measurements at 7 days post each vaccination<br>• Number, severity and relatedness of all unsolicited AEs during the first 28 days after each vaccination<br>• Number, severity and relatedness of serious adverse events (SAEs) throughout the entire study period<br>• Number, severity and relatedness of medically-attended AEs (MAAEs) throughout the entire study period<br>• Number, severity and relatedness of adverse events of special interest (AESI) throughout the entire study period, including AESI relevant to COVID-19, and potential immune-mediated medical conditions (PIMMC)<br>**Secondary Endpoints:**<br>**Immunogenicity:**<br>**For Secondary Objective 1 (Phase 1), and Objectives 1 and 2 (Phase 2):**<br>• 50% and 80% neutralizing antibody ($NT_{50}$ and $NT_{80}$) geometric mean titer (GMT) against SARS-CoV-2 pseudovirus at 28 days after the first vaccination, and 14 days, 6 months and 12 months after the second vaccination in subjects who are anti-S IgG seronegative at baseline<br>• Geometric mean fold rise (GMFR) (from baseline) in $NT_{50}$ and $NT_{80}$ against SARS-CoV-2 pseudovirus at 28 days after the first vaccination, and 14 days, 6 months and 12 months after the second vaccination<br>• Percentage of subjects with $NT_{50}$ and $NT_{80}$ seroresponses against SARS-CoV-2 pseudovirus as defined by (1) a ≥ 4-fold increase from baseline, and (2) a ≥ 10-fold increase from baseline at 28 days after the first vaccination, and 14 days, 6 months and 12 months after the second vaccination<br>**For Secondary Objective 2 (Phase 1), and Objectives 3 and 4 (Phase 2):**<br>• Anti-S IgG GMT at 28 days after the first vaccination, and 14 days, 6 months and 12 months after the second vaccination in subjects who are anti-S IgG seronegative at baseline<br>• GMFR (from baseline) in anti-S IgG GMT at 28 days after the first vaccination, and 14 days, 6 months and 12 months after the second vaccination<br>• Percentage of subjects with seroresponses in anti-S IgG titer as defined by (1) a ≥ 4-fold increase from baseline, and (2) a ≥ 10-fold increase from baseline, at 28 days after the first vaccination, and 14 days, 6 months and 12 months after the second vaccination<br>**For Exploratory Objectives 1 and 2 (Phase 2):**<br>• Magnitude, functionality, and T-helper cell (Th) polarization of S protein-specific T cells relative to baseline at 14 days and 6 months after the second vaccination<br>**For Exploratory Objective 3 (Phase 2):** |

| | |
|---|---|
| | • Anti-NDV HN IgG GMT at baseline, 28 days after the first vaccination, and 14 days and 6 months after the second vaccination |
| **STUDY DESIGN** | This prospective, single-center, randomized, placebo-controlled, observer-blind Phase 1/2 study includes two separate parts.<br><br>Part 1 is a first-in-human, Phase 1 study designed to evaluate the safety, tolerability and immunogenicity of the COVIVAC vaccine at three different dose levels (1, 3, and 10 µg) without adjuvant, and at one dose level (1 µg) with the adjuvant CpG 1018, in a total of 120 subjects aged 18-59 years. Eligible subjects will be randomly assigned to receive study vaccine or placebo as an IM injection in the deltoid muscle at Visit [V]1 as part of one of four sequential groups of escalating antigen dose level or added adjuvant. Each group will be divided into two sequential subgroups, a sentinel subgroup (n=6) and a larger subgroup (n=24) that includes the remaining subjects to be evaluated at that dose level (or for that formulation in the case of the one adjuvanted treatment group). Initially, a sentinel subgroup of 6 subjects will be randomized to receive COVIVAC 1 µg or placebo in a 5:1 ratio. Sentinel subjects will be monitored for reactogenicity and safety through Day 8 (Visit 2), including for the initial 24 hours post-injection in clinic. If the Protocol Safety Review Team (PSRT) has no safety concerns based on blinded review of all safety data, including clinical laboratory results, through Visit 2, the second subgroup of 24 subjects will be randomized to receive COVIVAC 1 µg or placebo at the same ratio; on the same day, the sentinel subgroup (n=6) for the second dose level to be evaluated (COVIVAC 3 µg) can be randomized to receive study vaccine or placebo in a 5:1 ratio, and so on for the subsequent dose level and formulation (see Table 4).<br><br>The PSRT will review blinded safety, reactogenicity and clinical laboratory data accrued on a weekly basis until D43 of the last subject in each phase, then at least once monthly until the end of the study. Subjects will receive 2 doses of assigned investigational product (IP) on D1 and D29 (V1 and V3), and be assessed in clinic for safety and reactogenicity at 7 days after each vaccination (V2 and V4), for safety at 28 days after the second vaccination (V6), and for safety and immunogenicity assessments at baseline (V1), 28 days after the first vaccination (V3) and 14, 168 and 336 days after the second vaccination (V5, V7, and V8, end-of-study [EOS] Visit) (see Visit Schedule Table below).<br><br>An interim analysis of Phase 1 data conducted after the last subject last visit for V6 (D57) will serve as the basis for decisions about down-selection and advancing to Part 2 of the study (Phase 2). Down selection and advancement to Part 2 (Phase 2) will be based on the following parameters:<br>• Post-dose 2 immunogenicity results at the aggregate treatment level<br> o A threshold immune response at Day 43 will be required: the observed seroresponse rate in a treatment group (defined as the percentage of subjects with at least a 4-fold rise from baseline in 80% neutralizing antibody titers) will need to be ≥52% at the LL of the 95% CI for that treatment (vaccine formulation) to be considered for advancement to Phase 2.<br>• Post-dose 1 and post dose 2 safety results including all solicited and unsolicited adverse events, serious adverse events, and clinical laboratory results. |

| | |
|---|---|
|  | The following process will be followed for the decision about down selection and advancing to Part 2 (Phase 2):<br>• The DSMB will review the unblinded safety data and provide a recommendation to the Sponsor on whether the safety profile is acceptable for advancing a formulation to Phase 2.<br>• The Sponsor will review the DSMB recommendation in conjunction with the immunogenicity data and select two formulations to advance to Phase 2.<br> ○ If multiple formulations achieve the threshold immune response (as well as have an adequate safety and tolerability profile per the DSMB), the Sponsor will select two formulations to advance to Phase 2 based on consideration of such factors as the relative functional immunogenicity of these formulations, opportunity for dose sparing, and opportunity to limit cost and possible supply constraints associated with use of the CpG adjuvant.<br>• The selection and recommendation to advance to Phase 2 along with the interim report will be jointly reviewed by NIHE's IRB and MoH prior to Phase 2 enrollment.<br>In Part 2 of this combined Phase 1/2 study, 300 adults aged 18-75 years will be randomized (2:5:5) to placebo, or one of two selected formulations of COVIVAC being evaluated in Phase 1. At least one-third of the subjects in Phase 2 will be aged ≥60 years to ensure that adequate safety and immune data will be available from older and elderly adults to inform the selection of the COVIVAC formulation to advance to Phase 3 studies. The Phase 2 cohort will follow the same visit schedule, and undergo the same procedures and assessments, as in Phase 1. In addition, as exploratory objectives, the anti-NDV HN IgG response will be assessed at V1, V3, V5, and V8 in all subjects, and 36 subjects (equally distributed between the two age strata) will be randomly selected in a 1:1:1 ratio to provide additional blood at V1, V5 and V7 to be used to isolate peripheral blood mononuclear cells (PBMCs) for assessment of T-cell-mediated immunity (CMI).<br><br>An interim analysis of Phase 2 data (combined with immunogenicity and safety data generated for the placebo group and two selected COVIVAC formulations from Phase 1) will be conducted after the last subject of the Phase 2 cohort completes V6 (D57) as the basis for selecting the optimal formulation of COVIVAC to advance to Phase 3 studies. As was the case for the Phase 1 interim analysis at the same timepoint, the data generated will include unblinded post-dose 1 and dose 2 safety results for review by the DSMB, and immunogenicity results aggregated by treatment group for review by the Sponsor. The DSMB will consider all accumulated safety data from both phases of the study prior to making any recommendation to the Sponsor that it not advance a formulation based on safety concerns. The Sponsor will ultimately select the formulation to advance to Phase 3 that, in addition to having been judged by the DSMB to have an adequate safety and tolerability profile, is optimal based on relative functional immunogenicity and other programmatic considerations such as those noted above. |

**TREATMENT GROUPS**

| Phase | Group | Sample Size | Study Product |
|---|---|---|---|
| 1 | 1 | 20 | Placebo |
| | 2 | 25 | COVIVAC 1 µg |
| | 3 | 25 | COVIVAC 3 µg |
| | 4 | 25 | COVIVAC 10 µg |
| | 5 | 25 | COVIVAC 1 µg + CpG 1018 1.5 mg |
| 2 | 1 | 50 | Placebo |
| | 2 | 125 | Formulation A |
| | 3 | 125 | Formulation B |

**VISIT SCHEDULE – PHASE 1 AND PHASE 2**

| | | V1 | V2 | V3 | V4 | V5 | V6 | V7 | V8 |
|---|---|---|---|---|---|---|---|---|---|
| Day (+ window) | -42 to 1 | 1 | 8 (+3) | 29 (+3) | 36 (+3) | 43 (+3) | 57 (+7) | 197 (+14) | 365 (+28) |
| Screening | X | | | | | | | | |
| Vaccination | | 1 | | 2 | | | | | |
| Immune labs Pseudovirus ELISAs* | | X | | X | | X | | X | X |
| CMI# | | X | | X | | X | | X | |
| Safety data collection: | | | | | | | | | |
| Safety visits | X | X | X | X | X | X | X | X | X |
| Clinical labs | X | | X | | X | | | | |
| Reactogenicity | | | | | | | | | |
| AEs | | | | | | | | | |
| MAAEs | | | | | | | | | |
| AESI | | | | | | | | | |
| SAEs | | | | | | | | | |

* Anti-NDV ELISA in Phase 2 only, at V1, V3, V5, and V8

### Phase 2 only, in subset of subjects

| | |
|---|---|
| **STUDY POPULATION** | A total of 420 male and female adults aged 18-75 (both inclusive) will receive Investigational Product (IP) and be evaluated for safety and immunogenicity as part of this combined Phase 1/2 study. At least one-third of the subjects evaluated in Phase 2 will be ≥60 years old. Phase 1 of the study will be conducted at Hanoi Medical University (HMU) and Phase 2 of the study will be conducted at Vu Thu district health center, Thai Binh province. |
| **STUDY DURATION** | Subjects will be followed for approximately 13 months after randomization (12 months following the 2$^{nd}$ dose of IP). |

#### 2. BACKGROUND AND RATIONALE

On 7 January 2020, a novel coronavirus was identified as the cause of a cluster of pneumonia cases first detected in December 2019 in Wuhan, the capital of China's Hubei province,[1] marking the third documented zoonotic transmission event of a coronavirus within the last two decades. Two other highly pathogenic coronaviruses,[1] both of the genus *Betacoronavirus*, also crossed the species barrier during this period: severe acute respiratory syndrome virus (SARS-CoV), which emerged in Guangdong province, China in November 2002, and ultimately led to 8,096 reported cases and 774 deaths in 27 countries before the last infection was detected in July 2003; and the Middle East respiratory syndrome coronavirus (MERS-CoV), which was first detected in Saudi Arabia in June 2012 (where it still circulates in camels) and has led to 2494 laboratory-confirmed cases and 858 deaths.[2] Phylogenic analysis from full-genome sequencing of the novel coronavirus (which became publicly available on 12 January 2020 [GenBank accession no. MN908947.2] indicated that it was also a Betacoronavirus, and that it belonged to the same subgenus (subgenus *Sarbecovirus* [subgroup B]) as SARS-CoV, with which it shares 79% nucleotide identity.[3] Given its taxonomic relationship to SARS-CoV, the novel coronavirus was designated severe acute respiratory syndrome virus 2 (SARS-CoV-2) by the International Committee on Taxonomy of Viruses. By 30 January 2020, the international outbreak of SARS-CoV-2 was declared a Public Health Emergency of International Concern by the World Health Organization (WHO), which on 11 February 2020 named the disease caused by the virus COVID-19 (coronavirus disease 2019). By March 11, 2020, WHO officially declared the outbreak of COVID-19 a global pandemic.

##### 2.1. Burden of Disease

As of 06 January, 2021, the confirmed global case count of COVID-19 has risen to over 87 million, and deaths directly due to SARS-CoV-2 infection have surpassed 1.85 million.[4] The large majority of deaths have occurred among the elderly in the United States, Brazil, India and Mexico, which may be largely attributable to poorly implemented control measures (such as surveillance, border closure, quarantine and social distancing) in those countries.[5] While there have been significantly fewer cases to date in most low- and middle-income countries (LMICs), there is great concern about the potential impact of easing of containment efforts, particularly in countries where robust and functioning public health infrastructure is lacking.[6] In addition to the potentially devastating direct disease burden of SARS-CoV-2 in LMICs, the disruptions to routine health services and economic systems due to the pandemic may result in an indirect disease burden of equal or even greater magnitude – a burden that would be borne by all demographic groups. One study has estimated that disruptions to health systems and access to food in LMICs as a result of the COVID-19 pandemic would result in a minimum of 253,500 children under 5 child deaths and 12.200 maternal deaths over a 6-month period. In the worse scenario, there would be 1,157,000 additional under-five child deaths and 56.700 maternal deaths over a 6-month period.[7] In Vietnam, as of 06 January 2021 there have been 1.504 confirmed cases and 35 deaths.

##### 2.2. Pathogen and Clinical Disease

Like all viruses in the *Coronaviridae* family, SARS-CoV-2 is an enveloped, positive-sense, single-stranded RNA virus, with a genome that (in addition to a replicase and accessory proteins) encodes four major structural proteins: the spike surface glycoprotein (S), small envelope protein, matrix protein, and nucleocapsid (N) protein. The diagnosis of COVID-19 is made by detection of SARS-CoV-2 RNA by reverse transcription polymerase chain reaction (RT-PCR), with a common gene target being one of the structural proteins. The coronavirus S protein plays the essential role of mediating cell entry by binding to the host-cell receptor, and thus is the primary determinant of viral tropism.[8] The S proteins of SARS-CoV and SARS-CoV-2 bind to the same primary host receptor, the angiotensin-converting enzyme 2 (ACE2) receptor, whereas MERS-CoV (which belongs to a different coronavirus lineage) uses dipeptidyl peptidase 4.

As is the case for both SARS-CoV and MERS-CoV, the median incubation period of SARS-CoV-2 is approximately four to five days after exposure.[9] Similar to SARS-CoV, pneumonia is the most frequent serious manifestation of SARS-CoV-2 infection, with dry cough, fever, myalgia, headache, dyspnea and sore throat being the most common presenting clinical features.[10] Diarrhea, nausea/vomiting, abdominal pain,[11] smell and taste disorders (i.e., anosmia and dysgeusia),[12] and rhinorrhea are also associated with COVID-19, though less commonly. While the spectrum of symptomatic SARS-CoV-2 infection ranges from mild to critical, most cases of COVID-19 are mild (approximately 80%).[13] In addition, multiple studies now suggest that asymptomatic infection may be very common – as high as 88% by one report.[14] This is in contrast to SARS-CoV, which uncommonly caused mild or asymptomatic disease.[15]

In patients with severe to critical disease (approximately 15% and 5% of total cases, respectively),[13] dyspnea, hypoxia, cytokine release syndrome, acute respiratory distress syndrome, shock, thromboembolic complications, sudden cardiac death and multiorgan failure have been reported.[16] Most patients who die from COVID-19 are older persons (~80% of deaths occur in those > 65 years old)[17] and/or persons with underlying medical comorbidities (obesity, cardiovascular disease, diabetes, chronic lung disease, cancer, chronic kidney disease, hypertension),[18] although severe illness can occur in otherwise healthy persons of any age. In rare cases children can be severely affected, manifesting clinically as a hyperinflammatory syndrome similar to Kawasaki disease (a rare acute pediatric vasculitis)[19] or toxic shock syndrome;[20] this syndrome has been termed multisystem inflammatory syndrome.[21] In multiple cohort studies, male gender is strongly associated with death from COVID-19.[22,23]

#### 2.3. Transmission and Infectivity

Like SARS-CoV, there is considerable evidence that SARS-CoV-2 originated in bats, and was transmitted to humans after amplification in an intermediate host such as the pangolin.[24] Human-to-human transmission is thought to occur as a result of direct contact of virus with the mucous membranes of the infected person, primarily via infected respiratory droplets and aerosols. The risk of transmission from an individual with SARS-CoV-2 infection varies by the type and duration of exposure, factors such as the amount of virus in respiratory secretions, and the use of preventive measures. The interval during which an individual with COVID-19 is infectious appears to be highly variable, with duration of viral shedding dependent upon the severity of the illness.[25] However, it is known that SARS-CoV-2 can be transmitted prior to, or in the absence of the development of symptoms, and throughout the course of illness, with highest levels of virus in the upper respiratory tract – and thus perhaps highest infectivity – soon after symptom onset.[26] It is not known to what extent asymptomatic infection has contributed to the pandemic spread of the virus.[27]

#### 2.4. Vaccine Development

Based on a basic reproduction number ($R_0$) of ~2-3,[28] 60% to 70% of the population would need to develop immunity to SARS-CoV-2 for pandemic spread to be contained in the absence of significant control measures. Recent seroprevalence surveys indicate that population immunity remains well below this threshold in countries with the highest burden of COVID-19.[29] These results make clear that herd immunity cannot be achieved through natural infection without incurring very significant additional costs to society in terms of mortality, morbidity, and ongoing economic and social disruption, and that the only acceptable route to achieving herd immunity is widespread – and ideally rapid – deployment of an effective vaccine.

Studies conducted previously in support of SARS-CoV and MERS-CoV vaccine development were instrumental in accelerating development of SARS-CoV-2 candidates. The fact that many COVID-19 vaccine programs have moved quickly with selection of the S protein as target antigen was due to prior preclinical and human immunology studies on SARS-CoV demonstrating that the S protein of SARS-CoV elicits neutralizing antibodies that are protective against SARS-CoV challenge (by blocking virus attachment and fusion, as well as possibly by triggering FcR-mediated cytolysis or phagocytosis), and that these antibodies persist in convalescent serum of patients who have recovered from SARS.[30] Not long after the onset of the COVID-19 pandemic it was similarly demonstrated that the S protein of SARS-CoV-2 induces anti-S IgG antibodies in infected humans that neutralize the virus in animal models,[31] and that these antibodies persist for many months in convalescent sera.[32] While it is likely that induction of CD8+ T-cells against S as well as other viral proteins [33] plays a complementary role in protection after natural infection, the importance of a robust anti-S IgG neutralizing antibody response in protection against disease after natural infection[34] or experimental vaccination[35] is now incontrovertible.,

While there is currently no licensed vaccine to prevent COVID-19, a large number of vaccine candidates employing a large diversity of vaccine platforms (mRNA, DNA, subunit, viral vectors, nanoparticle) are in development, including 11 candidates that are currently in late stage Phase 3 development.[36] Two of these vaccines, Pfizer/BioNTech's and Moderna's mRNA-based vaccines that encode the full-length S protein with two stabilizing proline mutations (S-2P) have recently been shown in interim analyses to have an efficacy against COVID-19 of greater than 90% after a two-dose regimen. These results, along with supportive early-phase study results from multiple other S-protein-based candidate vaccines that have elicited high-titer neutralizing antibodies further validate the selection of the S protein as target antigen.[37]

#### 2.5. Risk of Immune Enhancement

In addition to facilitating target antigen selection, preclinical studies of SARS-CoV and MERS-CoV vaccines have also highlighted the potential risk of a SARS-CoV-2 vaccine exacerbating disease caused by subsequent SARS-CoV-2 infection. An inactivated SARS-CoV vaccine was shown to induce disease enhancement in rhesus monkeys subsequently challenged with SARS-CoV, as a result of non-neutralizing antibodies generated by the vaccine against the SARS-CoV S protein.[38] Such antibody-dependent enhancement (ADE) also occurred in cats infected with the Alphacoronavirus feline infectious peritonitis virus (FIPV), after being vaccinated with an inactivated FIPV vaccine.[39] Vaccine-induced non-functional antibodies cause ADE by facilitating viral entry into host cells, by increasing binding efficiency of virus-antibody complexes to host cells via Fc receptors on those cells.[40]

A second mechanism by which immune enhancement of disease may occur is via vaccine-induced inflammation.[41] Apart from activating Fc-receptor mediated endocytosis, vaccine-induced antibodies may elicit Fc-mediated responses, such as complement activation and antibody-dependent cellular cytotoxicity (ADCC), that may contribute to immunopathology. This phenomenon was observed in ferrets and Cynomolgus monkeys challenged with SARS-CoV following vaccination with an inactivated whole SARS-CoV vaccine;[42] the animals developed lung immunopathology characterized by eosinophilic recruitment, increased mucus production and airway hyperresponsiveness. The same immunopathology was observed in 1966 in children given formalin-inactivated respiratory syncytial virus (RSV) vaccine who subsequently were infected by RSV.[43] Enhancement of RSV disease in that study led to frequent hospitalizations and, in the youngest cohort, two deaths.[44] The prominence of eosinophils with such lung immunopathology has often been interpreted as signifying that this immune enhancement was the result of Th2-biased immune responses.

To date, there has been no evidence of antibody-dependent disease enhancement or immunopathology in animal studies and human trials of SARS-CoV-2 candidate vaccines. This is likely due to the fact that these candidates, including an inactivated SARS-CoV-2 vaccine,[45] have been shown to elicit high-titer anti-S neutralizing antibody responses. SARS-CoV-1 vaccines that elicit neutralizing antibodies against the SARS-CoV S protein were similarly able to protect animals from SARS-CoV challenge without evidence of acute lung injury and immunopathology.[46] In addition, many SARS-CoV-2 vaccine candidates include Th1-polarizing adjuvants to minimize the risk of disease enhancement. Nevertheless, disease enhancement remains a theoretical risk, particularly after waning of antibody responses, and therefore needs to be closely monitored in human clinical trials of SARS-CoV-2 vaccines.

#### 2.6. Introduction to COVIVAC Vaccine

Given the global scope of the COVID-19 pandemic, there is an urgent need for a safe and effective vaccine against COVID-19 that will be affordable and able to be manufactured at a sufficient scale to supply Vietnam and other low- and-middle-income countries. To achieve this aim, IVAC and two other consortium manufacturers – Government Pharmaceutical Organization (GPO), Thailand, and Instituto Butantan, Brazil – are independently developing an inactivated whole chimeric virion vaccine based on a Newcastle Disease Virus (NDV) that has been modified to also express the SARS-CoV-2 S protein on its surface. This technology was invented at the Icahn School of Medicine at Mount Sinai (ISMMS) (New York, New York, USA). Since NDV grows well in embryonated chicken eggs, the NDV chimeric vaccine can be manufactured using the same inexpensive egg-based process employed for inactivated influenza virus vaccine (IIV).[47] In addition to taking advantage of the cost-effectiveness of the manufacturing process, an inactivated chimeric NDV vaccine propagated in eggs (prior to being inactivated by beta-propiolactone [BPL]) is also expected to be as safe as egg-based IIV, which has an excellent safety profile, including in infants, pregnant women, and elderly adults.

As in the case of many COVID-19 vaccines currently in development, the inactivated chimeric NDV vaccine expresses the ectodomain of the SARS-CoV-2 S protein – which is fused to the F protein transmembrane domain of NDV, resulting in abundant expression of membrane-bound S antigen (see Figure 1). While many candidate COVID-19 vaccines are targeting the S protein as antigen, the S protein expressed on the surface of the chimeric NDV virion is a much more stable construct than that utilized in other COVID-19 vaccines as it contains six stabilizing proline substitutions – four more than are introduced in the S protein antigen (S-2P) used in a number of COVID-19 candidate vaccines, including the Pfizer/BioNTech and Moderna vaccines. The resulting protein, called "HexaPro" (HXP) and developed by the University of Texas (Austin, TX, USA),[48] is more immunogenic (>10-fold higher) than S-2P in mice (data unpublished), and also provides higher yields than S-2P when expressed recombinantly. As a result of these properties it is expected that significant dose-sparing will be possible with the COVIVAC vaccine candidate relative to other vaccine candidates expressing the earlier-developed S-2P antigen. Dose-sparing translates to higher production volumes and lower cost of vaccine. Finally it is important to note that HexaPro has also been shown to be more thermostable than S-2P.[48] The fact that COVIVAC is expected to be thermally stable at 2-8 °C is a major advantage of the vaccine over other COVID-19 candidates in development that will require storage at -20°C or even lower temperature.

**Figure 1. Electron micrograph of COVIVAC (courtesy of GPO Thailand)**

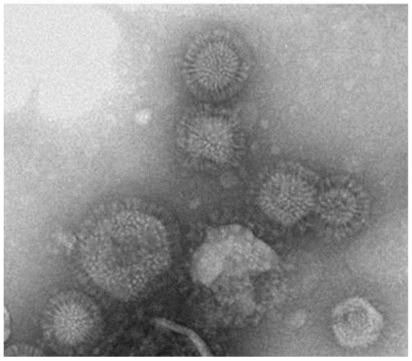

While it is likely that the whole virion COVIVAC vaccine alone will prove to be highly immunogenic in healthy adults due to endogenous adjuvanticity of the NDV virion, use of exogenous adjuvant may further increase the functional immune response to the vaccine – which may be important to ensure adequate protection in older adults, or possibly provide additional opportunity for dose-sparing. For this reason, the first part of this combined Phase 1/2 study is a first-in-human, Phase 1 evaluation of a 2-dose regimen of both unadjuvanted COVIVAC (at 1 µg, 3 µg, and 10 µg dose levels) and COVIVAC adjuvanted with CpG 1018 (at the 1 µg dose level) in healthy adults. CpG 1018, the adjuvant in the US-licensed HEPLISAV-B® vaccine (Dynavax Technologies Corporation, Emeryville, CA, USA), is a synthetic 22-mer oligodeoxynucleotide that exerts its action by targeting toll-like receptor 9 (TLR-9) expressed on a few key immune-cell types. When used as a vaccine adjuvant, CpG 1018 increases antibody concentrations to co-administered antigen, stimulates helper (CD4+) and cytotoxic (CD8+) T cell populations and generates robust T and B cell memory responses. Additionally, CpG 1018 strongly favors development of the Th1 subset of helper T cells, the type of helper T cell that is essential for protection from infections with viruses and intracellular bacteria. HEPLISAV-B was well-tolerated and shown to have an adequate safety profile in clinical trials in adults.[49] Interim analyses of an ongoing post-marketing study of over 30,000 HEPLISAV-B recipients and more than 35,000 active comparator recipients have raised no safety concerns.

#### 2.7. Summary of Nonclinical Studies

A complete summary of the COVIVAC preclinical development program is provided in the Investigator's Brochure (IB).

In brief, a pilot immunogenicity study in BALB/c mice and challenge study in golden Syrian hamsters (the latter evaluating multiple adjuvants including CpG 1018) have been conducted using bench-scale vaccine. Preliminary data show that a 2-dose vaccine regimen (21 days apart) in BALB/c mice and golden Syrian hamsters induces potent protective immunity, as reflected by the level of anti-S IgG antibody (see Figure 2), prevention of weight-loss post-challenge with SARS-CoV-2 (hamsters, Figure 3), and the reduction of SARS-CoV-2 in the lung on Days 2 and 5 post-challenge (hamsters, Figure 3). This protective effect is particularly notable in hamsters, as they are known to develop COVID-19 lung disease much like humans.[50]

**Figure 2. Induction of anti-S after vaccination of mice (top) and hamsters (bottom) with NDV-HXP-S**

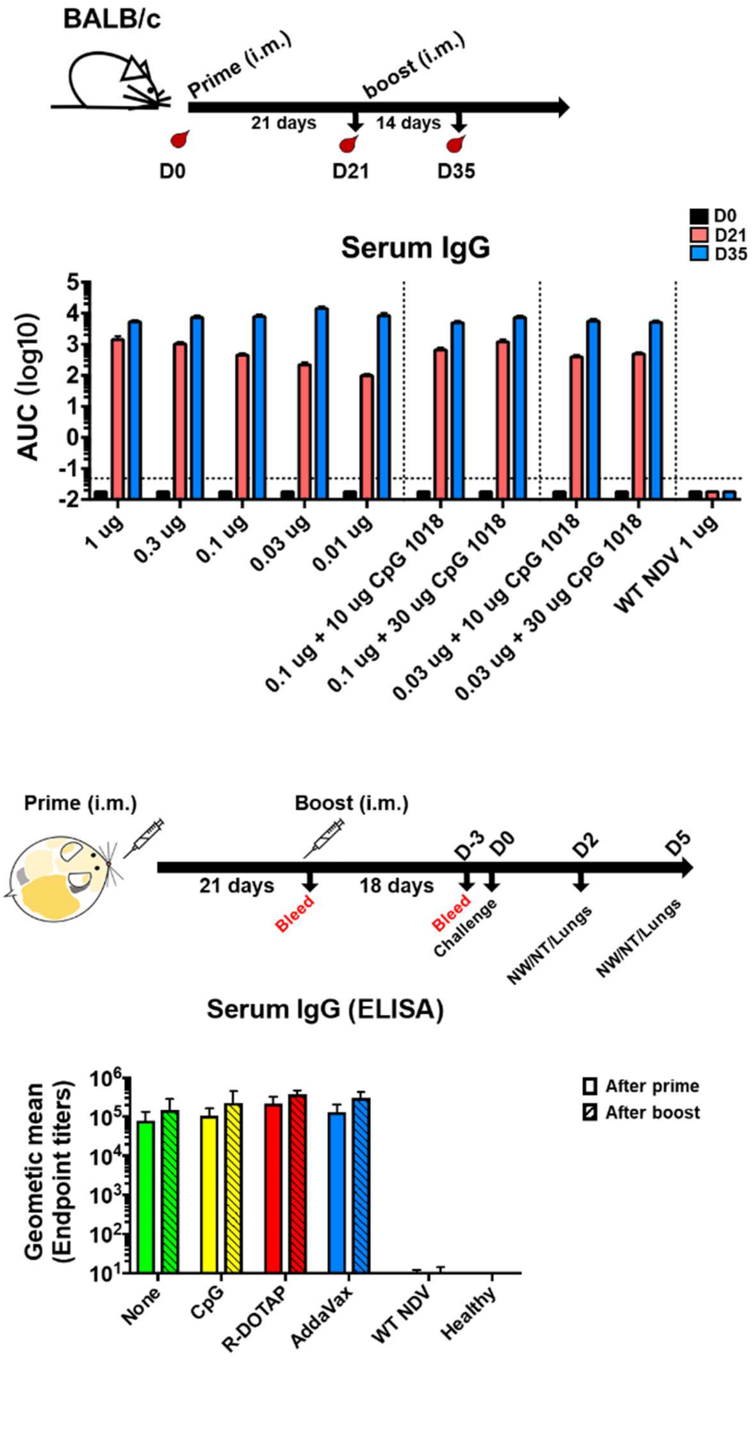

**Figure 3. Data from a pilot study showing protection of golden Syrian hamsters from infection by wild-type (WT) SARS-CoV-2.**

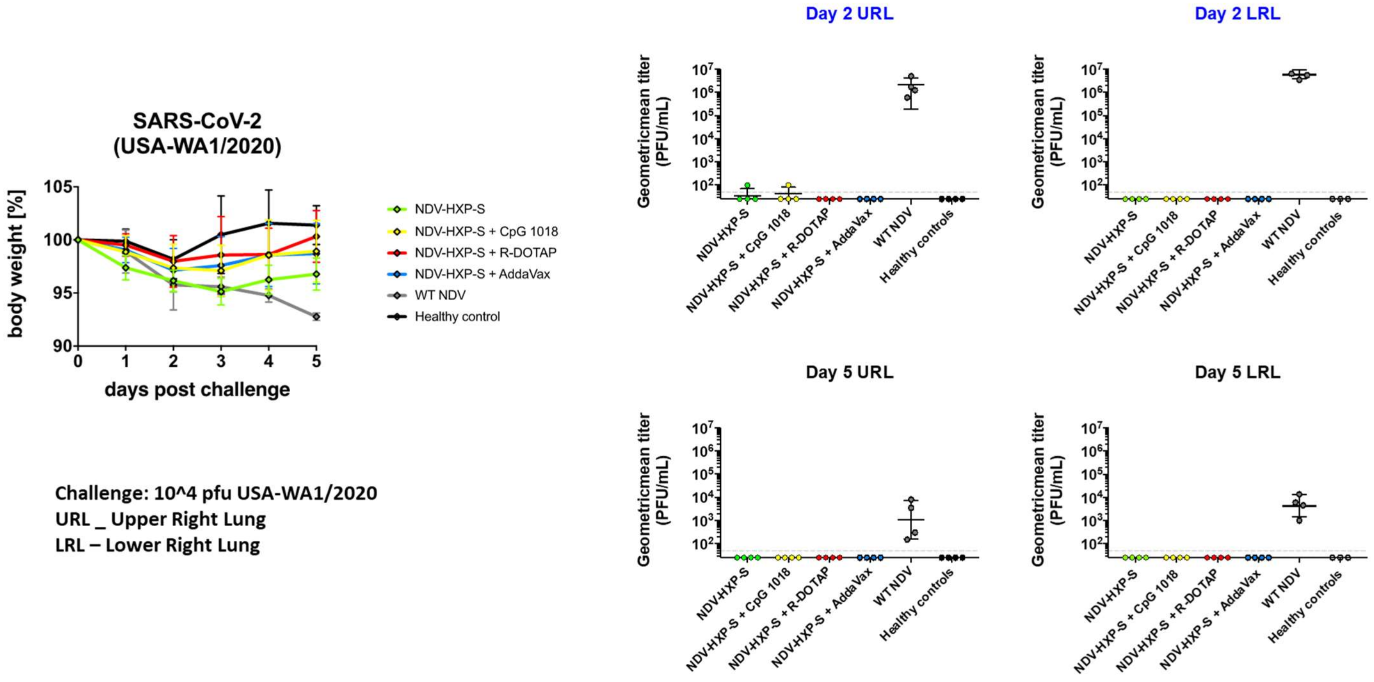

Additional preclinical studies (see Table 1) are described in brief below.

**Table 1. Summary of preclinical studies**

| Purpose | Species | Groups | Endpoints | Challenge virus | Institution |
|---|---|---|---|---|---|
| Cell-mediated immunity | Mice | NDV only (control)<br>NDV-HXP-S<br>NDV-HXP -S+CpG 1018 | Anti-CoV2 S ELISA<br>CD4, CD8 T cell, Th1, Th2 cytokine | None | ISMMS |
| Protection against challenge with WT SARS-CoV-2 | Hamsters | Mock challenge (control)<br>NDV only (control)<br>NDV-HXP-S<br>COVIVAC +CpG 1018 | Anti-CoV2 S ELISA<br>Weight loss x 5d, virus titer in lung | USA WA1/2020, $10^5$ pfu, IN (non-lethal) on D36 or D43 | ISMMS |
| GLP-Toxicology | Rat | Saline control<br>COVIVAC 10µg<br>COVIVAC 3µg+1.5mg CpG 1018 | Assess potential local and systemic toxicity of vaccine with and without CpG 1018 | None | Bioneeds India (GLP toxicology CRO, India) |

##### Evaluation of cell-mediated immunity in mouse study with GMP lot vaccine

Vaccine candidates that elicit strong T-cell responses with a Th1 bias may provide durable protection against COVID-19, as well as minimize risk of disease enhancement. To characterize the induction of cell-mediated immunity by COVIVAC vaccine with and without CpG 1018, each consortium manufacturer's vaccine will be evaluated in mice. Groups of mice (n=8) will be administered two doses of vaccine intramuscularly on day 0 and 21. Post dose 1 and 2, mice will be euthanized to isolate splenic T cells. T cells will be cultured overnight with peptide pools derived from S protein as the source of antigen stimulation, then immune-profiled to determine antigen specific CD4+ and CD8+ T-cell populations. The proportion of T cells secreting IFNγ or IL-4 cytokines will be estimated by ELISpot to assess Th polarization.

##### Comparing protective efficacy of GMP lots of vaccine in hamsters

A study is currently being conducted by ISMMS to evaluate the efficacy of each manufacturer's COVIVAC vaccine with and without CpG 1018 adjuvant against wild-type (WT) SARS-CoV-2 virus challenge in golden Syrian hamsters. Briefly, golden Syrian hamsters of 7-8 weeks of age (n = 8/group) will be immunized twice, three weeks apart using the GMP lot of COVIVAC vaccine from each manufacturer. A dose of 1 µg S antigen will be used, which corresponds to the low human dose planned for Phase 1. The adjuvanted vaccine group will also receive 100 µg of CpG 1018. The controls will be NDV vector only and mock challenged animals. Animals will be bled on Days 1, 21 and 33. Serum samples will be tested for anti-S IgG by ELISA. On Day 35, hamsters will be challenged with WT SARS-CoV-2. Body weight will be measured daily from all surviving animals. On Days 2 or 5 post-challenge, hamsters will be euthanized, and lung tissues collected to measure viral titers in the upper and lower lobes of the lung.

##### Repeat-dose toxicology study

A GLP non-clinical toxicology evaluation is being conducted to evaluate COVIVAC vaccine from each consortium manufacturer; interim results from this study have been reported, as detailed below. Per guidance provided by the World Health Organization (WHO), a repeat-dose toxicology study was designed to test COVIVAC (including CpG 1018) at the human dose level. Since COVIVAC will be evaluated in clinical trials as a two-dose vaccination schedule, the toxicology study evaluated IM administration of three doses, following the n+1 rule.

A total of 210 Sprague Dawley rats (105 Males and 105 Females) were randomly allocated to seven main groups (G) and seven recovery groups (R). Each main group consisted of 10 rats/sex and each recovery group consisted of 5 rats/sex. The animals allocated to group G1/G1R were administered 0.9% normal saline. COVIVAC was administered to the other groups as shown in Table 2.

**Table 2. COVIVAC Treatment Groups (Toxicology Study)**

| Antigen groups: | Groups | Dose |
|---|---|---|
| Antigen 1 (IVAC) | G2/G2R | COVIVAC 10 µg |
| | G5/G5R | COVIVAC 3 µg + CpG 1.5 mg |
| Antigen 2 (Instituto Butantan) | G3/G3R | COVIVAC 10 µg |
| | G6/G6R | COVIVAC (3 µg + CpG 1.5 mg |
| Antigen 3 (GPO) | G4/G4R | COVIVAC 10 µg |
| | G7/G7R | COVIVAC 3 µg + CpG 1.5 mg |

G = Group, R = Recovery group

Doses were administered IM in the thigh muscle of the animals on Days 1, 15 and 29. The total volume administered was 0.4 mL/animal, with 0.2 mL administered to each thigh. After treatment on Day 29, the main group animals (G1 to G7) were sacrificed on Day 32; the animals in the recovery groups (G1R to G7R) were not given any treatment and observed for reversibility or persistence of toxic effects, and sacrificed on Day 43. Animals were observed for clinical signs of toxicity twice daily, with detailed clinical examination weekly; local reactions at the site of injection were evaluated approximately 24, 48 and 72 hours after each dose. Rectal temperature was recorded pre-treatment and at approximately 6 hours (± 10 min) and 24 hours (± 20 min) after each dose on Days 1, 15 and 29. Body weight was recorded once on Day 1 prior to dosing, 3 hours post dose and on Day 2 and weekly thereafter. Feed consumption was measured daily. Blood was collected from all animals prior to treatment, on Day 3, 8 and at necropsy (Day 32 for main groups and Day 43 for recovery groups) from overnight-fasted animals. Urine was collected on Day 32 from main groups and on Day 43 from recovery groups and analysed. All animals were subjected to necropsy and detailed gross pathological examination at termination. Organs specified in the study plan were collected, weighed and preserved. Haptoglobin and alpha2-macroglobulin were analysed from main group animals (5 rats/sex) prior to treatment (pre-dose), on Days 8 and 32 using a kit-based ELISA method. Serum samples collected from all rats during pre-dose, Day 32 (main groups) and Day 43 (recovery groups) were assessed for immunogenicity.

Interim Results:

All three GMP lots of vaccine drug substance from GPO, IVAC and Butantan were well tolerated in rats with no morbidity/mortality found in any of the groups throughout the experimental period. There were no clinical signs of toxicity observed during the experimental period. A few statistically significant variations in percent change in body weight with respect to Day 1 were noted during the experimental period across groups that received any of the three vaccines; these were inconsistent and minimal in nature and hence considered as incidental. Also, there were no concomitant variations in mean body weights and/or feed consumption and hence these variations in body weight not considered to be related to vaccine administration.

No adverse treatment-related variations in body temperature after any dose were noted in groups of either sex. A few statistically significant variations of body temperature were noted during different measurement periods; however these were considered incidental as the variations were less than 2% and all the values were within 37±2°C. No skin reactions at the site of injection were noted after any administration of vaccine. Haptoglobin and alpha2-macroglobulin were increased on Day 8 and Day 32 when compared to pre-dose levels; this increase was also noted in the saline control group and hence considered not be related to treatment with vaccine. No adverse treatment-related changes were observed in hematology, coagulation, clinical chemistry and urinalysis parameters. Statistically significant changes were noted in hematology parameters on Days 3 and/or 8 when compared with pre dose sampling due to repeated blood withdrawal for clinical pathology analysis and hence not considered to be due to vaccine administration.

There were no adverse treatment-related changes in fasting body weights. However, in the main groups, statistically significant increases in absolute and relative organ weight of the liver (G5M, G6M, G5F, G6F and G7F), and decreases in relative weight of ovaries (G2F, G4F, G6F and G7F), heart (G5F, G6F) and brain (G6F) were noted. Further data from the recovery group animals and the histopathology data is necessary to interpret these data.

#### 2.8. Study Design Rationale

The Phase 1/2 combined study has been designed to rigorously evaluate the safety, tolerability (primary objective) and immunogenicity (secondary objective) of COVIVAC vaccine across a range of S-antigen dose levels – one with CpG 1018 as adjuvant – with the aim of expeditiously selecting a single candidate to advance to Phase 3 studies. Phase 1 will provide justification for advancement to the Phase 2 segment based on safety evaluations during an initial sentinel dose-escalation stage, and later an interim analysis of safety and immunogenicity (the latter primarily based on pseudovirus neutralization) through 28 and 14 days after the second dose, respectively. An independent Data and Safety Monitoring Board (DSMB) will review unblinded safety data through Day 57, and on the basis of this review provide a recommendation for advancing to Phase 2 (the DSMB will also be convened at any time during conduct of the study in the event that a study pause rule is triggered; see Section 9.3). In addition to this process for assessing adequate safety for advancing to Phase 2, a threshold immune response at Day 43 will be required for a vaccine formulation to be considered for advancement to Phase 2: namely, the observed seroresponse rate (defined as the percentage of subjects in a treatment group with at least a 4-fold rise from baseline in $NT_{80}$ titers) will need to be at least 52% at the lower limit (LL) of the 95% confidence interval (CI) . Based on this criterion, there is less than 10% chance of incorrectly rejecting a vaccine candidate with a true seroresponse rate of 85% or higher, which is estimated to be the minimum response rate that would be consistent with further investment in an additional second-wave COVID-19 vaccine.

If warranted based on safety and immunogenicity, the two optimal COVIVAC formulations will be selected for ongoing evaluation in Part 2 of the study based on results of the Phase 1 interim analysis. The Phase 2 segment of the study will include a sufficient number of older adults aged 60-75 (including those with stable chronic co-morbidities) to determine whether safety and immunogenicity of the vaccine may differ in this older population. It is critical that evaluation of COVIVAC in older adults informs the selection of the COVIVAC candidate to advance to Phase 3 studies (based on safety and immunogenicity through 14 days post-second vaccination in Phase 2), given the higher risk of severe COVID-19 in this age group, and the potential need for a higher dose level of antigen and/or use of adjuvant to achieve a comparable functional immune response as younger adults. Phase 2 immunogenicity and safety data through Day 43 and Day 57, respectively, will determine the vaccine composition to advance to Phase 3, registration, and commercialization. Cell-mediated immunity will also be evaluated in a subset of subjects in the Phase 2 segment as an exploratory objective to assess the S protein-specific T cell response including Th polarization. An exploratory evaluation of the IgG immune response against the NDV HN protein will also be conducted in all Phase 2 subjects to generate hypotheses about the utility of administering a booster dose of COVIVAC vaccine.

In addition to standard safety monitoring throughout the study, subjects will be evaluated for specific adverse event of special interest (AESI); these include potential immune-mediated medical conditions, which may be associated with vaccination. In addition subjects will be closely monitored for adverse events associated with COVID-19 to identify, and treat if indicated, potential cases of COVID-19 during the trial, and to assess for possible vaccine-induced immune enhancement of disease. In addition to mandatory use of masks and other exposure control measures by staff and subjects during clinic visits, study subjects will be instructed to contact the Principal Investigator (PI) if they meet criteria for COVID-19 testing based on specified signs and symptoms; these criteria will be provided to subjects in the form of a fact sheet.

##### 2.8.1. Justification for Dose

As shown above, the COVIVAC vaccine candidate elicits similar post-dose 2 titers of anti-S IgG antibody in mice over a 100-fold dilution (from 1 µg to 0.01 µg of total protein). The S protein of the vaccine content is estimated to be 10% of total protein. COVIVAC vaccine (5 µg total protein) administered twice to hamsters afforded excellent protection against virus challenge. These results suggest that relatively low doses of COVIVAC may be immunogenic in human subjects, despite the uncertainty of translating rodent doses to human doses. Consequently, 3 µg of S protein has been selected for unadjuvanted COVIVAC as the mid-dose range for clinical evaluation, with a high dose of 10 µg and a low dose of 1 µg. Evaluating this full dose range of COVIVAC in subjects aged 18-75 will provide the necessary safety and immunogenicity data to determine whether increasing the dose level of COVIVAC in elderly subjects in Phase 3 may be desirable.

Furthermore, the low dose level of COVIVAC vaccine will be evaluated ± CpG 1018 in the current study. CpG 1018 is approved for use in the US by the Food and Drug Administration (FDA) at a dose of 3 mg/ml in HEPLISAV-B. A dose of 1.5 mg/ml has been selected with COVIVAC for the following reasons:

- CpG 1018 has shown a dose-dependent adjuvant effect with other vaccines. Dynavax has shown that 1.5 mg/ml of CpG 1018 has an acceptable adjuvant effect with protein subunit vaccine.[51] Given that COVIVAC is an inactivated whole virion, it is expected that it will have intrinsic adjuvant effects that will complement the adjuvant effects of CpG 1018.
- Advanced adjuvants, such as CpG 1018, add considerable cost per dose of vaccine. Keeping the level of adjuvant relatively low but still effective, will assist in controlling the cost of

goods of the final product.

- CpG 1018 is being tested in multiple vaccine candidates. This may put significant pressure on the supply/availability of CpG 1018 for approved products. Formulating COVIVAC with a lower dose of CpG 1018 may be beneficial when manufacturing at commercial scale.

##### 2.9. Overall Clinical Development Strategy

The ultimate aim of developing COVIVAC vaccine for the prevention of COVID-19 is to obtain local registration of the vaccine in Vietnam, followed by prequalification by the WHO, with the goal of providing affordable access to a life-saving COVID-19 vaccine for Vietnam and other LMICs. Assuming this Phase 1/2 study results in advancement of an COVIVAC vaccine candidate for further evaluation, IVAC will likely conduct one Phase 3 trial to confirm lot-to-lot consistency and generate expanded safety data for that candidate. The nature of a second Phase 3 study to confirm vaccine effectiveness will be determined in discussions with the MoH of Vietnam. As COVID-19 vaccines become available and SARS-CoV-2 transmission rates decline due to population immunity and public health control measures, conducting placebo-controlled studies may be challenging if not infeasible. As a result, it may be the case that effectiveness can only be demonstrated by conducting an immunogenic non-inferiority trial that includes an approved comparator with a similar mechanism of protection.

#### 3. HYPOTHESES, OBJECTIVES AND ENDPOINTS

##### 3.1. Study Hypotheses

###### 3.1.1. Primary Hypothesis:

**Safety, Tolerability:**

- The COVIVAC vaccine (NDV-HXP-S) will have an acceptable safety profile and be well-tolerated at all dose levels when administered as a two-dose regimen either without adjuvant (at 1 µg, 3 µg, and 10 µg of S protein per 0.5 ml dose), or with the adjuvant CpG 1018 (at 1 µg of S protein per 0.5 ml dose).

###### 3.1.2. Secondary Hypothesis:

**Immunogenicity:**

- COVIVAC will elicit a measurable and dose-dependent neutralizing antibody response.

##### 3.2. Study Objectives

###### 3.2.1. Primary Objectives:

**Safety, Tolerability:**

**For Phase 1:**

1. To assess the safety and tolerability of each of four formulations of COVIVAC (at 1, 3 and 10 µg without adjuvant; and at 1 µg with CpG 1018) after the first and second dose of a two-dose regimen administered at a 28-day interval to adults aged 18-59 years

**For Phase 2:**

1. To assess the safety and tolerability of two selected formulations of COVIVAC (based on Phase 1 safety and immunogenicity) after the first and second dose of a two-dose regimen administered at a 28-day interval to adults aged 18-75 years
2. To assess the safety and tolerability of two selected formulations of COVIVAC after the first and second dose of a two-dose regimen administered at a 28-day interval to adults aged 18-59 years and 60-75 years

##### 3.2.2. Secondary Objectives:

**Immunogenicity:**

**For Phase 1:**

1. To assess the functional (neutralizing) humoral immune response elicited by each formulation of NDV-HXP-S, as measured by a SARS-CoV-2 PNA, at baseline, 28 days after the first vaccination, and 14 days after the second vaccination (to inform the selection of two formulations for evaluation in Phase 2), and at 6 months and 12 months after the second vaccination, in adults aged 18-59 years
2. To assess the IgG immune response elicited by each formulation of COVIVAC against the S protein of SARS-CoV-2, as measured by ELISA, at baseline, 28 days after the first vaccination, and 14 days, 6 months and 12 months after the second vaccination, in adults aged 18-59 years

**For Phase 2:**

1. To assess the functional (neutralizing) humoral immune response elicited by each of two selected COVIVAC formulations, as measured by PNA, at baseline, 28 days after the first vaccination, and 14 days, 6 months and 12 months after the second vaccination, in adults aged 18-75 years
2. To assess the functional (neutralizing) humoral immune response elicited by each of two selected COVIVAC formulations, as measured by PNA, at baseline, 28 days after the first vaccination, and 14 days, 6 months and 12 months after the second vaccination, in adults aged 18-59 years and 60-75 years
3. To assess the IgG immune response elicited by each of two selected COVIVAC formulations against the S protein of SARS-CoV-2, as measured by ELISA, at baseline, 28 days after the first vaccination, and 14 days, 6 months and 12 months after the second vaccination, in adults aged 18-75 years
4. To assess the IgG immune response elicited by each of two selected COVIVAC formulations against the S protein of SARS-CoV-2, as measured by ELISA, at baseline, 28 days after the first vaccination, and 14 days, 6 months and 12 months after the second vaccination, in adults aged 18-59 years and 60-75 years

##### 3.2.3. Exploratory Objectives:

**Immunogenicity:**

**For Phase 2:**

1. To assess the S protein-specific T cell response elicited by each of two selected COVIVAC formulations, as measured by ELISpot, at baseline, and 14 days and 6 months after the second vaccination, in adults aged 18-75 (subset of subjects)
2. To assess the S protein-specific T cell response elicited by each of two selected COVIVAC formulations, as measured by ELISpot, at baseline, and 14 days and 6 months after the second vaccination, in adults aged 18-59 years and 60-75 years (subset of subjects in each age stratum)
3. To assess the IgG immune response elicited by each of two selected COVIVAC formulations against the NDV HN protein, as measured by ELISA, at baseline, 28 days after the first vaccination, and 14 days and 12 months after the second vaccination, in adults aged 18-59 years

*Note: other exploratory studies to further characterize the immune response to COVIVAC may be conducted in Phase 1 or Phase 2. This includes evaluation of the functional response to COVIVAC as measured by a wild-type neutralization assay, which will be conducted if a collaboration for performing the assay is established. The collaborator for evaluation of the cell-mediated immune response to COVIVAC in Phase 2 is also yet to be determined.*

#### 3.3. Study Endpoints

##### 3.3.1. Primary Endpoints:

**Safety, Tolerability:**

- Number and severity of solicited local and systemic AEs during the first 7 days after each vaccination
- Number, severity, and relatedness of clinically significant hematological and biochemical measurements at 7 days post each vaccination
- Number, severity and relatedness of all unsolicited AEs during the first 28 days after each vaccination
- Number, severity and relatedness of SAEs throughout the entire study period
- Number, severity and relatedness of MAAEs throughout the entire study period
- Number, severity and relatedness of AESI throughout the entire study period, including AESI relevant to COVID-19, and potential immune-mediated medical conditions (PIMMC)

##### 3.3.2. Secondary Endpoints:

**Immunogenicity:**

**For Secondary Objective 1 (Phase 1), and Objectives 1 and 2 (Phase 2):**

- $NT_{50}$ and $NT_{80}$ GMT against SARS-CoV-2 pseudovirus at 28 days after the first vaccination, and 14 days, 6 months and 12 months after the second vaccination in subjects who are anti-S IgG seronegative at baseline

- GMFR (from baseline) in $NT_{50}$ and $NT_{80}$ against SARS-CoV-2 pseudovirus at 28 days after the first vaccination, and 14 days, 6 months and 12 months after the second vaccination
- Percentage of subjects with $NT_{50}$ and $NT_{80}$ seroresponses against SARS-CoV-2 pseudovirus as defined by (1) a ≥ 4-fold increase from baseline, and (2) a ≥ 10-fold increase from baseline at 28 days after the first vaccination, and 14 days, 6 months and 12 months after the second vaccination

**For Secondary Objective 2 (Phase 1), and Objectives 3 and 4 (Phase 2):**

- Anti-S IgG GMT at 28 days after the first vaccination, and 14 days, 6 months and 12 months after the second vaccination in subjects who are anti-S IgG seronegative at baseline
- GMFR (from baseline) in anti-S IgG GMT at 28 days after the first vaccination, and 14 days, 6 months and 12 months after the second vaccination
- Percentage of subjects with seroresponses in anti-S IgG titer as defined by (1) a ≥ 4-fold increase from baseline, and (2) a ≥ 10-fold increase from baseline, at 28 days after the first vaccination, and 14 days, 6 months and 12 months after the second vaccination

##### 3.3.3. Exploratory Endpoints:

**Immunogenicity**

**For Exploratory Objectives 1 and 2 (Phase 2):**

- Magnitude, functionality, and Th polarization of S protein-specific T cells relative to baseline at 14 days and 6 months after the second vaccination

**For Exploratory Objective 3 (Phase 2):**

Anti-NDV HN IgG GMT at baseline, 28 days after the first vaccination, and 14 days and 6 months after the second vaccination.

*Note: In this protocol one month in this study context is defined as 4 weeks or 28 days; then 6 months means 24 weeks or 168 days, and 12 months means 48 weeks or 336 days.*

#### 4. STUDY DESIGN

This prospective, single-center, randomized, placebo-controlled, observer-blind Phase 1/2 study includes two separate parts. The treatment groups to be evaluated in each part of the study (and their corresponding sample size) are presented in Table 3.

**Table 3. Treatment Groups**

| Phase | Group | Sample Size | Study Product |
|---|---|---|---|
| 1 | 1 | 20 | Placebo |
| | 2 | 25 | COVIVAC 1 µg |
| | 3 | 25 | COVIVAC 3 µg |
| | 4 | 25 | COVIVAC 10 ug |
| | 5 | 25 | COVIVAC 1 µg + CpG1018 1.5 mg |
| 2 | 1 | 50 | Placebo |
| | 2 | 125 | Formulation A |
| | 3 | 125 | Formulation B |

Part 1 is a first-in-human, Phase 1 study designed to evaluate the safety, tolerability, and immunogenicity of the COVIVAC vaccine at three different dose levels (1, 3, and 10 µg) without adjuvant, and at one dose level (1 µg) with the adjuvant CpG 1018, in a total of 120 subjects aged 18-59 years. Eligible subjects will be randomly assigned to receive study vaccine or placebo as an IM injection in the deltoid muscle at Visit [V]1 as part of one of four sequential groups of escalating antigen dose level or added adjuvant. Each group will be divided into two sequential subgroups, a sentinel subgroup (n=6) and a larger subgroup (n=24) that includes the remaining subjects to be evaluated at that dose level (or for that formulation in the case of the one adjuvanted treatment group). Initially, a sentinel subgroup of 6 subjects will be randomized to receive COVIVAC 1 µg or placebo in a 5:1 ratio (subjects within a sentinel subgroup will receive the first injection only after immediate reactogenicity assessments of the prior subject in the subgroup are completed). Sentinel subjects will be monitored for reactogenicity and safety through Day 8 (Visit 2), including for the initial 24 hours post-injection in clinic. If the PSRT has no safety concerns based on blinded review of all safety data, including clinical laboratory results, through Visit 2, the second subgroup of 24 subjects will be randomized to receive COVIVAC 1 µg or placebo at the same ratio; on the same day, the sentinel subgroup (n=6) for the second dose level to be evaluated (COVIVAC 3 µg) can be randomized to receive study vaccine or placebo in a 5:1 ratio, and so on for the subsequent dose level and formulation (see Table 4).

**Table 4. Phase 1 Dose Escalation Steps**

| Subgroup | Escalation Steps | | | | | | | | | | | | | |
|---|---|---|---|---|---|---|---|---|---|---|---|---|---|---|
| Dose 1µg & Placebo (5+1) | V1 | V2 | | | | | | | | | | | | |
| Dose 1µg & Placebo (20+4) | | | | V1 | V2 | | | | | | | | | |
| Dose 3µg & Placebo (5+1) | | | | V1 | V2 | | | | | | | | | |
| Dose 3µg & Placebo (20+4) | | | | | | | V1 | V2 | | | | | | |
| Dose 10µg & Placebo (5+1) | | | | | | | V1 | V2 | | | | | | |
| Dose 10µg & Placebo (20+4) | | | | | | | | | | V1 | V2 | | | |
| Dose 1µg+CpG & Placebo (5+1) | | | | | | | | | | V1 | V2 | | | |
| Dose 1µg+CpG & Placebo (20+4) | | | | | | | | | | | | | V1 | V2 |
| **Review** | | | PSRT | | | PSRT | | | PSRT | | | PSRT | | |

V1 = Visit 1 (Day 1)
V2 = Visit 2 (Day 8)
PSRT = Protocol Safety Review Team meeting

The PSRT will review blinded safety, reactogenicity and clinical laboratory data accrued on a weekly basis until D43 of the last subject in each phase of the study, then at least once monthly until the end of the study. Subjects will receive 2 doses of assigned IP on D1 and D29 (V1 and V3), and be assessed in clinic for safety and reactogenicity at 7 days after each vaccination (V2 and V4), for safety at 28 days after the second vaccination (V6), and for safety and immunogenicity assessments at baseline (V1), 28 days after the first vaccination (V3) and 14, 168 and 336 days after the second vaccination (V5, V7, and V8, EOS Visit).

An interim analysis of Phase 1 data conducted after the last subject last visit for V6 (D57) will serve as the basis for decisions about down-selection and advancing to Part 2 of the study (Phase 2). Down selection and advancement to Part 2 (Phase 2) will be based on the following parameters:

- Post-dose 2 immunogenicity results at the aggregate treatment level
  - A threshold immune response at Day 43 will be required: the observed seroresponse rate in a treatment group (defined as the percentage of subjects with at least a 4-fold rise from baseline in 80% neutralizing antibody titers) will need to be ≥52% at the LL of the 95% CI for that treatment (vaccine formulation) to be considered for advancement to Phase 2.
- Post-dose 1 and post dose 2 safety results including all solicited and unsolicited adverse events, serious adverse events, and clinical laboratory results.

The following process will be followed for the decision about down selection and advancing Part 2 (Phase 2):

- The DSMB will review the unblinded safety data and provide a recommendation to the Sponsor on whether the safety profile is acceptable for advancing a formulation to Phase 2.
- The Sponsor will review the DSMB recommendation in conjunction with the immunogenicity data and select two formulations to advance to Phase 2.

- If multiple formulations achieve the threshold immune response (as well as have an adequate safety and tolerability profile per the DSMB), the Sponsor will select two formulations to advance to Phase 2 based on consideration of such factors as the relative functional immunogenicity of these formulations, opportunity for dose sparing, and opportunity to limit cost and possible supply constraints associated with use of the CpG adjuvant.

- The selection and recommendation to advance to Phase 2 along with the interim report will be jointly reviewed by NIHE's IRB and MoH prior to Phase 2 enrollment.

In Part 2 of this combined Phase 1/2 study, 300 adults aged 18-75 years will be randomized (2:5:5) to placebo, or one of two selected formulations of COVIVAC being evaluated in Phase 1. At least one-third of the subjects in Phase 2 will be aged ≥60 years to ensure that adequate safety and immune data will be available from older and elderly adults to inform the selection of the COVIVAC formulation to advance to Phase 3 studies. The Phase 2 cohort will follow the same visit schedule, and undergo the same procedures and assessments, as in Phase 1. In addition, as exploratory objectives, the anti-NDV HN IgG response will be assessed at V1, V3, V5, and V8 in all subjects, and 36 subjects (equally distributed between the two age strata) will be randomly selected in a 1:1:1 ratio to provide additional blood at V1, V5 and V7 to be used to isolate PBMCs for assessment of T-cell-mediated immunity. Other exploratory assays to characterize the immune response to COVIVAC may also be performed.

An interim analysis of Phase 2 data (combined with immunogenicity and safety data generated for the placebo group and two selected COVIVAC formulations from Phase 1) will be conducted after the last subject of the Phase 2 cohort completes V6 (D57) as the basis for selecting the optimal formulation of COVIVAC to advance to Phase 3 studies. As was the case for the Phase 1 interim analysis at the same timepoint, the data generated will include unblinded post-dose 1 and dose 2 safety results for review by the DSMB, and immunogenicity results aggregated by treatment group for review by the Sponsor. The DSMB will consider all accumulated safety data from both phases of the study prior to making any recommendation to the Sponsor that it not advance a formulation based on safety concerns. The Sponsor will ultimately select the formulation to advance to Phase 3 that, in addition to having been judged by the DSMB to have an adequate safety and tolerability profile, is optimal based on relative functional immunogenicity and other programmatic considerations such as those noted above.

### 5. STUDY POPULATION

#### 5.1. Study Population

The study population will consist of eligible, Vietnamese male and female adults aged 18-75 years old, inclusive.

#### 5.2. Clinical Trial Site

- Phase 1:

  Phase 1 of the study will be conducted at Hanoi Medical University (HMU), where facilities are available to conduct intensive in-clinic safety monitoring for 24 hours after the 1st study product administration, and for 4 hours after the 2nd study product administration. Hanoi Medical University is experienced in the conduct of clinical trials. Study subjects in Phase 1 will be recruited via advertisement in mass media, and websites of NIHE, HMU and IVAC; volunteers will contact the study team for registration. The advertisement materials will be reviewed by relevant Ethics Committees before going live.

As of 6 January 2021, Hanoi has recorded 198 COVID-19 cases. For approximately the last 4 months, there has been no case of COVID-19 in Vietnam resulting from community transmission; new cases have been imported from outside of Vietnam and quarantined upon arrival. As a result exposure to SARS-CoV-2 is likely to be limited during conduct of the study, thus limiting censoring of subjects from immunogenicity analyses requiring baseline seronegativity.

- Phase 2:

Phase 2 of the study will be conducted at Vu Thu district health center, Thai Binh province in Vietnam. National Institute of Hygiene and Epidemiology (NIHE) will collaborate with staff of the commune health centers, district health center, district hospital, as well as the staff of the Provincial Center for Disease Control for conduct of this study. Thai Binh CDC and district health center have collaborated with NIHE for several vaccine clinical trials funded by international sponsors and monitored by contract research organizations (CROs).

According to the health system in Vietnam, the district hospitals and their corresponding district health centers, the provincial general hospital, and the CDC are all managed by the Provincial Health Services; so it is straightforward to coordinate care among them should a serious adverse event occur. The staff of commune and district health centers conduct routine vaccinations in the community and have the capacity to handle serious vaccine reactions. The district hospitals also have good capacity in terms of both personnel and equipment to provide medical care, including emergency care. The district health center is only approximately 7 kilometers (10 minute drive) from provincial general hospitals that would be engaged to manage higher grade and serious adverse events requiring hospitalization or referred events requiring consultation with a specialist. This will assure rapid patient transport and referral to tertiary care hospitals at provincial levels in case such referral is necessary. The study documents will be stored at the provincial CDC where data entry will occur, and will be moved to the district health center when a study visit is occurring there. The study samples will also be stored at the provincial CDC lab. The study team will recruit subjects for this study from the communes around the Vu Thu district health center. Subjects will be seen at the Vu Thu district health center for scheduled study visits, and receive routine medical care at Vu Thu district hospital. Subjects will be encouraged to contact the study team if they have any new onset medical conditions, medically attended events, notable signs and/or symptoms of COVID-19, or other concerns during their period of enrollment in the study to ensure all safety data are collected in a complete, accurate and timely manner.

As of 6 January 2020, there has been no confirmed case of COVID-19 in the Vu Thu district. In daily life, people living in Vu Thu district do not often have contact with people outside of their community, as travel outside the district – and migration into the district – is rare. This is expected to facilitate compliance with study visits and safety monitoring generally. Also, exposure to SARS-CoV-2 is likely to be very limited during conduct of the study, thus limiting censoring of subjects from immunogenicity analyses requiring baseline seronegativity.

Regardless of estimated exposure risk, HMU and Vu Thu district health center will follow the policies and procedures implemented by Ministry of Health to minimize risk of SARS-CoV-2 transmission at these facilities, including mandatory use of masks by all staff, social distancing, and frequent handwashing/sanitizing. This policy will be extended to include study subjects (and prospective subjects) during clinic visits, and will be strictly enforced. Study subjects will also be instructed on how to minimize risk of exposure to SARS-CoV-2 while outside of the study clinic, including during transit to and from study visits at HMU and Vu Thu district health center.

Prospective subjects who are consented to participate (i.e., enrolled) in this Phase 1/2 study will only be eligible for randomization if, during screening, they meet all of the inclusion criteria, and none of the exclusion criteria, as follows:

#### 5.3. Inclusion Criteria

**Phase 1 Only:**

1. Adult 18 through 59 years of age inclusive at the time of randomization.
2. Healthy, as defined by absence of clinically significant medical condition, either acute or chronic, as determined by medical history, physical examination, screening laboratory test results, and clinical assessment of the investigator.

**Phase 2 Only:**

1. Adult 18 through 75 years of age inclusive at the time of randomization.
2. Having no clinically significant acute medical condition, and no chronic medical condition that has not been controlled within 90 days of randomization, as determined by medical history, physical examination, screening laboratory test results, and clinical assessment of the investigator.

**Both Phase 1 and Phase 2:**

3. Has provided written informed consent prior to performance of any study-specific procedure.
4. Has a body mass index (BMI) of 17 to 40 $kg/m^2$, inclusive, at screening.
5. Resides in study site area and is able and willing to adhere to all protocol visits and procedures.
6. If a woman is of childbearing potential, must not be breastfeeding or be pregnant (based on a negative urine pregnancy test at screening and during the 24 hours prior to receipt of the first dose of IP), must plan to avoid pregnancy for at least 28 days after the last dose of IP, and be willing to use an adequate method of contraception consistently and have a repeated pregnancy test prior to the second (last) dose of IP.

#### 5.4. Exclusion Criteria

1. Use of any investigational medicinal product within 90 days prior to randomization or planned use of such a product during the period of study participation.
2. History of administration of any non-study vaccine within 28 days prior to administration of study vaccine or planned vaccination within 3 months after enrolment.

   Note: receipt of any COVID-19 vaccine that is licensed or granted Emergency Use Authorization in Vietnam during the course of study participation is not exclusionary if administered after Visit 5.
3. Previous receipt of investigational vaccine for SARS or MERS, or any investigational or licensed vaccine that may have an impact on interpretation of the trial results

4. History of hypersensitivity reaction to any prior vaccination or known hypersensitivity to any component of the study vaccine
5. History of egg or chicken allergy
6. History of angioedema
7. History of anaphylaxis
8. Acute illness (moderate or severe) and/or fever (body temperature measured orally $\geq 38^{\circ}C$)
9. Any abnormal vital sign deemed clinically relevant by the PI
10. Abnormality in screening laboratory test deemed exclusionary by the PI in consultation with the Sponsor
11. A positive serologic test for hepatitis B (HBsAg) or hepatitis C (HCV Ab)
12. History of confirmed HIV
13. History of laboratory-confirmed COVID-19
14. History of malignancy, excluding non-melanoma skin and cervical carcinoma in situ
15. Any confirmed or suspected immunosuppressive or immunodeficient state
16. Administration of immunoglobulin or any blood product within 90 days prior to first study injection or planned administration during the study period.
17. Administration of any long-acting immune-modifying drugs (e.g., infliximab or rituximab) or the chronic administration (defined as more than 14 days) of immunosuppressants within six months prior to first study injection, or planned administration during the study period (includes systemic corticosteroids at doses equivalent to ≥ 0.5 mg/kg/day of prednisone; the use of topical steroids including inhaled and intranasal steroids is permitted).
18. History of known disturbance of coagulation or blood disorder that could cause anemia or excess bleeding. (e.g, thalassemia, coagulation factor deficiencies).
19. Recent history (within the past year) or signs of alcohol or substance abuse.
20. Any medical, psychiatric or behavior condition that in the opinion of the PI may interfere with the study objectives, pose a risk to the subject, or prevent the subject from completing the study follow-up.
21. Employee of any person employed by the Sponsor, the contract research organization (CRO), the PI, study site personnel, or site.

Note: specific exclusion criteria (e.g., ≥Grade 2 acute illness, or abnormal vital sign deemed clinically relevant by the PI/designee) will be reassessed at both vaccination visits. Any subject who cannot be vaccinated due to an acute abnormality assessed at a vaccination visit (Visit 1 or Visit 3) may return once the acute issue has resolved, if deemed appropriate by the PI/designee. A minimum of 48 hours must have passed after a documented fever before a subject can be vaccinated. This safety requirement will not be deemed a protocol deviation should the visit fall outside the vaccination window; however, it will be encouraged to maintain the vaccination window whenever possible in these situations. Clinical laboratory test results and vital signs used to determine subject eligibility will be those obtained at screening. These tests may be repeated once if deemed appropriate by the investigator and determined to be due to a transient condition that has resolved. In addition, a test may also be repeated for test results determined to be spurious by the investigator (e.g., following improper specimen collection). The last measurement will be taken as the baseline for purposes of analysis.

##### 5.5. Screen Failures

Prospective subjects who are enrolled in the study, and who subsequently do not meet one or more criteria required for participation in the study during the screening period, are considered screen failures. Any individual deemed a screen failure may not be rescreened. All screen failures will be recorded in the Case Report Form (CRF). Any individual deemed a screen failure due to a previously undiagnosed medical condition will be provided with appropriate referral within the health care system.

#### 6. STUDY PRODUCT

##### 6.1. Product Descriptions

COVIVAC is an inactivated NDV chimera expressing a trimeric pre-fusion form of the SARS-CoV-2 S protein that contains six proline mutations (HexaPro). The vaccine is a clear and slightly opalescent liquid.

All study vaccine products to be administered on a given day (based on the study stage) will be formulated separately by the assigned site staff under aseptic conditions at the start of the clinic day. A detailed mixing procedure will be provided in the Pharmacy Manual or SOP for study product preparation. Each vaccine product will be formulated by filling a 4 ml (4R) [with the height of 46 mm and diameter of 16/1 mm] USP Type I glass vial with bulk drug substance, bulk adjuvant (if indicated based on formulation) and phosphate-buffered saline (PBS) to allow for five 0.5 ml doses per vial, as well as sufficient residual volume for possible dose verification. Each vial label will include the following information: name of the medicinal product, composition, fill volume, route of administration, lot number, manufacturing date, storage condition, and a cautionary statement ("For Clinical Trial Use Only").

The placebo control is PBS (pH 7.2), manufactured by IVAC and tested by National Institute for Control of Vaccines and Biological (NICBV).

##### 6.2. Stability and Storage

The temperature of bulk COVIVAC drug substance and CpG 1018 will be monitored during shipment to and storage at the clinical site to ensure that temperature deviations do not occur. The temperature of shipments will be monitored throughout transit using a continuous temperature monitoring system. Vaccine and adjuvant will not be used until their temperature throughout transit has been confirmed to be within acceptable limits. Upon receipt at HMU (for Phase 1) and the Thai Binh CDC (for Phase 2), bulk COVIVAC vaccine, CpG 1018 adjuvant, and PBS will be stored at 2°C to 8°C in a dedicated refrigerator that is safe, locked, and not accessible to unauthorized staff – including study team personnel blinded for study conduct purposes. The refrigerator will be under continuous temperature monitoring with maintenance of daily temperature logs and connected to a power source with a reliable back-up system. COVIVAC vaccine, CpG 1018 and PBS must not be frozen.

It is the responsibility of designated unblinded site staff to ensure that study products have not been exposed to temperatures outside the allowed range during transport or storage at HMU and Thai Binh CDC prior to being dispensed for vaccination. Should there be a deviation outside the allowed temperature range, the affected vaccine will be quarantined. The temperature deviation will be reported to the Sponsor and CRO within 24 hours of awareness who will advise the unblinded investigator team of the action to be taken based on the magnitude and duration of the temperature deviation. All drug accountability procedures will be documented and are the responsibility of the unblinded study staff.

#### 6.3. Preparation and Administration

A limited number of appropriately trained, unblinded study staff, including the site pharmacist, will be responsible for preparing study products (in accordance with the randomly determined assignment), administering the study vaccine, and handling all drug accountability procedures. These personnel will not participate in the other aspects of the clinical trial, to help ensure the integrity of the blind at the site. The unblinded staff will not reveal subjects' randomization assignments to subjects, or staff associated with the Sponsor, CRO, or site.

Unblinded staff will retrieve a subject's randomization assignment via secure Interactive Web Response System (IWRS) or alternative approach (randomization envelope) after being informed by the PI or designee that a subject is eligible for randomization. They will prepare the 0.5 ml dose of study product based on the subject's randomization assignment in a setting distinct from the clinic staff. Since COVIVAC vaccine may appear opalescent, the syringe barrels of study product will be masked to maintain the observer-blind study design. A total of five doses of study product may be drawn from each vial of formulated study vaccine prepared for clinic on the same day; however, in no circumstance may a dose be prepared and administered more than 6 hours after a vial of study product has been formulated (the vials will be stored between 2°C and 8°C during this interval). To be prepared for the highest level of possible medical risk, vaccination will take place in a clinical setting in which there is immediate access to the medical personnel, equipment, and medications required for emergency resuscitation.

Since COVIVAC vaccine is a suspension, study vaccine must be shaken gently immediately prior to use, in order to obtain a uniform homogenous suspension (clear to opalescent in color). Inspection of each vial will occur immediately prior to use. If a vial or its contents appear unusual in any way, the vial will not be used, and procedures detailed in the Pharmacy Manual for documentation and disposal will be followed.

COVIVAC vaccine or PBS placebo will be administered as an IM injection into the mid-deltoid muscle of the subject's non-dominant arm. Should the non-dominant arm not be acceptable based on clinical assessment (e.g., local infection or pre-existing swelling) the dominant arm may be used and the reason for the change documented. The site at which study vaccine is administered will be documented in the CRF for all subjects. A detailed account of procedures related to preparation and administration of study vaccine will be included in the Pharmacy Manual or SOP for study product preparation.

#### 6.4. Accountability and Disposal

The site pharmacist is required to maintain complete records of all study products received from the Sponsor and will be responsible for maintaining an accurate record of the randomization codes, inventory, and an accountability record of vaccine and placebo supplies for the study. The site pharmacist will also be responsible for ensuring the security of these documents. All used vials of study product will be stored in a dedicated space that is accessible only to the unblinded site staff and the unblinded CRO monitor, who is responsible for the monitoring and final accountability of all study products. Partially used vials after the completion of a clinic day will not be used for human administration or for any other investigational purpose. After final drug accountability is completed, all used and unused study products must be disposed of according to the Sponsor's instructions. Due to the need to maintain blinding, no drug accountability records will be sent to the Sponsor or included in the trial master file (TMF) until after database lock.

### 7. STUDY PROCEDURES

#### 7.1. Recruitment

Details on recruitment and retention of subjects will be outlined in a Recruitment and Retention Plan prior to study initiation.

In Phase 1 of the study, subjects will be recruited via advertisement in mass media, and websites of NIHE, HMU and IVAC; volunteers will contact study team for registration. The advertisement materials will be reviewed by relevant Ethics Committees before going live. Potential subjects who are interested in study participation will be invited to Hanoi Medical University to attend an ICF meeting and then a one-to-one consultation with the investigator to address questions and concerns before being consented and signing the ICF.

In Phase 2 of the study, the study staff will work together with the appropriate district health and commune health staff to recruit study participants, using IRB-approved recruitment strategies.

The commune staff will work with population collaborators/village health workers to develop a list of potential study participants based on the existing Village Population Register Books. After that they will go household to household to introduce the study and eligibility criteria to potential study participants using information from the "Information Sheet" of the consent document.

Once a potential study participant confirms their interest, they will be invited to Vu Thu District Hospital to attend an ICF meeting and participate in screening if they are consented.

At the ICF meeting, there will be an information session presented by the PI/designee to provide all necessary information related to the study. After the information session, each potential participant will have a one-and-one meeting with one of the assigned study investigators to further discuss the trial and ask any questions they may have. The investigator will assess the potential participant's understanding of key aspects of the study, such as the voluntary nature of study participation and potential risks and benefits, using an assessment of understanding. The investigator will review with the participant any answers they did not understand in order to clarify any misunderstandings. Only after that, if participant provided consent and signed the ICF, can the screening procedures be started.

The study staff will follow inclusion/exclusion criteria to determine eligibility. Screening will be conducted on all people who have signed the study consent, and their results will be reviewed with them, regardless of eligibility.

##### 7.1.1. Initial and Continuing Informed Consent

Informed consent is the process of ensuring that study subjects fully understand the purpose of the study and what will and may happen to them while participating in the study. Initial written informed consent is required before any study procedures are performed. The informed consent process continues throughout the study. If any new information becomes available that, in the judgement of the PI or Sponsor, may affect subjects' decision to continue in the trial, such information will be shared with subjects, who may be asked to sign a new consent form.

#### 7.2. Study Visits

Procedures to be performed at each study visit are shown in Table 5 as follows:

**Table 5. Schedule of Study Visits and Procedures – Phase 1 and Phase 2**

| VISIT | Screen | V1 | V2 | V3 | V4 | V5 | V6 | V7 | V8 |
|---|---|---|---|---|---|---|---|---|---|
| Study Day (allowed window in days) | **-28 to 1** | **1** | **8 (+3)** | **29 (+7)** | **36 (+3)** | **43 (+3)** | **57 (+7)** | **197 (+14)** | **365 (+28)** |
| Informed consent | ✓ | | | | | | | | |
| Demographics | ✓ | | | | | | | | |
| Medical History | ✓ | | | | | | | | |
| Concomitant medications[A] | ✓ | ✓ | ✓ | ✓ | ✓ | ✓ | ✓ | ✓ | ✓ |
| Eligibility check | ✓ | | | ~ | | | | | |
| Vital signs | ✓ | ✓^✓ | ✓ | ✓^✓ | ✓ | ✓ | ✓ | ✓ | ✓ |
| Complete physical exam | ✓ | | | | | | | | |
| Targeted physical exam[B] | | ✓ | ✓ | ✓ | ✓ | ✓ | ✓ | ✓ | ✓ |
| Clinical chemistry[C] | ✓ | | ✓ | | ✓ | | | | |
| Hematology[D] | ✓ | | ✓ | | ✓ | | | | |
| Viral serology tests[E] | ✓ | | | | | | | | |
| Urine pregnancy test[F] | ✓ | ✓ | | ✓ | | | | | |
| Unsolicited AEs[G] | ✓ | ✓ | ✓ | ✓ | ✓ | ✓ | ✓ | ✓ | ✓ |
| Concomitant medications[H] | ✓ | ✓ | ✓ | ✓ | ✓ | ✓ | ✓ | ✓ | ✓ |
| Randomization | | ✓ | | ~ | | | | | |
| Blood for humoral immunity | | ✓ | | ✓ | | ✓ | | ✓ | ✓ |
| Blood for CMI[I] | | ✓ | | | | ✓ | | ✓ | |
| Administer study product | | ✓ | | ✓ | | | | | |
| Observation/solicited AEs | | ✓ | | ✓ | | | | | |
| Provide Diary Card | | ✓ | | ✓ | | | | | |
| Review Diary Card | | | ✓ | | ✓ | | | | |
| Exit study | | | | | | | | | ✓ |
| Blood volumes (mL): | | | | | | | | | |
| Safety laboratory tests (Phase 1 only) | 4 | | 4 | | 4 | | | | |
| HBV and HCV screening | 2 | | 0 | | 0 | | | | |
| Humoral immunity | | 10 | | 10 | | 10 | | 10 | 10 |
| Cell-mediated immunity | | 16 | | | | 16 | | 16 | |

~ Confirmation
^ Evaluations will be conducted twice – before and after vaccination
[A] After Visit 6, only medications associated with an AESI, MAAE or SAE will be recorded.
[B] Targeted PE will be conducted only in the event of new symptom, sign or, until Visit 6, any new AE; or, after Visit 6, any new AESI, MAAE or SAE.
[C] Serum creatinine, ALT, AST, total bilirubin; only in Phase 1.
[D] WBC count, hemoglobin, platelets; only in Phase 1.
[E] HBsAg, HCV Ab
[F] Women of childbearing potential (WOCBP) only
[G] After Visit 6, only AESI, MAAEs, and SAEs will be recorded.
[H] After Visit 6, only concomitant medications associated with a newly reported MAAE or SAE will be recorded
[I] Only in subset of subjects in Phase 2

##### 7.2.1. Screening (Day -42 to Day 1)

Once informed consent has been documented, the prospective subject will be considered to be enrolled in the trial and may be screened to determine study eligibility. All inclusion/exclusion criteria must be assessed from data obtained within the screening period, unless otherwise specified in the eligibility criteria. After informed consent has been obtained, the following screening procedures will be performed:

1. Subject identification (ID)/screening number will be assigned.
2. The following information will be obtained from the subject:
    a. Demographic and contact information, including home address, telephone number(s), and email address
    b. Complete medical history of relevance to study eligibility
    c. History of medication use in the past 30 days, and of medications taken that are of specific relevance to study eligibility (e.g., immunosuppressive medications)
    d. Vaccination history
3. Height and weight will be measured, BMI calculation.
4. A full physical examination (PE) will be performed, including vital signs (temperature, pulse rate, and respiratory rate) and assessment of the major organ systems.
5. A urine pregnancy test will be performed for all women of childbearing potential (WOCBP).
6. Venous blood samples (approximately 6 mL in phase 1, and 2 mL in phase 2) will be obtained (non fasting) for clinical laboratory tests, as follows:
    a. White blood cell (WBC) count, hemoglobin (Hgb), platelets (Plt)
    b. Serum creatinine, alanine transaminase (ALT), aspartate transaminase (AST), total bilirubin (t bili)
    c. HBsAg, HCV Ab

Grade 1 vital sign and safety laboratory abnormalities (see Appendix 2 and 3) will not be considered to be exclusionary at screening nor an AE for the study unless judged to be clinically significant by the PI/designee, in consultation with the Sponsor. Prospective subjects may return for repeat assessments once during the screening period to be reassessed for eligibility. The last measurement will be taken as the baseline for purposes of analysis. The PI/designee will use good clinical judgment in considering a prospective subject's overall eligibility.

##### 7.2.2. Visit 1 (Day 1)

At Visit 1, the following procedures will be performed:

1. Subject ID, addresses and telephone number(s) will be confirmed.
2. A urine pregnancy test will be performed for all WOCBP.
3. Unsolicited AEs will be recorded, including assessment of severity and outcome. Ongoing unsolicited AEs will be reviewed.
4. Concomitant medications will be recorded.

5. Vital signs will be measured prior to dosing.
6. Targeted PE will be performed to evaluate any symptom, sign or reported AE, and to confirm absence of exclusionary acute illness or abnormality of the extremity (skin and lymph nodes) targeted for vaccination.
7. Eligibility for vaccination will be confirmed based on review of inclusion/exclusion criteria. Any subject who cannot be vaccinated due to an acute abnormality (e.g., ≥ Grade 2 acute illness, or abnormal vital sign deemed clinically relevant by the PI) may return once the acute issue has resolved, if deemed appropriate by the PI. A minimum of 48 hours must have passed after a documented fever (oral temperature of ≥ 38°C) before a subject can be vaccinated.
8. The subject will be randomized if eligibility is confirmed by the PI.
9. Venous blood samples (approximately 10 mL) will be obtained for immunogenicity testing. For participants in the CMI subset of phase 2, additional 16 mL will be collected.
10. The randomization ID will be recorded by unblinded study staff, who will maintain a list documenting the study vaccine assigned and administered to given randomization IDs in a secure location that is not accessible to blinded study staff. The subject will be referred to by subject ID for the remainder of the study. The randomization ID will be required on select CRFs.
11. The assigned study product will be administered.
12. After the study product is administered, the subject will be observed by blinded study staff for at least 30 minutes post-injection.
13. Vital signs and any solicited AEs will be recorded at 30 (+10) minutes post-injection. See Appendices 1 and 2 for relevant severity grading scales.
14. During the observation period, the subject will be provided with a ruler, thermometer, and Diary Card, and given instructions on how to complete the Diary Card. The subject will be instructed to collect solicited AEs daily for seven days (the day of Visit 1 and the following six days) by recording them in the Diary Card, preferably at approximately the same time each day. The subject will also be instructed to record unsolicited AEs and concomitant medications on the Diary Card. Finally the subject will be instructed to call the PI in the event of any severe (Grade 3) symptoms, or any other health issues that require medical care.
15. The subject will also be provided with a fact sheet on COVID-19 that will include contact details for the PI, and instructions to contact the PI in the event of COVID-19-associated symptoms that meet the following criteria:

    At least ONE of the following:
    - Fever (≥38°C)
    - Shortness of breath
    - Difficulty breathing

    OR

    TWO of the following for TWO or more days:
    - Chills
    - Cough

- Fatigue
- Muscle or body aches
- Headache
- New loss of smell or taste
- Sore throat
- Nausea or Vomiting or Diarrhea

The fact sheet will be reviewed with the subject in clinic prior to discharge.

16. Finally, the subject will be reminded to contact the study team to report any MAAE or SAE.
17. The subject will be scheduled for Visit 2 (Day 8).
18. Safety monitoring within 7 days post study vaccination will be as follows:

- For Phase 1: The study subject will be monitored for safety on site of HMU for 24 hours after study vaccination. The subject will then be discharged for home and will be contacted on Day 4 by telephone or home visit to allow the subject to report any solicited or unsolicited AEs, to address any questions or concerns, and to remind the subject to complete the Diary Card. On Day 6, she/he will be contacted again by telephone or home visit for the same purpose and as a reminder to attend Visit 2 (D8).
- For Phase 2: On the third day (48 hours) post-injection (Day 3), the subject will be contacted by telephone or home visit for a health check, to address any questions or concerns, and to remind the subject to complete the Diary Card. On Day 5, she/he will be contacted again by telephone or home visit for the same purpose and as a reminder to attend Visit 2.

##### 7.2.3. Visit 2 (Day 8) and Visit 4 (Day 36)

At Visit 2 and Visit 4, the following procedures will be performed:

1. Subject ID, addresses and telephone number(s) will be confirmed.
2. Unsolicited AEs will be recorded, including assessment of relatedness to vaccination, severity, and outcome. Ongoing unsolicited AEs will be reviewed.
3. Concomitant medications will be recorded.
4. Vital signs will be measured.
5. Targeted PE will be performed to assess the injection site and draining lymph nodes, and to evaluate any new or ongoing AE.
6. The completed Diary Card will be reviewed with the subject for completeness and accuracy. Possible inaccuracies will be clearly documented. Then completed pages of the Diary Card will be collected, and blank pages and/or additional pages will be returned to subjects for recording any health issue and concomitant medications until the next visit (V3/D29 and V7/D57 respectively).
7. Venous blood samples (approximately 4 mL, only in Phase 1) will be obtained for clinical laboratory tests (non-fasting), as follows:

a. WBC count, Hgb, Plt
b. Serum creatinine, ALT, AST, total bilirubin

8. The COVID-19 fact sheet will be reviewed with the subject (and provided again if needed), including instructions to contact the PI/designee if clinical criteria for COVID-19 are met.
9. The subject will be reminded to contact the study team to report any MAAE or SAE.
10. The date of the subsequent clinic visit will be established.

##### 7.2.4. Visit 3 (Day 29)

At Visit 3, the following procedures will be performed:

1. Subject ID, addresses and telephone number(s) will be confirmed.
2. A urine pregnancy test will be performed for all WOCBP.
3. Unsolicited AEs will be recorded, including assessment of relatedness to vaccination, severity, and outcome. Ongoing unsolicited AEs will be reviewed.
4. Concomitant medications will be recorded.
5. Vital signs will be measured prior to dosing.
6. Targeted PE will be performed to evaluate any new or ongoing AE, and to confirm absence of exclusionary acute illness or abnormality of the extremity (skin and lymph nodes) targeted for vaccination.
7. Eligibility for vaccination will be confirmed based on review of inclusion/exclusion criteria. Any subject who cannot be vaccinated due to an acute abnormality (e.g., ≥ Grade 2 acute illness, or abnormal vital sign deemed clinically relevant by the PI/designee) may return once the acute issue has resolved, if deemed appropriate by the PI/designee. A minimum of 48 hours must have passed after a documented fever (oral temperature of ≥ 38°C) before a subject can be vaccinated.
8. Venous blood samples (approximately 10 mL) will be obtained for immunogenicity testing.
9. The subject's randomization ID will be recorded by unblinded study staff, who will prepare a dose of the assigned study product.
10. The assigned study product will be administered.
11. After the study product is administered, the subject will be observed by blinded study staff for at least 30 minutes post-injection.
12. Vital signs and any solicited AEs will be recorded at 30 (+10) minutes post-injection. See Appendices 1 and 2 for relevant severity grading scales.
13. During the observation period, the subject will be provided with a ruler, thermometer, and Diary Card, and given instructions on how to complete the Diary Card. The subject will be instructed to collect solicited AEs daily for seven days (the day of Visit 3 and the following six days) by recording them in the Diary Card, preferably at approximately the same time each day. The subject will also be instructed to record unsolicited AEs and concomitant medications on the Diary Card. Finally the subject will be instructed to call the PI/designee in the event of any severe (Grade 3) symptoms, or any other health issues that require medical

care.

14. The COVID-19 fact sheet will be reviewed with the subject (and provided again if needed), including instructions to contact the PI/designee if clinical criteria for COVID-19 are met.
15. The subject will be reminded to contact the study team to report any MAAE or SAE.
16. The subject will be scheduled for Visit 4 (Day 36).
17. Safety monitoring within 7 days post study vaccination will be as follows:

    For Phase 1: The study subject will be monitored for safety on site of HMU's hospital for 4 hours after study vaccination. The subject will then be discharged for home and will be contacted on Day 32 by telephone or home visit for a health check, to address any questions or concerns, and to remind the subject to complete the Diary Card. On Day 34, she/he will be contacted again by telephone or home visit for the same purpose and as a reminder to attend Visit 4 (D36).

    For Phase 2: On the third day (48 hours) post-injection (Day 31), the subject will be contacted by telephone or home visit for a health check, to address any questions or concerns, and to remind the subject to complete the Diary Card. On Day 33, she/he will be contacted again by telephone or home visit for the same purpose and as a reminder to attend Visit 4.

###### 7.2.5. Visit 5 (Day 43)

At Visit 5, the following procedures will be performed:

1. Subject ID, addresses and telephone number(s) will be confirmed.
2. Unsolicited AEs will be recorded, including assessment of relatedness to vaccination, severity, and outcome. Ongoing unsolicited AEs will be reviewed.
3. Concomitant medications will be recorded.
4. Vital signs will be measured.
5. Targeted PE will be performed to evaluate any new or ongoing AE.
6. Venous blood samples (approximately 10 mL) will be obtained for immunogenicity testing. For participants in the CMI subset of phase 2, additional 16 mL will be collected.
7. The COVID-19 fact sheet will be reviewed with the subject (and provided again if needed), including instructions to contact the PI/designee if clinical criteria for COVID-19 are met.
8. Finally the subject will be reminded to contact the study team to report any MAAE or SAE.
9. The subject will be scheduled for Visit 6 (Day 57).

###### 7.2.6. Visit 6 (Day 57)

At Visit 6, the following procedures will be performed:

1. Subject ID, addresses and telephone number(s) will be confirmed.
2. Unsolicited AEs will be recorded, including assessment of relatedness to vaccination, severity, and outcome. Ongoing unsolicited AEs will be reviewed.

3. Concomitant medications will be recorded.
4. Vital signs will be measured.
5. Targeted PE will be performed to evaluate any new or ongoing AE.
6. The COVID-19 fact sheet will be reviewed with the subject (and provided again if needed), including instructions to contact the PI/designee if clinical criteria for COVID-19 are met.
7. Finally the subject will be reminded to contact the study team to report any MAAE or SAE.
8. The subject will be scheduled for Visit 7 (Day 197).
9. The subject will receive a text or email every two weeks as a reminder to contact the site to report any MAAE or SAE.

##### 7.2.7. Visit 7 (Day 197)

At Visit 7, the following procedures will be performed:

1. Subject ID, addresses and telephone number(s) will be confirmed.
2. MAAEs or SAEs that have not previously been reported will be recorded, including assessment of relatedness to vaccination, severity, and outcome. Ongoing MAEs and SAEs will be reviewed.
3. Concomitant medications associated with a newly reported MAAE or SAE will be recorded, and ongoing concomitant medications reviewed.
4. Vital signs will be measured.
5. Targeted PE will be performed to evaluate any new or ongoing AE, and intervening MAAE or SAE.
6. Venous blood samples (approximately 10 mL) will be obtained for immunogenicity testing. For participants in the CMI subset, additional 16 mL will be collected.
7. The COVID-19 fact sheet will be reviewed with the subject (and provided again if needed), including instructions to contact the PI/designee if clinical criteria for COVID-19 are met.
8. The subject will be reminded to contact the study team to report any MAAE or SAE.
9. The subject will be scheduled for Visit 8 (Day 365).
10. The subject will receive a text or email every two weeks as a reminder to contact the site to report any MAAE or SAE.

##### 7.2.8. Visit 8 (Day 365, EOS)

At Visit 8, the following procedures will be performed:

1. Subject ID, addresses and telephone number(s) will be confirmed.
2. MAAEs or SAEs that have not previously been reported will be recorded, including assessment of relatedness to vaccination, severity, and outcome. Ongoing MAEs and SAEs will be reviewed.
3. Concomitant medications associated with a newly reported MAAE or SAE will be recorded,

and ongoing concomitant medications reviewed.

4. Vital signs will be measured.
5. Targeted PE will be performed to evaluate any new or ongoing AE, and intervening MAAE or SAE.
6. Venous blood samples (approximately 10 mL) will be obtained for immunogenicity testing.
7. The subject will exit the study after completion of the EOS CRF.

###### 7.2.9. Unscheduled Contacts and Visits

Unscheduled contacts and visits may take place at subject request or as deemed necessary by the PI or site staff at any time during the study. Subjects will be instructed to contact the PI/designee if clinical criteria for COVID-19 are met (as specified on the COVID-19 fact sheet) throughout the entire study period. Subjects will also be instructed to contact the site to report any MAAE or SAE throughout the entire study period. All unscheduled contacts and visits will be documented in the subject's study records and on applicable CRFs.

##### 7.3. Subject Discontinuation

Subject discontinuation from study procedures prior to completion of the last study visit may occur for any of the following reasons:

- Withdrawal (defined as discontinuation initiated by a subject): Participation in the study is strictly voluntary. Subjects have the right to withdraw their consent from study participation at any time and for any reason, without penalty. The subject may also withdraw due to an adverse event.
- PI-initiated: The PI/designee may, at her discretion, discontinue a subject from the study if it is considered to be in the subject's best interest to do so (e.g., for safety concerns), or if the subject does not comply with the study requirements. Any subject who is determined to be infected with SARS-CoV-2 prior to Visit 3 (Day 29) will be discontinued from the study (i.e., will not receive the second study injection) but will continue to be followed for safety assessments through Visit 8 (EOS).
- Lost to follow-up: For subjects who fail to attend scheduled visits, study staff are to make at least three attempts to contact the subject prior to considering the subject as lost to follow-up. These attempts should be recorded in the source documents.
- Sponsor-initiated: For example, if the Sponsor is obliged to end the study for administrative or any other reason, such as a recommendation by the DSMB based upon safety review.

Subjects who discontinue prior to first administration of the study product will be replaced, whereas those withdrawn after first administration of the study product will not be replaced.

The reason for and date of subject discontinuation will be recorded in the source documents and relevant CRF. Before concluding the reason for the subject's discontinuation from the study, the investigator should make every effort to investigate whether an AE may have been related to the subject's discontinuation from the study. For subjects considered lost to follow-up, the discontinuation date for the subject to be captured on the discontinuation CRF page is the date of the subject's last completed study visit.

In the event of subject discontinuation from the study, reasonable efforts should be made to conduct the following procedures (unless subject consent to do so has been withdrawn):

- Update subject contact information.
- Review the Diary Card if still in use prior to discontinuation.
- Update any AEs that remained ongoing at the time of the subject's last visit prior to discontinuation.
- Collect any new reportable AEs and concomitant medications since the subject's last visit, based on the protocol-defined reporting period at the time of discontinuation.
- Perform a targeted PE to evaluate any newly reported AEs since the subject's last visit.

#### 7.4. Concomitant Medications

All concomitant medications will be recorded in source documents from Visit 1 (Day 1) through Visit 5 (Day 57). After Visit 5, only those concomitant medications associated with MAAEs and SAEs will be recorded. Details on concomitant medications to be recorded include the generic and/or trade name, indication, dosage, regimen, route of administration, and start and end dates of the medication.

The following concomitant medications are prohibited during the study; however, they must not be withheld by the treating physician if clinically indicated to treat a subject:

- Any investigational medicinal product other than the study product
- Administration of immunoglobulins or any blood products
- Administration of any long-acting immune-modifying drugs (e.g., infliximab or rituximab) or the chronic administration (defined as more than 14 days) of immunosuppressive medications (includes systemic corticosteroids at doses equivalent to ≥ 0.5 mg/kg/day of prednisone; the use of topical steroids including inhaled and intranasal steroids is permitted).

Use of any prohibited medication must be recorded in the CRF. Whether a subject who uses a prohibited medication will be included in the Per Protocol Population will be evaluated on a case-by-case basis.

#### 7.5. Emergency Unblinding Procedure

In the event of a medical emergency, the PI/designee may require that the blind be broken for the subject experiencing the emergency when knowledge of the subject's treatment assignment may influence the subject's clinical care. Every effort will be made not to unblind the subject unless it is considered absolutely necessary for the welfare of the subject. Prior to unblinding, the investigator is encouraged (to the extent possible, without jeopardizing the subject's health) to contact the Sponsor (or designee) to discuss the decision to break the blind.

Unblinding will occur through the secure IWRS or according to the emergency unblinding SOP, to which there will be 24-hour access. Documentation of the unblinding event will be captured by the IWRS or unblinded staff. In the unlikely event that the IWRS cannot be accessed, a backup process (such as contacting the unblinded pharmacist) will be provided. The PI/designee will be expected to provide a rationale for the necessity of unblinding, based on the expectation that knowledge of the subject's treatment assignment will have a meaningful impact on the subject's medical care in the short term. If a subject's treatment assignment is unblinded, the subject will remain in the study and continue with protocol-defined study visits and procedures, unless there is another reason for subject discontinuation. The decision to unblind will be communicated to all regulatory bodies as required. At the end of the study, documentation of all unblinded subjects (and the rationale for unblinding) will be incorporated into the TMF.

#### 7.6. Management of Pregnancy

All WOCBP will be monitored for pregnancy during the study and for the use of adequate contraception until completion of Visit 6 (Day 57).

If a female subject becomes pregnant after Visit 1 (Day 1) but prior to Visit 3 (Day 29) no further study product will be administered. The subject will be encouraged to complete all remaining study visits for safety assessments alone, and if possible and agreed to by the subject, continue to be followed through the pregnancy outcome. The pregnancy and its outcome will be documented, even if birth occurs after the scheduled end of the study for the subject. The PI/designee is required to notify the Sponsor (or designee) within 24 hours of knowledge of a pregnancy.

#### 7.7. Clinical Procedures

**Vital Signs**

- Temperature will be measured in degrees Celsius (recorded to the nearest 0.1 degree) by oral (sublingual) thermometer following at least 20 minutes of avoidance of food and drink.
- Respiratory rate will be measured in breaths per minute.
- Blood pressure will be measured in millimeters (mm) of mercury and heart rate in beats per minute by automated device or manually on the arm not used for study product administration.
- Vital signs will be graded according to the toxicity grading scale in Appendix 2.

**Height and Weight**

- Height will be measured in centimeters (cm) and recorded to the nearest 0.1 cm.
- Weight will be measured in kilograms (kg) and recorded to the nearest 0.1 kg.

**Complete Physical Examination**

- A complete physical examination will include head, eyes, ears, nose, oropharynx, neck, chest (auscultation), lymph nodes (neck, supraclavicular, axillary), back, abdomen (auscultation and palpation), musculoskeletal, skin (especially hands, arms, injection sites), and nervous system.

**Injection Site Examination**

- Erythema will be examined under standard lighting conditions and measured based on the maximum diameter and recorded to the nearest 0.1 cm.
- Swelling/induration will be examined by palpation and visual inspection under standard lighting conditions; the examiner may temporarily mark the skin at the margins of visible swelling/induration, then measure at the maximum diameter and record the distance to the nearest 0.1 cm.
- Local reactions will be graded according to the toxicity grading scale in Appendix 1.

### 8. SAFETY ASSESSMENT AND REPORTING

The PI/designee is responsible for the detection and documentation of events meeting the criteria and definition of an AE or SAE as provided in this protocol for the duration of the study.

#### 8.1. Collection of Safety Events

AEs will be systematically collected at all clinic visits. Solicited AEs will be assessed in all subjects immediately (30 minutes) after each injection. Subjects will record solicited AEs daily in Diary card for the seven days following each injection, as well as record unsolicited AEs during this period. If a solicited AE is ongoing at a 7-day post-injection follow-up visit (Visit 2 or 4), it will be continued to be followed as per AE monitoring requirements. All unsolicited AEs and Solicited AEs, but occurs after 7 days post-injection and clinically significant safety lab tests will be recorded on the AE CRF.

All AEs, including MAAEs, SAEs and AESI, will be collected through the 28-day post-second-injection follow-up visit (Visit 6). After Visit 6, only MAAEs, SAEs, and AESI will be recorded.

#### 8.2. Definitions

##### 8.2.1. Adverse Event

An adverse event is any untoward, undesired, or unplanned event in the form of signs, symptoms, disease, or laboratory or psychological/physiologic observations occurring in a subject enrolled in the clinical trial. This includes all subjects from whom consent has been obtained whether or not they have yet been randomized and received a study product. The event does not need to be causally related to trial participation or receipt of a study product. An AE is temporally related to participation in the study and will be documented as to whether or not it is considered to be related to vaccine. An AE includes, but is not limited to, the following:

- An intercurrent illness or injury during the course of the study
- Any clinically significant worsening of a preexisting condition

##### 8.2.2. Solicited Adverse Event

Solicited adverse events are pre-specified injection-site-specific local and systemic AEs that occur relatively more frequently, or are known to be associated with, immunization, and which are monitored actively as potential indicators of vaccine reactogenicity. Investigators will not be required to assess causality of solicited AEs if the onset is during the solicitation period.
The following specific solicited AEs will be monitored for this study:

Local solicited AEs assessed at the injection site:

- Pain or tenderness
- Swelling or induration
- Erythema

Systemic solicited AEs:

- Fever (defined as oral temperature ≥ 38°C)
- Headache
- Fatigue or malaise
- Myalgia

- Arthralgia
- Nausea or vomiting

##### 8.2.3. Unsolicited Adverse Event

An unsolicited adverse event is any AE reported spontaneously by the subject, observed by the study staff during study visits or those identified during review of medical records or source documents. Solicited AEs with an onset after the seven-day solicitation period will be considered unsolicited AEs. In the absence of a diagnosis, abnormal physical examination findings or abnormal clinical safety laboratory test results that are assessed by the investigator to be clinically significant will be recorded as an AE.

##### 8.2.4. Medically-Attended Adverse Event

A medically-attended adverse event is an unsolicited AE for which the subject received medical attention, such as during an emergency room visit or a visit to or from medical personnel (e.g., medical doctor).

##### 8.2.5. Adverse Event of Special Interest

An adverse event of special interest (AESI) is an AE of scientific and medical concern specific to the Sponsor's product or program, for which ongoing monitoring and rapid communication by the investigator to the Sponsor can be appropriate. Such an event might warrant further investigation in order to characterize and understand it.

The AESI to be monitored in this study include the following:

- Potential immune-mediated medical conditions (PIMMC), given that vaccination has been associated with autoimmunity (See Appendix 4)
- AEs associated with COVID-19, given the importance to subject safety of managing any occurrence of COVID-19, and the possible association of vaccination with enhancement of COVID-19 (See Appendix 5)

There may be additional PIMMC, AEs associated with COVID-19, or other AESI not included in Appendices 4 and 5 that are established by the Sponsor or regulatory authorities prior to or during conduct of the study.

##### 8.2.6. Protocol-Related Adverse Event

A protocol-related adverse event is an AE that occurs from the time of enrollment until the EOS visit that is not considered to be related to receipt of the study vaccine, but is considered by the PI/designee or the Sponsor to be related to the research conditions, i.e., related to the fact that a subject is participating in the study. For example, a protocol-related AE may be an untoward event occurring during blood sampling or other protocol-specified activity.

##### 8.2.7. Treatment-Emergent Adverse Event

A treatment-emergent AE is defined as an AE that is not present prior to administration of the study product, or, if present prior to the administration of the study medication, increases in intensity after administration of the study medication during the course of the study.

##### 8.2.8. Serious Adverse Event

An SAE is a specific AE that:

- Results in death.
- Is life-threatening.*
- Requires inpatient hospitalization or prolongation of an existing hospitalization.**
- Results in a persistent or significant disability or incapacity.***
- Results in a congenital anomaly or birth defect.

***Life-threatening** refers to immediate risk of death as the event occurred per the reporter. A life-threatening event does not include an event that, had it occurred in a more severe form, might have caused death but, as it actually occurred, did not create an immediate risk of death.

For example, hepatitis that resolved without evidence of hepatic failure would not be considered life-threatening, even though hepatitis of a more severe nature can be fatal. Similarly, an allergic reaction resulting in angioedema of the face would not be life-threatening, even though angioedema of the larynx, allergic bronchospasm, or anaphylaxis can be fatal.

****Hospitalization** is an admission to a health facility in the situation where there is an AE. A period of observation at a clinical trial site is not considered to represent hospitalization for the purposes of SAE reporting. Hospitalization or prolongation of a hospitalization constitutes a criterion for an AE to be serious; however, it is not in itself considered an SAE. In absence of an AE, a hospitalization or prolongation of a hospitalization should not be reported as an SAE by the PI/designee on a SAE form. Such situations include, but are not limited to, the following:

- A hospitalization for a preexisting condition that has not worsened.
- Hospitalization for social reasons.

*****Disability** is defined as a substantial disruption in a person's ability to conduct normal life functions. If there is any doubt about whether the information constitutes an SAE, the information is treated as an SAE.

Medical and scientific judgment should be exercised in deciding whether expedited reporting is appropriate in other situations, such as **important medical events** that may not be immediately life-threatening or result in death or hospitalization but may jeopardize the patient or may require intervention to prevent one of the other outcomes listed in the definition above. These should also usually be considered serious. Examples of such events are intensive treatment in an emergency room or at home for allergic bronchospasm, or blood dyscrasias or convulsions that do not result in hospitalization.

##### 8.2.9. Severity (Intensity) of Adverse Event

The severity of all solicited AEs will be graded from Mild (Grade 1) to Potentially Life Threatening (Grade 4), based on the criteria given in Appendix 1. All AEs leading to death are Grade 5 events. Adverse events are graded based on the worst severity grade during the illness/symptoms. All other unsolicited AEs will be classified as an AE and graded based on the AE severity scale in **Error! Reference source not found.** 6 below. The grading scales for solicited and unsolicited AEs have been derived from the *Division of AIDS (DAIDS) Table for Grading the Severity of Adult and Pediatric Adverse Events* (Version 2.1, July 2017), from the US National Institutes of Health.

**Table 6. Severity Grading**

| Grade | Description |
|---|---|
| 1 | Causes no or minimal interference with normal daily activities; intervention not indicated |
| 2 | Interferes with but does not prevent normal daily activities; intervention indicated |
| 3 | Prevents normal daily activities; intervention or hospitalization indicated |
| 4 | Causes inability to perform basic self-care activities; intervention indicated to prevent permanent impairment, persistent disability, or death |

##### 8.2.10. Causal Relationship of an Adverse Event

A suspected adverse drug reaction (ADR) means any AE for which there is a reasonable possibility that the study vaccine caused the AE. A reasonable possibility means there is evidence to suggest a causal relationship between the vaccine and the AE. All cases judged by either the PI/designee or the Sponsor as having a reasonable suspected causal relationship to the study vaccine will qualify as ADRs. Medical judgment will be used to determine the relationship, considering all relevant factors, including pattern of reaction, temporal relationship, confounding factors such as concomitant medication, concomitant diseases, and relevant history.

The likelihood of the relationship of the AE to study vaccine is to be recorded as follows:

- Related: There is a reasonable causal relationship between the vaccine administered and the AE.
- Not Related: There is no reasonable causal relationship between the vaccine administered and the AE.

Note: solicited reactogenicity events will not be judged for relatedness.

##### 8.2.11. Assessment of Outcome of Adverse Event

The outcome of the AE will be assessed and recorded as per the following categories:

- Ongoing.
- Recovered/resolved.
- Recovered/resolved with sequelae.
- Fatal.
- Unknown.

#### 8.3. Adverse Event Recording and Reporting

Recording and reporting of all AEs will occur from signing of the ICF (enrollment) through 28 days post-second injection (Visit 6) for each study subject receiving study product. SAEs, MAAEs, and AESI will be recorded throughout the study period. The Study staff must completely and promptly record each AE in the source documentation and on the AE CRF, regardless of relationship to the vaccine administered/procedure as determined by the PI/designee. The PI/designee will attempt, if possible, to establish a diagnosis based on the signs and symptoms. When a diagnosis for the reported signs or symptoms is known, the PI/designee will report the diagnosis as the AE, not the signs and symptoms. Adverse events will be classified by MedDRA term and by severity/intensity, relatedness, and outcome.

Enrolled subjects who subsequently screen fail (i.e., who never underwent randomization) or withdraw consent will have any AEs recorded from enrollment until the time they are determined to be ineligible for randomization or withdraw consent. These AEs will be listed in separate appendices from those subjects randomized and vaccinated. For the purposes of data capture they will be closed at the point the subject is deemed ineligible.

Reporting of AEs will follow the regulatory guidelines of the Ministry of Healthand NIHE's EC in regard to requirements, processes and forms.

##### 8.4. Serious Adverse Event Reporting

If an AE is classified as serious, an SAE form will be completed and submitted within 24 hours of the PI/designee becoming aware of the SAE, including information on the location, severity, relatedness, and clinical summary of the event to the Sponsor to initiate any PSRT evaluation and any additional reporting requirements. In addition, the SAE submission will follow the regulatory guidelines of the Ministry of Health andNIHE's IRB in regards to requirements, processes and forms. It is the responsibility of the Sponsor to ensure that Dynavax Technologies Corporation (manufacturer of the adjuvant CpG 1018) and the other Consortium manufacturers are notified of SAEs and other notable safety events per agreed procedures and timelines. Any SAE deemed related to study vaccine that is ongoing at the time of last subject last visit (LSLV) will continue to be followed until it is resolved, assessed to be resolved with sequelae, or assessed to be stable/chronic. SAEs deemed not related to study vaccine that are unresolved at the time of LSLV will be classified as ongoing.

##### 8.5. Unanticipated Problems

All unanticipated problems will be reported in the continuing review report submitted to the Ministry of Healthand NIHE's IRB per reporting requirements of each regulatory body. All serious unanticipated problems involving risk to participants or others will be promptly (within 48 hours) reported by telephone, by email, or by facsimile to the Sponsor. Follow-up reports will be submitted as soon as additional information becomes available.

#### 9. SAFETY MONITORING

The PI/designee will be responsible for continuous monitoring of all study subjects' safety. In case of urgent need, subjects will have the means to get in contact with study staff at any time (24 hours per day). The PI/designee will also be available by cell phone 24-hours per day for medical emergencies.

##### 9.1. Protocol Safety Review Team

Safety will be monitored routinely throughout the study by the PSRT, which includes the PI/designee and other site investigators, the IVAC Clinical Lead, and the Medical Monitor. The PSRT will be continuously involved in safety monitoring and be available to address any urgent medical queries or safety concerns related to any subject's participation in the study. Data management personnel will ensure that the PSRT receives immediate notification of all reported SAEs and any other predefined

AEs (e.g., Grade 3 AEs). The PSRT will convene to review blinded safety data on a weekly basis until D43 of the last subject in each phase, then at least once monthly until the end of the study.

During meetings the PSRT will review blinded safety reports and review any outstanding medical or safety queries from the previous review. The blinded safety reports will contain at a minimum, subject disposition and discontinuations, all new Grade 3 and persistent (beyond 6 days post-IP administration) solicited AEs, and all new unsolicited AEs. Cumulative safety data reports will also be made available continuously for review by PSRT members. In addition, the PSRT will review all major protocol deviations in an expedited manner and all other protocol deviations at least on a monthly basis to assess for any potential safety implications, and will provide guidance in the preparation of corrective action plans. The PSRT may also discuss any other study conduct issues that impact study integrity and subject safety, including but not limited to data quality and critical monitoring findings. The PSRT may refer any safety concerns to the DSMB. The full responsibilities and procedures of the PSRT will be detailed in the Safety Management Plan.

#### 9.2. Data and Safety Monitoring Board

A DSMB, composed of at least three independent members with expertise in vaccine clinical trials, will be convened to provide additional safety oversight. In Phase 1, the DSMB will meet to review unblinded safety data through Day 57 (V6), and on the basis of this review provide a recommendation for advancing to Phase 2 (i.e. the DSMB will indicate whether advancement to Phase 2 is warranted based on the reviewed safety data, and whether specific COVIVAC formulations should not be considered for advancement due to safety concerns). In the Phase 2 segment, the DSMB will again meet to review unblinded safety data through Day 57 (V6), and on the basis of this review provide a recommendation for advancing to Phase 3 (i.e. the DSMB will indicate whether there are any safety concerns with one or the other selected COVIVAC formulation evaluated in Phase 2).

The DSMB will also meet if a study pause rule is met, or the PSRT elects to implement a study pause. If this occurs, no further enrollment will occur, and no study product will be administered until the DSMB approves lifting the pause based on unblinded review of all safety data accrued during the trial. If the study is permanently terminated, subjects who have already received study product will continue with all scheduled protocol visits and assessments; however, they will not receive any further study product.

DSMB reviews will indicate whether or not safety concerns were identified, and whether the trial should continue without change, be modified, or be terminated. The Sponsor will carefully consider the DSMB recommendations. If the Sponsor does not agree with these recommendations, a meeting will be held between the Sponsor, PI, and DSMB to reach consensus on the appropriate action(s) to take in regard to the trial. However, if attempts to reach consensus fail, the Sponsor's opinion will prevail. In such situations, the Sponsor will inform the MoHand NIHE's EC of the Sponsor's perspective, and any changes to the trial. The PSRT, PI or IVAC Clinical Lead may also seek additional guidance from the DSMB as dictated by the occurrence of certain events that do not warrant a study pause.

The composition, responsibilities and procedures of the DSMB will be detailed in the DSMB Charter.

#### 9.3. Pause Rules

The DSMB will be convened if it is established that any of the following study pause rules has been met during the conduct of the trial:

- **Rule 1**: Occurrence of any SAE attributed to study vaccine in ≥ 1 subject
- **Rule 2**: Occurrence of the following Grade 3 or greater injection site solicited AE in ≥ 1 subject: ulceration, necrosis, or sterile abscess at the injection site requiring drainage or surgical intervention.
- **Rule 3**: Occurrence of the same Grade 3 solicited AE, unsolicited AE attributed to study vaccine, or laboratory abnormality in ≥ 10.0% of unique subjects. In the case of fever, pain and tenderness, the episode must last longer than 24 hours, and, in the case of fever, be confirmed by the PI/designee without evidence of other medical causes. Note: for the first 20 randomized subjects, this pause rule will only be met if the grade 3 AE or laboratory abnormality occurs in ≥ 2 unique subjects.

##### 9.4. Protocol Deviation

A protocol deviation is any noncompliance with the clinical trial protocol, GCP, site SOP requirements, or departure from applicable regulatory requirements. The noncompliance may be either on the part of the subject or the site team/PI.

Major protocol deviations are a subset of protocol deviations that may significantly impact the completeness, accuracy, and/or reliability of the study data or that may significantly affect a subject's rights, safety, or well-being. Examples of major protocol deviations may include: failure to obtain informed consent, failure to report SAEs, enrolling subjects in violation of key eligibility criteria designed to ensure a specific subject population, or failing to collect data necessary to interpret primary endpoints, as this may compromise the scientific value of the trial..

When appropriate, corrective actions and preventive actions (CAPAs) will be developed by the site to address deviations, and will be implemented promptly. These practices will be consistent with ICH E6 Guidelines.

#### 10. STATISTICAL CONSIDERATIONS

##### 10.1. Overview and General Considerations

This is a randomized, placebo-controlled, observer-blind, two-stage Phase 1/2 trial in healthy male and non-pregnant female subjects of age 18-75 years old, inclusive. A maximum of 120 eligible subjects will be accrued in the first stage (Phase 1), and a maximum of 300 eligible subjects will be accrued in the second stage (Phase 2).

The first stage is a first-in-human study in 120 healthy subjects of age 18-59 years old designed to evaluate the COVIVAC vaccine at three different dose levels (1 µg, 3 µg, 10 µg) without adjuvant and one dose level (1 µg ) with the adjuvant CpG 1018. Randomization into each group will be age stratified with half of subjects between 18-39 years of age and the other half 40-59 years of age.

In the second stage of this combined Phase 1/2 study, 300 adults aged 18-75 years will be randomized (2:5:5) to placebo, or one of two selected formulations of COVIVAC being evaluated in first phase. To ensure that adequate safety and immune data will be available from older and elderly adults to inform the selection of COVIVAC formulation to advance to Phase 3 studies, randomization will be age stratified by group with approximately one third of subjects in 18-39, 40-59, and 60-75 years of age.

##### 10.2. Randomization Procedures

Randomization for Part 1 (Phase 1) will be conducted after accumulation of 120 eligible subjects. Randomization will be stratified by gender and age (less than 40 years old and 40 to 59 years old, with approximately 50% of subjects in each age stratum). This is to ensure the groups are as similar (homogenous) as possible to ensure the down-selection of formulations is not biased by differences in demographic characteristics of the groups (such as age and gender). Randomization details will be outlined in a Randomization Plan prior to study initiation.

Each group will be divided into two subgroups, a sentinel subgroup (n=6) and a larger subgroup (n=24) that includes the remaining subjects to be evaluated at that dose level (or for that formulation in the case of the one adjuvanted treatment group). Initially, a sentinel subgroup of 6 subjects will be randomized to receive COVIVAC 1 µg or placebo in a 5:1 ratio. Sentinel subjects will be monitored for reactogenicity and safety through Day 8 (Visit 2), including for the initial 24 hours post-injection in clinic. If the PSRT has no safety concerns based on blinded review of all safety data, including clinical laboratory results, through Visit 2, the second subgroup of 24 subjects will be randomized to receive COVIVAC 1 µg or placebo at the same ratio; on the same day, the sentinel subgroup (n=6) for the second dose level to be evaluated (COVIVAC 3 µg) can be randomized to receive study vaccine or placebo in a 5:1 ratio, and so on for the subsequent dose level and formulation (see Table 4). The same procedure will be used for the COVIVAC 10 µg and COVIVAC 1 µg + CpG arms for a total of n=25 in each active group and n=20 in the placebo group. Randomization will be stratified by age (less than 40 years old and 40 to 59 years old) with approximately 50% of subjects in each stratum.

After the interim analysis of data from the first phase informs the down selection to two candidate groups 300 adults aged 18-75 years will be randomized (2:5:5) to placebo, or one of two selected formulations of COVIVAC being evaluated in Phase 1. Twelve (12) subjects in each of the three Phase 2 groups (equally distributed between the three age strata) will be selected for CMI at the time of randomization. Randomization will be age stratified by group with approximately one third of subjects in 18-39, 40-59, and 60-75 years of age.

##### 10.3. Sample Size and Power

This Phase 1/2 study has a two-part selection design with group elimination after the first part (i.e. Phase 1). A total of 25 ($n_1$) subjects per group will be randomized across 4 candidate and one placebo group ($n_1$=20) in the first phase. Assay titers will be analyzed on the log scale. The sample size and power calculations assume an interim analysis will be conducted after D43 of the first phase to eliminate two candidate groups by selecting the two groups with the largest means. Additionally, the observed seroresponse rate (defined as the percentage of subjects in a treatment group with at least a 4-fold rise from baseline in 80% neutralizing antibody titers) will need to be at least 52% at the lower limit of the 95% exact confidence interval. Based on this criterion, there is less than 10% chance of incorrectly failing to consider advancing a vaccine candidate with a true seroresponse rate of 85% or higher, which is estimated to be the minimum response rate that would be consistent with further investment in an additional second-wave COVID-19 vaccine. The Sponsor may consider other factors in selecting the two most suitable candidates to advance to second phase. In the second phase, 125 and 50 ($n_2$) additional subjects will be randomized to the two candidate groups and the placebo group, respectively. The final analysis will be conducted on the full samples from the first and second phase ($n_1 + n_2$).

Safety: Assuming 25 evaluable subjects per group in the first phase (and for elderly subjects enrolled in the second phase) and 150 total subjects ($n_1 + n_2$) in the two candidate groups included in the second phase, the probability of observing at least one serious or severe adverse event by the underlying rate is shown in Table 7. If no events are observed the upper bound of the exact 95% confidence interval would be 13.7% for the three eliminated groups and 2.4% in the two selected groups.

**Table 7. Probability to observe at least one serious or severe adverse event by sample size and underlying event rate.**

| Sample size per group | True Event Rate | Probability to observe at least one |
|---|---|---|
| 25 | 10.0% | 92.8% |
| 25 | 7.5% | 85.8% |
| 25 | 5.0% | 72.3% |
| 25 | 2.5% | 46.9% |
| 150 | 10.0% | 99.9% |
| 150 | 7.5% | 99.9% |
| 150 | 5.0% | 99.9% |
| 150 | 2.5% | 97.8% |
| 150 | 1.0% | 77.9% |
| 150 | 0.5% | 52.9% |

Immunogenicity: Groups with seroresponse below 52% at the lower limit of the 95% exact confidence interval will not be considered for selection for the second phase. This provides 90% power to correctly advance candidates with a true seroresponse rate ≥85%.

This is a two-stage selection design for selecting the group with the largest mean. Power for selecting the candidate with the largest true mean is driven both by the variability and the smallest difference between the highest mean and the means in the other three candidate groups.

Table 8 shows the power to correctly select the candidate with the largest true mean in the first phase (to advance to the second phase) and the overall power for the Phase 1/2 by minimum fold-difference (on the titer scale) between the highest mean and the other means, assuming log10 variability of 0.5. These calculations assume in the first phase there are $n_1$=22 evaluable subjects in the four candidate groups, two groups are eliminated after the first phase, there are $n_2$=112 additional evaluable subjects in the remaining two candidate groups at D43 of second phase ($n_1 + n_2$=134), and $\mu_4 > \mu_1 = \mu_2 = \mu_3$.

**Table 8. Power to select the candidate with the highest true mean to progress to second phase and overall for the Phase 1/2 overall by minimum fold-difference between the highest candidate mean and the other means, assuming log10 variability of 0.5.**

| Fold-difference | Phase 1 down-selection power | Power for overall Phase 1/2 |
|---|---|---|
| 1.1 | 62% | 46% |
| 1.2 | 73% | 66% |
| 1.3 | 82% | 78% |
| 1.4 | 87% | 86% |
| 1.5 | 91% | 91% |
| 1.6 | 94% | 94% |
| 1.7 | 96% | 96% |
| 1.8 | 98% | 98% |
| 1.9 | 99% | 99% |
| 2.0 | 99% | 99% |

#### 10.4. Definitions of Populations to be Analyzed

##### 10.4.1. Enrolled Population

All subjects who provide written informed consent, regardless of the subject's screening, randomization, and treatment status in the study.

##### 10.4.2. Exposed Population

All subjects in the enrolled population who were randomized and received at least one vaccination dose (i.e., were accrued).

##### 10.4.3. Safety Analysis Population

All subjects in the exposed population for whom any safety data is available. All safety analyses will be performed using this population. The denominators for different safety endpoints may vary according to the number of subjects with available data for the specific endpoint. For instance, the solicited local and systemic adverse event endpoint will be based only on those who have the corresponding CRF data regardless of other safety data. Treatment groups for safety analysis will be assigned according to the actual treatment received.

##### 10.4.4. Full Analysis Population

All subjects in the exposed population for whom any post-study product administration immunogenicity results are available. An immunogenicity analysis will also be performed using this population.

###### 10.4.5. Per Protocol Population

All subjects in the exposed population who have no major protocol deviations that are determined to potentially interfere with the immunogenicity assessment of the study product.

This population will serve as the primary analysis population for the immunogenicity endpoints. The population will be adapted by time point to include all eligible subjects' data up to the time of disqualifying protocol deviation. The criteria (e.g., intake of prohibited medication expected to influence the immune response) for exclusion of subjects from the per protocol population, and determination of any exclusions based on a blinded review of the data, will be established before database freeze and lock for both interim and final analyses, respectively.

##### 10.5. Analysis Sequence and Analytical Methodology

Interim analyses will be performed after the last subject in the first phase has completed Visit 6 (Day 57) assessments and all the results are available. At the time of the interim analysis safety results (unblinded to the DSMB only) and immunogenicity results (unblinded at group level only) will aid in the selection of formulations to evaluate in the second phase of the study. Appendix 6 is a template of the interim study report that will be generated based on this analysis.

A second interim analysis will be performed after the last subject has completed Visit 6 (Day 57) assessments and all the results are available. The second interim analysis of immunogenicity and safety endpoints will aid in the selection of a formulation to be evaluated in a future Phase 3 trial.

A final analysis on all safety and immunogenicity data will be performed after the study ends, when all additional safety and immunogenicity data have been collected following the last subject's last study visit (Visit 8, Day 365), and the database is cleaned and locked. Detailed statistical procedures, listings, table shells, and figures will be provided in a separate statistical analysis plan (SAP) which will be finalized prior to the interim analyses.

Unless otherwise specified, all statistical test will be two-sided with a significance level of 0.05. A 95% CI will be provided for estimates, as appropriate. All statistical analyses will be performed using SAS® software version 9.4 or later.

###### 10.5.1. Analysis of Demographic and Baseline Characteristics

Baseline demographics and characteristics, including age, height, weight, sex, race ethnicity, and BMI will be summarized for both the exposed and per protocol populations by treatment group using descriptive statistics (mean, standard deviation, median, $25^{th}$ and $75^{th}$ quartiles, minimum and maximum for continuous and rates for categorical).

For the exposed population, medical history will be listed and summarized by category. Using the WHO Drug Dictionary, concomitant medications will be tabulated by anatomical therapeutic chemical (ATC) classification, preferred drug name and treatment group. Medical history will be tabulated by MedDRA System Organ Class (SOC), Preferred Term (PT) and treatment group.

Summaries of subject disposition will be prepared for all subjects, including the number and percent enrolled, screened, randomized, and administered study product, as well as a CONSORT diagram describing study participation and discontinuation. The reasons for screen failures and discontinuations will be summarized and listed.

A summary and listing of visit attendance will be prepared, in addition to a summary and listing of study product administration and sample collection/availability for each sample.

##### 10.5.2. Analysis of Primary Objectives

All safety assessment will take place in the safety analysis population, according to the treatment received. All subject-level percentages (solicited/unsolicited AEs, clinical safety laboratory abnormalities, etc.) will be supplemented with two-sided 95% CIs computed via the Clopper-Pearson method. Individual summaries (denominators for percentages) will be limited to the number of subjects with the appropriate analysis population with the data available for analysis for the given endpoint. Summaries will be provided overall by group and by stage and age strata.

Solicited local and systemic adverse events: All solicited AEs will be summarized according to defined severity grading scales. Frequencies and percentages of subjects experiencing each solicited AE will be presented for each symptom severity. Summary tables showing the occurrence of any local or systemic solicited AE during the first 7 days after each administration of study product. Data listings of all solicited AEs will be provided by subject. For common AEs group comparisons may be explored.

Clinically significant hematological and biochemical measurements: For clinical safety laboratory data collected at screening and 7 days post-first administration of study product (Visit 2, Day 8) and post-second administration of study product (Visit 4, Day 36), individual Hgb, WBC count, Plt, AST, ALT, total bilirubin, and creatinine values will be presented as number of subjects out of range (above and below normal range as appropriate) and tabulated by toxicity grading, relatedness, and study group. In addition, changes from baseline (median/interquartile range) will be presented.

Unsolicited AEs: All unsolicited AEs with onset occurring during the first 28 days following study product administration will be assessed for severity and as either related or not related to study product by the investigator.

MAAEs and SAEs: All MAAEs and SAEs through Visit 8 (Day 365) will be recorded and the number, severity, and relatedness to study product will be summarized. Subject-wise data listing will be provided.

When an AE occurs more than once for a subject, the subject will be only counted once for the corresponding PT according to the maximum severity of the events. A summary table will be prepared for unsolicited AEs comprised of the following categories:

- Unsolicited AEs
- Related unsolicited AEs
- MAAEs
- Related MAAEs
- SAEs
- Related SAEs
- SAEs leading to death
- Unsolicited AEs leading to subject discontinuation

Adverse events of special interest: The number, severity, and relatedness of adverse events of special interest (AESI), including

- PIMMC

- AEs specific to COVID-19

will be summarized and provided in subject-wise data listings.

##### 10.5.3. Analysis of Secondary Objectives

The analysis of immunogenicity will be performed on the per protocol population as the primary analysis and the full analysis population as a secondary analysis. Analyses described below will be summarized in the interim analyses for all subjects included in the first phase of the study. In the second interim and final analyses, the candidates that advanced to the second phase will be summarized by phase, age group (18-59 years and 60-75 years), and overall. Relative endpoints derived from GMFR will include all subjects from the per protocol population. Absolute endpoints such as those derived from the GMT will exclude baseline seropositive subjects.

Immunogenicity data will be descriptively analyzed. For estimation of the GMT, GMT ratio, and corresponding confidence limits, analyses will incorporate censoring where appropriate and age-adjusted log-scale coefficients will be back-transformed in order to compute the estimate and corresponding confidence limits for the relevant quantity. GMFRs will be computed using age-adjusted estimates of the log difference of the paired samples, with corresponding CIs computed via the *t*-distribution, utilizing the antilog transformation to present the ratio. Analyses of binary variables will include 95% CIs computed via the Clopper-Pearson method. Additionally, the distribution of the titers will be summarized using reverse cumulative distribution curves.

NAb titers against SARS-CoV-2 pseudovirus will be measured by pseudovirus-based neutralization assay and summarized at baseline, 28 days after the first vaccination, and 14 days, 6 months and 12 months after the second study injection overall and by age group (18-59 years and 60-75 years) with:

- GMT with 95% CIs (including baseline),
- GMFR from baseline with 95% CI, and
- Percentage of subjects with $NT_{50}$ and $NT_{80}$ seroresponses against SARS-CoV-2 pseudovirus at 28 days after the first vaccination, and 14 days, 6 months and 12 months after the second vaccination as defined by
  - a ≥ 4-fold increase from baseline, and
  - a ≥ 10-fold increase from baseline.

Anti-S IgG antibody titers will be measured by ELISA and summarized at baseline, 28 days after the first vaccination, and 14 days, 6 months and 12 months after the second study injection overall and by age group (18-59 years and 60-75 years) with:

- GMTs with 95% CIs (including baseline),
- GMFR from baseline with 95% CI, and
- Percent of subjects with IgG seroresponses and 95% CI of
  - ≥ 4-fold increase from baseline
  - ≥ 10-fold increase from baseline

##### 10.5.4. Analysis of Exploratory Objectives

The S protein-specific T cell response elicited by each of two selected COVIVAC formulations will be measured via ELISpot assay at baseline, 14 days, and 6 months after the second vaccination in a subset of subjects enrolled in the second phase of the study. The magnitude, phenotype, and cytokine-expressions pattern of S protein-specific T cells will be descriptively analyzed by timepoint (absolute and relative to baseline) by study arm. The anti-NDV HN IgG immune response elicited by each formulation of COVIVAC will be measured by ELISA, at baseline, 28 days after the first vaccination, and 14 days and 12 months after the second vaccination, in subjects enrolled in the second phase of the study, and be summarized with GMTs with 95% CIs.

Other exploratory assays to characterize the immune response to COVIVAC may also be performed. Details of all exploratory analyses will be described in the SAP.

##### 10.6. Multiplicity

As the two-stage selection design relies on ranking of GMTs, not testing of differences between candidate groups, no adjustment for multiplicity will be performed.

##### 10.7. Handling of Subject Discontinuations and Missing Data

Missing immunogenicity data will not be imputed and will be analyzed as if they were randomly missing. Over the study period, the frequency and percentage of subjects who discontinue from the study will be provided by treatment group. All subjects who discontinue post-randomization will be further described regarding their time to and their reasons for discontinuation. For subjects who discontinue from the study, their data collected before discontinuation will be analyzed under the analysis populations as applicable.

#### 11. LABORATORY EVALUATIONS

##### 11.1. Blood Sample Collection, Distribution, and Storage

Venous blood samples obtained for clinical laboratory tests will be processed and labeled at the site, and sent to the lab of HMU's hospital. Blood samples obtained to evaluate secondary immunogenicity objectives will be processed, and serum samples aliquoted, labeled, and stored at -70°C in freezers, as per instructions provided in the Laboratory Manual, before being tested at Nexelis (Quebec, Canada). All stored serum samples will be logged into a secure database.

The volume of blood required for the different categories of assays at different visits are shown in Table 9.

**Table 9. Blood Volume Requirements**

| **Lab Panels**<br>Tests | **Screen** | **Visit 1** | **Visit 2** | **Visit 3** | **Visit 4** | **Visit 5** | **Visit 6** | **Visit 7** | **Visit 8** |
|---|---|---|---|---|---|---|---|---|---|
| **Hematology[a]:**<br>WBC, hgb, plts | 2.5 mL | - | 2.5 mL | - | 2.5 mL | - | - | - | - |
| **Viral Serology:**<br>HBsAg, HCV Ab | 2 mL | - | - | - | - | - | - | - | - |
| **Chemistry[a]:**<br>creatinine, AST, ALT, t bili | 1.5 mL | - | 1.5 mL | - | 1.5 mL | - | - | - | - |
| **Humoral Immunity:** | - | 10 mL | - | 10 mL | - | 10 mL | - | 10 mL | 10 mL |

| **Lab Panels** Tests | **Screen** | **Visit 1** | **Visit 2** | **Visit 3** | **Visit 4** | **Visit 5** | **Visit 6** | **Visit 7** | **Visit 8** |
|---|---|---|---|---|---|---|---|---|---|
| PNA, ELISAs | | | | | | | | | |
| **CMI:*** ELISpot | - | 16 mL | - | - | - | 16 mL | - | 16 mL | - |
| **Total Volume (+ CMI)** | **6 mL** | **10 mL (26 mL)** | **4 mL** | **10 mL** | **4 mL** | **10 mL (26 mL)** | **-** | **10 mL (26 mL)** | **10 mL** |

[a] Only in Phase 1

* Subset of subjects in Phase 2

##### 11.2. Safety Clinical Laboratory Assays

Protocol-mandated screening and clinical safety laboratory tests will be performed at the clinical laboratory of HMU's hospital, which is certified with International Organization for Standardization (ISO) 15189.

If clinically significant abnormalities are identified during screening, subjects will be referred for further medical management. If identified during the study, subjects may be asked to return to the study clinic for further evaluation, including clinical evaluation and repeat laboratory testing as warranted.

##### 11.3. Immunological Assays (Secondary Objectives)

Assays to evaluate secondary immunogenicity objectives will be performed at Nexelis, as follows:

**SARS-CoV-2 pseudovirus neutralization assay (PNA)**

The PNA will be used to quantify the functional humoral immune response against SARS-CoV-2 by measuring the 50% and 80% neutralizing titer ($NT_{50}$ and $NT_{80}$) against a SARS-CoV-2 pseudovirus. The pseudotyped virus particles are made using a genetically modified Vesicular Stomatitis Virus that expresses the wild-type spike protein of SARS-CoV-2 (Wuhan strain) and contains a luciferase reporter which can be quantified in relative luminescence units (RLU). $NT_{50}$ and $NT_{80}$ are determined based on the dilution of serum required to achieve 50% and 80% reduction in RLU value compared to pseudoparticle control. The assay has been qualified.

**SARS-CoV-2 pre-fusion Spike IgG ELISA**

The ELISA assay detects IgG antibodies to the full-length pre-fusion spike protein of SARS-CoV-2. Serum antibody content is expressed in ELISA Units (ELU)/ml. The assay has been validated.

At the completion of all the testing, the samples will be either destroyed or stored at an appropriate place in a designated freezer at Nexelis or at a Sponsor-designated facility. The Sponsor will be responsible for the oversight of sample storage and destruction.

##### 11.4. Microbiological Assays

Nasal/pharyngeal swabs will be collected from subjects who experience signs and/or symptoms of COVID-19 that are of sufficient severity to warrant testing for SARS-CoV-2 infection. The testing will be performed at the Laboratory of virus transmitted from animal to human, Virus Lab Department of NIHE, a CLIA-certified diagnostics laboratory, using a singleplex RT-PCR assay. In addition to detecting presence of virus, the assay also provides the PCR cycle threshold (Ct) value, which may be used as an estimate of viral load. When swabs are received at the laboratory, they will be processed and then divided into two aliquots. The first aliquot will be used to test for presence of SARS-CoV-2, with results available on the day the swab was received. The second aliquot will be stored in a designated freezer for possible additional testing, including for other respiratory viral pathogens if judged to be helpful in the clinical management of individual cases.

##### 11.5. Assay Qualification, Standardization, and Validation

Assays employed to evaluate the primary safety laboratory endpoint have been properly validated and will be run with adequate controls. Immunological assays have been validated, or qualified, and are run with adequate controls.

##### 11.6. Biohazard Containment

As transmission of blood-borne pathogens can occur through contact with contaminated needles, blood, and blood products, appropriate blood and secretion precautions will be employed by all personnel in the drawing of blood and handling and shipping of all specimens for this study, as recommended by the US Centers for Disease Control and Prevention. All dangerous goods materials, including diagnostic specimens and infectious substances, must be transported according to instructions detailed in the International Air Transport Association (IATA) Dangerous Goods Regulations. Biohazardous waste will be contained according to institutional, transportation/carrier, and all other applicable regulations.

#### 12. DATA MANAGEMENT

The Sponsor-designated CRO’s study monitors will visit the site at regular intervals, as per the Monitoring Plan, and perform pre-agreed source data verification of the data recorded in the electronic data capture (EDC) system against the source documents available at the site. In addition, missing data forms and fields will be queried by daily electronic edit checks or through manual edit checks of the data by the data management team. Study monitors will closely evaluate pre-screening data, inclusion/exclusion criteria, informed consents, data entry timeliness, and visit dates and windows to ensure integrity of the study is maintained.

Any data discrepancies generated by the system will be flagged in the EDC system for the site staff to provide a satisfactory resolution within the EDC system. The data management team will review all the data discrepancy responses by the site to ensure the correctness of data. The medical history events and the AEs will be coded using MedDRA dictionary version 21.1 or later and the concomitant medications will be coded using the WHO Drug Dictionary. After completion of data coding and resolution of all the queries in the database, the database will be declared to be complete and accurate and will be locked for final statistical analysis of primary and secondary endpoints.

##### 12.1. Case Report Form Development and Completion

Based on the final protocol of the study, a comprehensive set of CRFs will be prepared to capture all the relevant data required for analysis and reporting. This study will utilize an EDC system such that the entire study data can be maintained in a secure electronic system. No written or electronic data recorded prior to the study will be included in the CRFs or EDC system respectively.

All study data will be collected by the clinical study staff using designated source documents and will be entered in the EDC system in an anonymized form. The study database will identify study subjects only by unique study identification numbers through screening (subject ID) and randomization assignments (randomization ID) and will not contain any identifying information such as name, address or personal contact information, or any other regional/state/national identification number. CRFs will be reviewed by the clinical team who are responsible for ensuring that they are accurate and complete.

The data management activities will be performed as per the CRO's SOPs. The appropriately trained site personnel will ensure that the study data recorded in the EDC system is verifiable with the source documents available at the site. To ensure that data are entered in a timely fashion so as to monitor safety of the study, it is expected that the site will maintain data entry with a minimal expectation of three business days from subject clinic visit. The study monitor plan will include assessments of data entry timeliness.

The study site will maintain the source documents for each study subject. The source documents and other supporting documents will be kept in a secure location. Source documentation will be available for review by the study monitor to ensure that the collected data are consistent with the CRFs.

#### 12.2. Record Archival

##### 12.2.1. Archiving Data at Study Site

The study site will maintain appropriate medical and research records for this trial, in compliance with ICH GCP, regulatory, sponsoring organization and institutional requirements for the protection of confidentiality of subjects. The site will permit authorized representatives of the Sponsor and regulatory agencies to examine (and when required by applicable law, to copy) clinical records for the purposes of quality assurance reviews, audits, and evaluation of the study safety and progress. After completion of data coding and resolution of all the queries in the database, the database will be declared to be complete and accurate and will be locked for final statistical analysis and made available to the Sponsor for long-term storage in a master file.

##### 12.2.2. Data Storage and Archival

The study monitor will provide the PI with an Investigator Site File, which will be used to file the investigator's brochure (IB), protocol, drug accountability records, correspondence with the EC, Sponsor, CRO, and other study-related documents. The PI/designee will maintain, and store securely, complete, accurate and current study records throughout the study.

As required by ICH GCP guidelines, the PI will keep essential documents until at least two years after the last approval of a marketing application in an ICH region, and until there are no pending or contemplated marketing applications in an ICH region, or at least two years have elapsed since the formal discontinuation of clinical development of the study product. These documents should be retained for a longer period, however, if required by the applicable regulatory requirements or by an agreement with the Sponsor. The documents will be archived either at Clinical Trial Center at NIHE or at any other secure location as agreed upon with the Sponsor. It is the responsibility of the Sponsor to inform the PI/institution as to when these documents no longer need to be retained. Subjects' medical records and other original data will be archived in accordance with the local regulations or facilities of the investigational site.

##### 12.3. Posting of Information on Clinicaltrials.gov

Study information from this protocol will be posted on Clinicaltrials.gov.

##### 12.4. Confidentiality

Documented evidence that the PI is aware and agrees to the confidential nature of the information related to the study must be obtained by means of a confidentiality agreement.

All information provided by the Sponsor and all data and information generated by the site as part of the study (other than a subject's medical records) will be kept confidential by the PI and other site staff. This information and data will not be used by the PI or other site personnel for any purpose other than conducting the study.

##### 12.5. Publication

IVAC will work with the PI and other relevant personnel at NIHE on the publication of the complete Phase 1/2 study outlined in this protocol in a timely fashion. Primary publication of the trial results will be shared between the NIHE and IVAC. Other individuals having input into the study justifying authorship will similarly be included in publication(s). Additional publications resulting from the analysis of the study data will be agreed between IVAC and the NIHE on a case-by-case basis but will generally include authors from both organizations. IVAC will be acknowledged in all publications as the Sponsor of the trial.

If a written contract for the conduct of the study, which includes publication provisions inconsistent with this statement, is executed between IVAC and the study site, that contract's publication provisions shall apply rather than this statement.

#### 13. QUALITY ASSURANCE AND QUALITY CONTROL

##### 13.1. General Considerations

The study will be conducted in full compliance with the protocol and ICH GCP to provide public assurance that the rights, safety, and well-being of study subjects are protected, and that the clinical study data are credible. To ensure quality and standardization, the site will develop SOPs for key protocol procedures and conduct the study guided by the study and applicable site SOPs and other written guidelines. The site will also develop routine operational checks to verify that critical protocol requirements and procedures are executed correctly and completely at the time the work is being performed. Prior to study initiation, the Sponsor and the designated CRO will conduct training on the protocol and applicable SOPs for the study staff.

The investigational site will provide direct access to all study-related documents, source data/documents, and reports for monitoring and auditing by the Sponsor, and inspection by regulatory authorities.

##### 13.2. External Monitoring

Sponsor monitoring responsibilities will be provided by the CRO. A site initiation visit will be conducted prior to beginning the study, and monitoring will be conducted at initiation, during, and at closeout of the study by the study monitor or designee.

During the course of the study, the monitor will assess site activities at intervals to verify compliance to the protocol; completeness, accuracy, and consistency of the data and study product accountability; adherence to protocol and regulatory obligations; and to ensure that conduct of the research follows GCP. The extent and frequency of the monitoring visits will be described in a separate Clinical Monitoring Plan developed prior to study initiation. The monitor should have access to subject medical records, study product accountability and other study-related records needed to verify the entries on the CRFs.

The PI and the monitor will cooperate to ensure that any problems detected in the course of these monitoring visits, including EDC completion and query resolution, are resolved in a predefined time frame described in the Clinical Monitoring Plan.

To ensure the quality of clinical data for all subjects, a clinical data management review will be performed on subject data received by the CRO. During this review, subject data will be checked for consistency, omissions, and any apparent discrepancies. In addition, the data will be reviewed for adherence to the protocol and GCP. To resolve any questions arising from the clinical data management review process, data queries and/or site notifications will be sent to the site for resolution as soon as possible and within the time frame described in the Clinical Monitoring Plan; all queries must be resolved prior to database lock.

##### 13.3. Independent Auditing

Sponsor representatives may audit the study to ensure that study procedures and data collected comply with the protocol and applicable SOPs of the site and the CRO, and that data are correct and complete. The PI will permit auditors (employees of the Sponsor or an external company designated by the Sponsor) to verify source data validation of the regularly monitored clinical study. The auditors will compare the entries in the CRFs with the source data and evaluate the study site for its adherence to the clinical study protocol and GCP guidelines and applicable regulatory requirements.

##### 13.4. Regulatory Agency Auditing

The PI must be aware that representatives from regulatory authorities may wish to inspect the CRFs and associated study records. The PI will notify the Sponsor within 24 hours following contact by a regulatory agency. The PI will make the relevant records available for inspection and will be available to respond to reasonable requests and audit queries made by authorized representatives of regulatory agencies. The PI will provide the Sponsor with copies of all correspondence that may affect the review of the current study or her/his qualification as PI in clinical studies conducted by the Sponsor. The Sponsor will provide any needed assistance in responding to regulatory audits or correspondence.

#### 14. OBLIGATIONS AND ROLES OF THE SPONSOR, PI, AND STUDY PERSONNEL

This study will be conducted according to GCP as well in accordance with Ministry of Health regulations. The Sponsor will assure the trial is conducted in compliance with the protocol, GCP, and regulatory authority requirements. The Sponsor will provide the PI with the funding and information needed to conduct the trial properly, ensuring proper monitoring of trial activities, and that the trial is conducted in accordance with the general investigational plan and protocol contained in the submissions to the regulatory authorities. The Sponsor will ensure that the PI and regulatory authorities are immediately informed (within 24 hours of the Sponsor becoming aware) of significant new adverse effects or risks with respect to the study vaccine. The Sponsor will ensure that they will be immediately informed (within 24 hours of awareness) of any other safety concerns which could influence decisions regarding informed consent, enrollment and vaccination in this trial.

The PI agrees to perform the research in strict accordance with this protocol, the ICH GCP (E6), as well as in conformity with local regulations regarding the conduct of clinical studies (see Statement of Compliance).

In addition, the PI will follow local and institutional requirements including, but not limited to, investigational vaccines, clinical research, informed consent and ethics regulations. The Sponsor will provide notification to the PI of protocol and amendment approvals by regulatory authorities when applicable. Any modifications to the research protocol, the ICF, and/or change in PI will be submitted for review and approval to regulatory authorities per their guidelines. The PI may deviate from the protocol without prior approval only when the deviation is necessary to eliminate an apparent immediate hazard to the study subject.

While the PI may delegate study duties to appropriate study personnel, the PI is ultimately responsible for the conduct of all aspects of the study.

#### 15. ETHICAL CONSIDERATIONS AND INFORMED CONSENT

The study will be performed in accordance with SOPs generated and agreed between the Sponsor-designated CRO and the PI. The CRO has the responsibility for ensuring the site has the appropriate SOPs to perform the study and the authority to defer to the site SOP rather than the CRO SOP when appropriate. These SOPs have been developed in accordance with ICH Guidelines for GCP E6 R2 (2018), Directive 2001/20/EC, which are consistent with the Ethical Guidelines outlined in the Declaration of Helsinki (2013), thus ensuring protection of the subjects. The study will commence only after receipt of a favorable opinion from the EC listed in this protocol and Ministry of Health.

##### 15.1. Ethical Review

Ethical review of this study for the clinical site will be jointly conducted by the Ministry of Health IRBand NIHE EC. The Sponsor will enter into a reliance agreement with ECs for ethical review. The PI is responsible for obtaining approval from the Ministry of Health IRBand NIHE EC, which will review and approve the protocol, the informed consent form, and any recruitment materials (advertising or informational material). This includes any modifications to these documents prior to, or during the study. Any change to the protocol or informed consent form must be reviewed and approved prior to implementation, except when necessary to eliminate apparent immediate hazard to study subjects. In such a case, the change must be later documented in an amendment and reported to the ECs as soon as possible. When a change involves only logistical or administrative aspects of the study (e.g., change of a telephone number), formal EC approval may not be required, but such amendments shall still be submitted to the ECs for information purposes, and the PI must provide the Sponsor with written confirmation that such logistical or administrative amendments have been submitted to the ECs. The PI will provide the Sponsor with a statement from the ECs confirming that the ECs are organized and operates according to GCP and applicable laws and regulations. The PI is also responsible for obtaining continuing review throughout the duration of the study in accordance with existing regulations

##### 15.2. Informed Consent Process

Informed consent is a process that is initiated prior to the individual agreeing to participate in the study and continues throughout the subject's participation in the study. Before any study-relatedactivities, the PI must ensure that the potential subject is fully informed about the objectives, procedures, potential risks, and potential benefits of study participation.

Potential subjects will be provided with the EC-approved ICF, allowed ample time to read the consent form, encouraged to ask questions about the study, have their questions answered and thenbe given time to decide if they would like to participate in the study. If the subject is illiterate, there will be a witness or legally appropriate representative attending the consent process to assure the subject is properly consented. It will be emphasized that their participation is voluntary, and that they have the right to decline to participate or to discontinue from the study at any time without giving any reason and without compromising their other rights in any way.

The PI must obtain the subject's voluntary, signed and dated ICF before any study-related procedures are performed. Study staff must document the informed consent process. The original, signed ICF must be kept in the site study file. A copy of the informed consent document will be given to the subject for their records.

##### 15.3. Study-Related Injury

IVAC will maintain product liability insurance to cover treatment for study-related injuries. IVAC will also maintain employer liability insurance or shall self-insure, as necessary, to meet its liability obligations under this protocol, as well as sufficient levels of all legally mandated insurance, including (at a minimum) professional liability coverage for the investigator, trial team, and all other employees, contractors, and agents providing services to this trial. As applicable, IVAC will maintain insurance to cover any general liability and product liability to meet its obligations under Vietnam law.

##### 15.4. Risk/Benefit

There are certain symptoms and signs, systemic or injection site, which have historically been associated with immunization, and if they do occur, usually are of mild or moderate intensity and transient in nature. Consequently, in early clinical trials of investigational vaccines, these symptoms and signs are typically considered as risks and are solicited as potential AEs in the first week following immunization. In this trial, the solicited injection site AEs include pain, tenderness, swelling or induration, and erythema. The solicited systemic AEs are fever, headache, fatigue or malaise, myalgia, arthalgia, and nausea or vomiting. Since there may be unknown risks associated with the COVIVAC vaccine, all subjects will be monitored for any unsolicited AEs for 28 days following study product administration and for any MAAEs or SAEs for the entire study period. All subjects will also be monitored for specific AESI; these include potential immune-mediated medical conditions, which may be associated with vaccination. In addition subjects will be closely monitored for adverse events associated with COVID-19 to identify, and treat if indicated, potential cases of COVID-19 during the trial, and to assess for possible vaccine-induced immune enhancement of disease.

CpG 1018 is the adjuvant used in HEPLISAV-B vaccine, which was licensed by the US FDA in 2018. HEPLISAV-B was well-tolerated and shown to have an adequate safety profile in clinical trials in adults.[49] Common adverse reactions from HEPLISAV-B included injection site pain, erythema, and swelling, headache, malaise, myalgia, and fatigue. SAEs, autoimmune adverse events, and deaths were infrequent in the HEPLISAV-B clinical program and similar in frequency between HEPLISAV-B and the active comparator (Engerix-B). No deaths have been considered by the investigator to be related to HEPLISAV-B. Interim analyses of an ongoing post-marketing study of over 30,000 HEPLISAV-B recipients and more than 35,000 active comparator recipients have raised no safety concerns.

Hypersensitivity reactions may occur following the administration of any vaccine, including licensed vaccines, which in rare circumstances may be life-threatening. All subjects will be informed in the ICF of this possibility following study vaccine administration and will be observed for a minimum of 30 minutes following study vaccine administration. Appropriate emergency medical treatment will be available in case of severe immediate reactions, such as anaphylaxis. Subjects with a known hypersensitivity to any component of the study vaccine, or a history of hypersensitivity to any vaccine, will be excluded from the study.

Blood drawing- and venipuncture-associated risks may include minor bleeding or bruising at the venous access site, mild discomfort, upset stomach, dizziness, light-headedness, syncope, or very rarely infection. Blood samples will only be drawn by trained staff members using aseptic technique, and medical assistance will be available in case of any complications. Subjects will be informed of these risks in the ICF and will be in a seated or supine position during blood draws.

No benefits can be guaranteed to subjects for their participation in this research study. Subjects who participate in this study may benefit from the clinical assessments (e.g., medical history, physical examination, and routine clinical safety laboratory tests) conducted at screening and during the study, including the screening for infections. If the subject is found to have any newly diagnosed medical condition or infection, the investigator will ensure that the subject is provided with appropriate and adequate referrals within the health care system. The information gained from this study may be useful in the development of a safe and effective vaccine to prevent COVID-19 for which there is currently no licensed vaccine. Subjects may gain from knowing that they have been involved in this development process. The known risks of participation in this study are believed to be outweighed by the value of the information to be gained.

#### 15.5. Subject Confidentiality

Every effort will be made to protect subject privacy and confidentiality. Personal identifiers will not be included in any study reports. Medical records containing identifying information will be made available for review when the study is monitored by the Sponsor or an authorized regulatory agency. Direct access may include examining, analyzing, verifying, and reproducing any records and reports that are important in the evaluation of the study.

All study-related information will be stored securely at the study site. All subject information will be stored in locked file cabinets in areas with access limited to study staff. Data collection, process, and administrative forms, and other reports will be identified only by a unique trial-related subject identification code (subject/randomization ID) to maintain subject confidentiality. Laboratory reports may include the name and date of birth of the subject to minimize the risk of errors in the busy clinical laboratories. All local databases will be secured with password-protected access systems.

Forms, lists, logbooks, appointment books, and any other listings that link subject ID numbers to other identifying information will be stored in a separate, locked file in an area with limited access. Subjects' study information will not be released without their written permission, except as necessary for monitoring, or as required/permitted by law/regulatory authorities.

##### 15.6. Reimbursement

Pending EC approval, subjects will be compensated for their time and effort in this study. The study ICF will state the plan for reimbursement. Subjects will not be charged for study injections, research clinic visits, research-related examinations, or research-related laboratory tests.

##### 15.7. Storage of Specimens

Stored study research samples will be labeled by a code that only the study site can link to the study subject. All stored research samples will be logged into a secure database and any use documented. Samples may be stored at the clinical sites and laboratories included in the contact cover page of this protocol, to complete the analyses required to meet study primary, secondary and exploratory analyses. As a part of the informed consent process, subjects will be informed of and asked to agree to long-term storage of specimens for use in future, related research.

When samples are no longer needed for the purposes of the study, they will be kept or destroyed, depending on whether subjects consent to remaining samples being used in future research. Their use will be governed by a repository plan that is mutually agreeable to the Sponsor and NIHE. No genetic testing will be done on the samples.

### 16. APPENDICES

#### 16.1. Appendix 1: Solicited Injection Site and Systemic Reactions Toxicity Grading Table

| Reaction to Injectable Product | Mild (Grade 1) | Moderate (Grade 2) | Severe (Grade 3) | Potentially Life Threatening (Grade 4) |
|---|---|---|---|---|
| **Pain/ Tenderness** | Does not interfere with activity | Some interference with activity OR Repeated use of non-narcotic pain reliever > 24 hours | Significant; prevents daily activity OR Any use of narcotic pain reliever | Emergency room (ER) visit or hospitalization |
| **Erythema[a]** | 2.5 – 5 cm | 5.1 – 10 cm | > 10 cm and/or ulceration OR secondary infection OR phlebitis OR sterile abscess OR drainage | Necrosis or exfoliative dermatitis |
| **Swelling/ Induration[a]** | 2.5 – 5 cm and does not interfere with activity | 5.1 – 10 cm OR Interferes with activity | > 10 cm OR prevents daily activity and/or ulceration OR secondary infection OR phlebitis OR sterile abscess OR drainage | Necrosis |
| **Temperature (oral)** | 38.0 – 38.4°C 100.4 – 101.1°F | 38.5 – 38.9°C 101.2 – 102.0°F | 39.0 – 40°C 102.1 – 104°F | > 40°C > 104°F |
| **Headache** | No interference with activity | Some interference with activity OR Repeated use of nonnarcotic pain reliever >24 hours | Significant; prevents daily activity OR Any use of narcotic pain reliever | ER visit or hospitalization |
| **Fatigue/ Malaise** | No interference with activity | Some interference with activity | Significant; prevents daily activity | ER visit or hospitalization |
| **Myalgia** | No interference with activity | Some interference with activity | Significant; prevents daily activity | ER visit or hospitalization |
| **Arthralgia** | No interference with activity | Some interference with activity | Significant; prevents daily activity | ER visit or hospitalization |
| **Nausea/ Vomiting** | No interference with activity | Some interference with activity | Prevents daily activity OR Requires outpatient IV hydration | ER visit or hospitalization for hypotensive shock |

[a] For erythema and swelling/induration, longest diameter should be noted in centimeters. Swelling/induration should be evaluated and graded using the functional scale as well as the actual measurement.

The grading scales for local and systemic AEs have been derived from *Guidance for Industry, Toxicity Grading Scale for Healthy Adult and Adolescent Volunteers Enrolled in Preventive Vaccine Clinical Trials*, U.S. Department of Health and Human Services, FDA, CBER, September 2007( https://www.fda.gov/media/73679/download) and the *Division of AIDS (DAIDS) Table for Grading the Severity of Adult and Pediatric Adverse Events* (Version 2.1, July 2017) from the US NIH (https://rsc.niaid.nih.gov/sites/default/files/daidsgradingcorrectedv21.pdf).

#### 16.2. Appendix 2: Vital Signs Toxicity Grading Table

| Vital Signs[a] | Mild (Grade 1) | Moderate (Grade 2) | Severe (Grade 3) | Potentially Life Threatening (Grade 4) |
|---|---|---|---|---|
| **Tachycardia – beats per minute** | 101 – 115 | 116 – 130 | > 130 | Hospitalization for arrhythmia |
| **Bradycardia –beats per minute[b]** | 50 – 54 | 45 – 49 | < 45 | ER visit or hospitalization for arrhythmia |
| **Hypertension (systolic) – mm Hg** | 141 – 150 | 151 – 165 | > 165 | ER visit or hospitalization for malignant hypertension |
| **Hypertension (diastolic) – mm Hg** | 91 – 99 | 100 – 105 | > 105 | ER visit or hospitalization for malignant hypertension |
| **Hypotension (systolic) – mm Hg** | 85 – 89 | 80 – 84 | < 80 | ER visit or hospitalization for hypotensive shock |
| **Respiratory Rate – breaths per minute** | 21 – 23 | 24 – 27 | > 27 | Intubation |

[a] Subject should be at rest for all vital sign measurements.
[b] Grade 1 bradycardia or grade 1 tachypnea will not be considered an abnormality for this study, unless judged to be clinically significant by the PI in consultation with the Sponsor

The grading scales for abnormal vital signs have been derived from *Guidance for Industry, Toxicity Grading Scale for Healthy Adult and Adolescent Volunteers Enrolled in Preventive Vaccine Clinical Trials*, U.S. Department of Health and Human Services, FDA, CBER, September 2007.

#### 16.3. Appendix 3: Serum and Hematology Toxicity Grading Table

| **Serum** | **Mild (Grade 1)** | **Moderate (Grade 2)** | **Severe (Grade 3)** | **Potentially Life Threatening (Grade 4)** |
|---|---|---|---|---|
| Creatinine (mg/dL) | 1.1 – 1.3 x ULN | > 1.3 – 1.8 x ULN OR increase of > 0.3 above baseline | > 1.8 – < 3.5 x ULN OR increase of 1.5 – < 2.0 x above baseline | ≥ 3 .5 x ULN OR increase of > 2.0 x above baseline |
| Liver function tests – AST, ALT increased | 1.25 – < 2.5 x ULN | 2.5 – <5.0 x ULN | 5.0 – < 10.0 x ULN | ≥ 10.0 x ULN |
| Total bilirubin | 1.1 – < 1.6 x ULN | 1.6 – < 2.6 x ULN | 2.6 – < 5.0 x ULN | ≥ 5.0 x ULN |
| **Hematology** | **Mild (Grade 1)** | **Moderate (Grade 2)** | **Severe (Grade 3)** | **Potentially Life Threatening (Grade 4)** |
| Hemoglobin [Female] (g/dL) | 11.0 – 12.0 | 9.5 – 10.9 | 8.0 – 9.4 | < 8.0 |
| Hemoglobin [Male] (g/dL) | 12.5 – 13.5 | 10.5 – 12.4 | 8.5 – 10.4 | < 8.5 |
| OR Hemoglobin, change from baseline value (g/dL) | 1.0 – 1.5 | 1.6 – 2.0 | 2.1 – 5.0 | > 5.0 |
| WBC, increased (cell/$mm^3$) | 10,800 – 15,000 | 15,001 – 20,000 | 20,0001 – 25,000 | > 25,0000 |
| WBC, decreased (cell/$mm^3$) | 2,500 – 3,400 | 1,500 – 2,499 | 1,000 – 1,499 | < 1,000 |
| Platelets, decreased (cell/$mm^3$) | 100,000 – <125,000 | 50,000 – <100,000 | 25,000 – < 50,000 | < 25,000 |

Abbreviation: ULN = upper limit of normal range

Note: the laboratory values provided in this table serve as guidelines and are dependent upon institutional normal parameters.

The grading scales for laboratory abnormalities have been derived from *Guidance for Industry, Toxicity Grading Scale for Healthy Adult and Adolescent Volunteers Enrolled in Preventive Vaccine Clinical Trials*, U.S. Department of Health and Human Services, FDA, CBER, September 2007, and the *Division of AIDS (DAIDS) Table for Grading the Severity of Adult and Pediatric Adverse Events* (Version 2.1, July 2017) from the US NIH.

#### 16.4. Appendix 4: Potential Immune-Mediated Medical Conditions (PIMMC)

| Categories | Diagnoses |
|---|---|
| Neuroinflammatory Disorders: | Acute disseminated encephalomyelitis (including site specific variants: e.g., non-infectious encephalitis, encephalomyelitis, myelitis, myeloradiculomyelitis), cranial nerve disorders including paralyses/paresis (e.g., Bell's palsy), generalized convulsion, Guillain-Barre syndrome (including Miller Fisher syndrome and other variants), immune-mediated peripheral neuropathies and plexopathies (including chronic inflammatory demyelinating polyneuropathy, multifocal motor neuropathy and polyneuropathies associated with monoclonal gammopathy), myasthenia gravis, multiple sclerosis, narcolepsy, optic neuritis, transverse myelitis, uveitis |
| Musculoskeletal and Connective Tissue Disorders: | Antisynthetase syndrome, dermatomyositis, juvenile chronic arthritis (including Still's disease), mixed connective tissue disorder, polymyalgia rheumatic, polymyositis, psoriatic arthropathy, relapsing polychondritis, rheumatoid arthritis, scleroderma (including diffuse systemic form and CREST syndrome), spondyloarthritis (including ankylosing spondylitis, reactive arthritis [Reiter's Syndrome] and undifferentiated spondyloarthritis), systemic lupus erythematosus, systemic sclerosis, Sjogren's syndrome |
| Vasculidities: | Large vessels vasculitis (including giant cell arteritis such as Takayasu's arteritis and temporal arteritis), medium sized and/or small vessels vasculitis (including polyarteritis nodosa, Kawasaki's disease, microscopic polyangiitis, Wegener's granulomatosis, Churg–Strauss syndrome [allergic granulomatous angiitis], Buerger's disease [thromboangiitis obliterans], necrotizing vasculitis and anti-neutrophil cytoplasmic antibody [ANCA] positive vasculitis [type unspecified], Henoch-Schonlein purpura, Behcet's syndrome, leukocytoclastic vasculitis) |
| Gastrointestinal Disorders: | Crohn's disease, celiac disease, ulcerative colitis, ulcerative proctitis |
| Hepatic Disorders: | Autoimmune hepatitis, autoimmune cholangitis, primary sclerosing cholangitis, primary biliary cirrhosis |

| **Categories** | **Diagnoses** |
|---|---|
| Renal Disorders: | Autoimmune glomerulonephritis (including Immunoglobulin A nephropathy, glomerulonephritis rapidly progressive, membranous glomerulonephritis, membranoproliferative glomerulonephritis, and mesangioproliferative glomerulonephritis |
| Cardiac Disorders: | Autoimmune myocarditis/cardiomyopathy |
| Skin Disorders: | Alopecia areata, psoriasis, vitiligo, Raynaud's phenomenon, erythema nodosum, autoimmune bullous skin diseases (including pemphigus, pemphigoid and dermatitis herpetiformis), cutaneous lupus erythematosus, morphoea, lichen planus, Stevens-Johnson syndrome, Sweet's syndrome |
| Hematologic Disorders:<br><br>Metabolic Disorders:<br><br>Other Disorders: | Autoimmune hemolytic anemia, autoimmune thrombocytopenia, antiphospholipid syndrome, thrombocytopenia<br><br>Autoimmune thyroiditis, Grave's or Basedow's disease, Hashimoto thyroiditis[A], diabetes mellitus type 1, Addison's disease<br><br>Goodpasture syndrome, idiopathic pulmonary fibrosis, pernicious anemia, sarcoidosis |

[A]New onset only

This list was derived from Tavares Da Silva F, De Keyser F, Lambert PH, Robinson WH, Westhovens R, Sindic C. Optimal approaches to data collection and analysis of potential immune mediated disorders in clinical trials of new vaccines. *Vaccine*. 2013 Apr 3;31(14):1870-6.

#### 16.5. Appendix 5: Adverse Events Specific to COVID-19 Disease

| Body System | Diagnoses |
|---|---|
| Immunologic | Enhanced disease following immunization<br>Multisystem inflammatory syndrome in children (MIS-C) |
| Respiratory | Acute respiratory distress syndrome (ARDS) |
| Cardiac | Acute cardiac injury including:<br>• Microangiopathy<br>• Heart failure and cardiogenic shock<br>• Stress cardiomyopathy<br>• Coronary artery disease<br>• Arrhythmia<br>• Myocarditis, pericarditis |
| Hematologic | Coagulation disorder<br>• Deep vein thrombosis<br>• Pulmonary embolus<br>• Cerebrovascular stroke<br>• Limb ischemia<br>• Hemorrhagic disease<br>• Thrombotic complications |
| Renal | Acute kidney injury |
| Gastrointestinal | Liver injury |
| Neurologic | Guillain Barré Syndrome<br>Anosmia, ageusia<br>Meningoencephalitis |
| Dermatologic | Chilblain-like lesions<br>Single organ cutaneous vasculitis<br>Erythema multiforme |

This listing is based on Safety Platform for Emergency Vaccines (SPEAC*) D2.3 Priority List of Adverse Events of Special Interest: COVID-19*, V2.0 Date May 25, 2020. https://brightoncollaboration.us/wp-content/uploads/2020/06/SPEAC_D2.3_V2.0_COVID-19_20200525_public.pdf

#### 16.6. Appendix 6: Template of interim study report for transition to Phase 2

1. **Cover pages (**according to the CSR template by MOH VN)

- Study title, protocol number
- Key individuals and institution involving the study.
- Timeline for study implementation for Phase 1

2. **Protocol synopsis**

3. **Brief Investigational plan**

4. **Study Participant disposition until D43**

- Enrollment/screening counts: total participants screened/enrolled/screen failures/Early termination
- Screening failure summary: illegible, withdrawn consent, study no longer for enrollment, others.
- Study progress: number of subject attending ICF/screening, V1/D1, V2/D8, V3/D29, V4/D36, V5/D43, V6/D57 visits
- Protocol deviations

5. **Demographic and key baseline characteristics**

6. **Immunogenicity analysis till cutoff date**

- Humoral (functional) immune response

7. **DSMB safety recommendation (unblinded)**

8. **Safety analysis till cutoff date (blinded)**

- Local solicited AEs
- Systemic solicited AEs
- Unsolicited AEs
- SAEs
- MAAEs
- AESI

9. **Discussion and recommendation of two selected doses for Phase 2**

#### 16.7. Appendix 7: References

[1] Zhu N, Zhang D, Wang W, Li X, Yang B, Song J, et al. A novel coronavirus from patients with pneumonia in China, 2019. N Engl J Med 2020.

[2] WHO. Middle East respiratory syndrome coronavirus (MERS-CoV) MERS Monthly Summary, November 2019. https://www.who.int/emergencies/mers-cov/en/. Accessed 22 May 2020.

[3] Lu, R., Zhao, X., Li, J., Niu, P., Yang, B., Wu, H., Wang, W., Song, H., Huang, B., Zhu, N., *et al.* (2020). Genomic characterisation and epidemiology of 2019 novel coronavirus: implications for virus origins and receptor binding. Lancet. 2020 02 22; 395(10224):565-574.

[4] Johns Hopkins University & Medicine Coronavirus Resource Center. https://coronavirus.jhu.edu/map.html Accessed 6 January 2020.

5 Cowling BJ, Ali ST, Ng TWY, et al. Impact assessment of non-pharmaceutical interventions against coronavirus disease 2019 and influenza in Hong Kong: an observational study. *Lancet Public Health*. 2020;5(5):e279-e288.

[12] Levinson R, Elbax M, Ben-Ami R, Shasha D, Levinson T, Choshen G, Petrov K, Gadoth A, Paran Y, Anosmia and dysgeusia in patients with mild SARS-CoV-2 infection. medRxiv 2020.04.11.20055483

[13] Wu Z, McGoogan JM. Characteristics of and Important Lessons From the Coronavirus Disease 2019 (COVID-19) Outbreak in China: Summary of a Report of 72 314 Cases From the Chinese Center for Disease Control and Prevention [published online ahead of print, 2020 Feb 24]. *JAMA*. 2020;10.1001/jama.2020.2648.
